# From menarche to menopause: the impact of reproductive factors on the metabolic profile of over 65,000 women

**DOI:** 10.1101/2022.04.17.22273947

**Authors:** Gemma L Clayton, Maria Carolina Borges, Deborah A Lawlor

**Affiliations:** MRC Integrative Epidemiology Unit, University of Bristol, Bristol, UK; Population Health Sciences, Bristol Medical School, University of Bristol, Bristol, UK; NIHR Bristol Biomedical Research Centre, Bristol, UK

## Abstract

We explored the relation between age at menarche, parity and age at natural menopause with 249 metabolic traits, measured using nuclear magnetic resonance (NMR), in up to 65,487 UK Biobank women using multivariable regression (MV), Mendelian randomization (MR) and a male negative control (parity only). Older age of menarche was related to a less atherogenic metabolic profile in MV and MR, which was largely attenuated when accounting for adult body mass index. In MV, higher parity related to complex changes in lipoprotein-related traits; these were not observed in male negative controls and were imprecisely estimated in MR. In MV and MR, older age at natural menopause was related to lower concentrations of inflammation markers, but inconsistent results were observed for LDL-related traits due to chronological age-specific effects. Our findings support a role of reproductive traits on later life metabolic profile and provide insights into identifying novel markers for the prevention of adverse cardiometabolic outcomes in women.

**Summary box:** *What is new?:*

- Markers of women’s reproductive health are associated with several common chronic conditions. Whilst some attempts have been made to explore the extent to which these associations are causal, metabolites could act as mediators of the relationship between reproductive markers and chronic diseases.
- Older age of menarche was related to a less atherogenic metabolic profile in multivariable regression and Mendelian randomization, however, this was largely attenuated when accounting for adult body mass index.
- In multivariable regression, higher parity related to complex changes in lipoprotein-related traits. Whilst these were not observed in male negative controls, suggesting a potential causal effect in females, they were not replicated in the Mendelian randomization, possibly due to imprecise estimates.
- Older age at natural menopause was related to lower concentrations of inflammation markers in both multivariable regression and Mendelian randomization. Consistent results were observed for LDL-related traits when stratified by chronological age.

*Implications:*

- Given that the age at menarche results were largely attenuated to the null when accounting for adult BMI, it is likely that age at menarche itself may not causally relate to the metabolic profile.
- These results, particularly for parity and age at menopause, could contribute to identifying novel markers for the prevention of adverse cardiometabolic outcomes in women and/or methods for accurate risk prediction. For example, consistent with other studies, higher parity was associated with unfavourable (e.g. higher number of particles and lipid content in VLDL and higher glycine) changes in the metabolic profile. Similarly, older age at menopause was related to higher lipid content in HDL particles and lower systemic inflammation, as proxied by GlycA.

## Introduction

Markers of women’s reproductive health, such as age at menarche, parity and age at menopause, have been associated with several common chronic conditions, including cardiometabolic diseases ^1-6^ and breast, ovarian and endometrial cancer ^7-12^. Some attempts have been made to explore the extent to which these associations are causal, as opposed to explained by residual confounding, using approaches such as Mendelian randomization (MR) and negative control designs, which are less prone to bias by key confounders from conventional observational studies. MR studies suggest a direct positive effect of age at menarche on breast cancer and an indirect inverse effect via body mass index (BMI) ^13^, as well as a possible bidirectional relationship between age at menarche and BMI ^13,14^. MR also supports a protective effect of older age at first birth on type 2 diabetes and cardiovascular diseases ^15^ and lower mean levels of BMI, fasting insulin and triglycerides in women and men ^16^, while a partner negative control study provides some evidence of a ‘J-shaped’ effect of parity on coronary heart disease risk ^5^. In addition, evidence from MR studies indicate that older age at menopause increases the risk of breast, endometrial and ovarian cancer, reduces the risk of bone fractures and type 2 diabetes, and do not substantially affect BMI or cardiovascular diseases risk ^17^.

Metabolites could act as mediators of the relationship of reproductive markers, and related hormonal changes, with chronic diseases ^18-20^. Determining the effect of women’s reproductive markers on multiple metabolites would be the first step to exploring this and could provide crucial insights into mechanisms underlying women’s long-term health. We have previously shown marked changes in metabolites, such as lipids, fatty acids, amino acids and inflammatory markers during pregnancy ^20^, through the menopausal transition ^21^, and among women on hormonal contraceptives containing estrogen ^22^. Many of these same metabolic measures are also related to cardiovascular diseases ^19^ and some cancers ^23-26^. The aim of this paper is to explore the extent to which women’s reproductive markers have a causal effect on 249 metabolic measures (covering lipids, fatty acids, amino acids, glycolysis, ketone bodies and an inflammatory marker). We focus on three reproductive traits that represent key events in women’s reproductive lives: (i) age at menarche, a marker of puberty timing, (ii) parity, a marker of repeated exposure to the physiological challenges of pregnancy, and (iii) age at menopause, a marker of reproductive aging. We explore the causal relationships between reproductive markers and metabolic measures by triangulating evidence ^27^ across multivariable regression, a negative control design (for parity only), and MR (**Figure 1**). Given each of these approaches has unique strengths and limitations, results that agree across them are less likely to be spurious ^27^.

**Fig 1.**
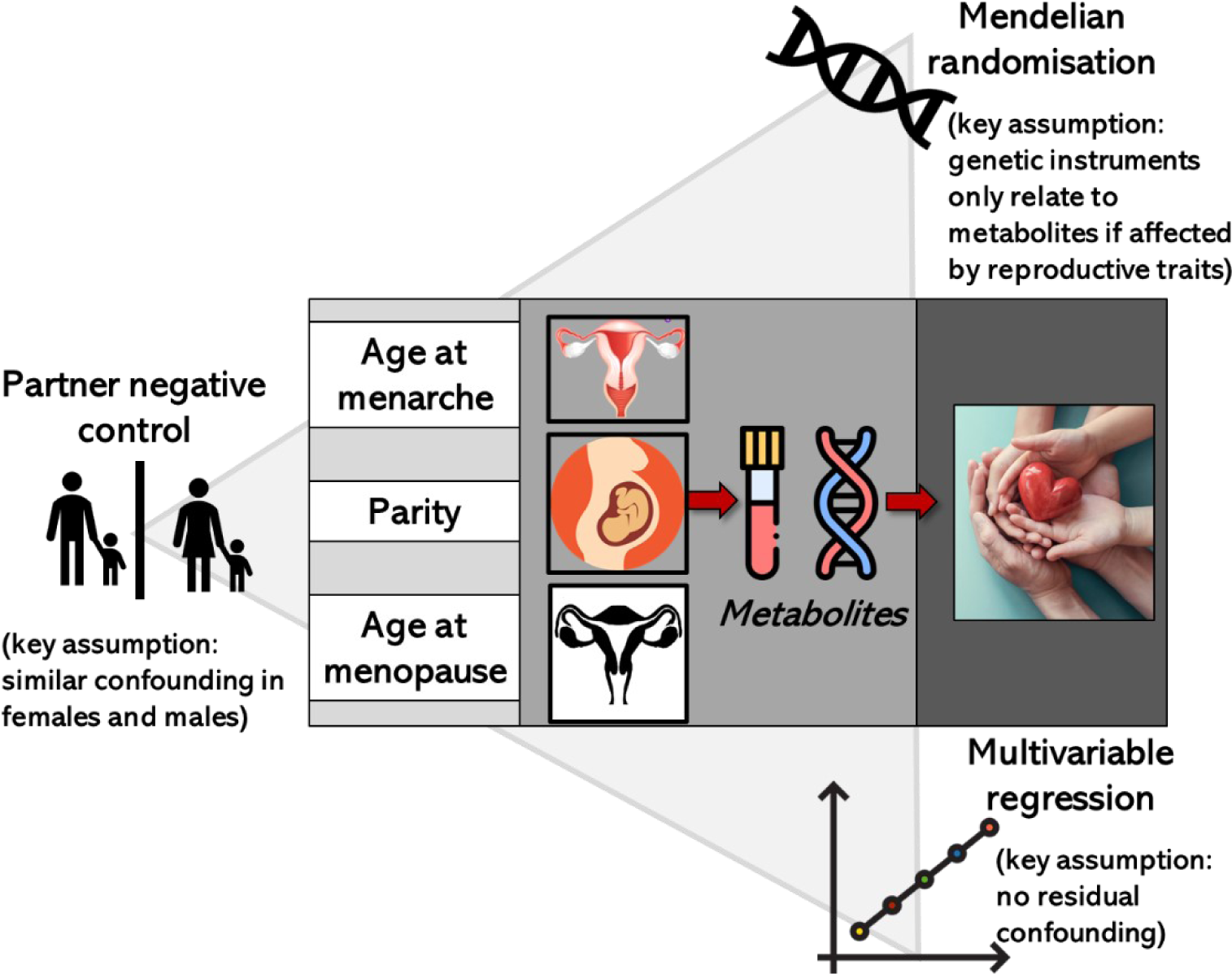
Overview of study design: key assumptions and biases Infographic summarising the different approaches taken to assess associations between reproductive traits and metabolites The figure illustrates key assumptions and sources of bias for each method (and differences across methods) in the context of our study. An exhaustive review of assumptions/biases for each method is outside the scope of this work. However, we acknowledge that there are other potential sources of biases that could affect findings such as selection bias related to the low response in UKB.

## Results

We used data from 65,699 UK Biobank female participants with 249 metabolic measures quantified by nuclear magnetic resonance (NMR). Self-reported age at menarche (in years), parity (in number of live born children) and age at menopause (in years) were reported at baseline when participants mean age was 56 years old (range: 37 to 73). NMR metabolites were measured on blood samples taken at baseline or first repeat assessment (more details in methods). The characteristics of these participants are shown in **Table 1** (and split by each of our reproductive markers (categorized) in **Supplementary Tables 1-3**). At recruitment (baseline) women were aged (mean) 56 (SD=8.0) years, 21% drank three or four times a week and 40% were previous/currents smokers. 81% of women had one or more live births whilst the mean age of menarche was 13 years (SD=1.3). 59% (37, 428) women reported they went through a natural menopause with a mean age of menopause of 49.7 years (SD=5.1). **Supplementary Table 4** shows the distribution of NMR metabolic measures among UK Biobank females. The proportion of women with missing data across metabolic measures ranged from 0.3% to 6.1%.

**Table 1.**
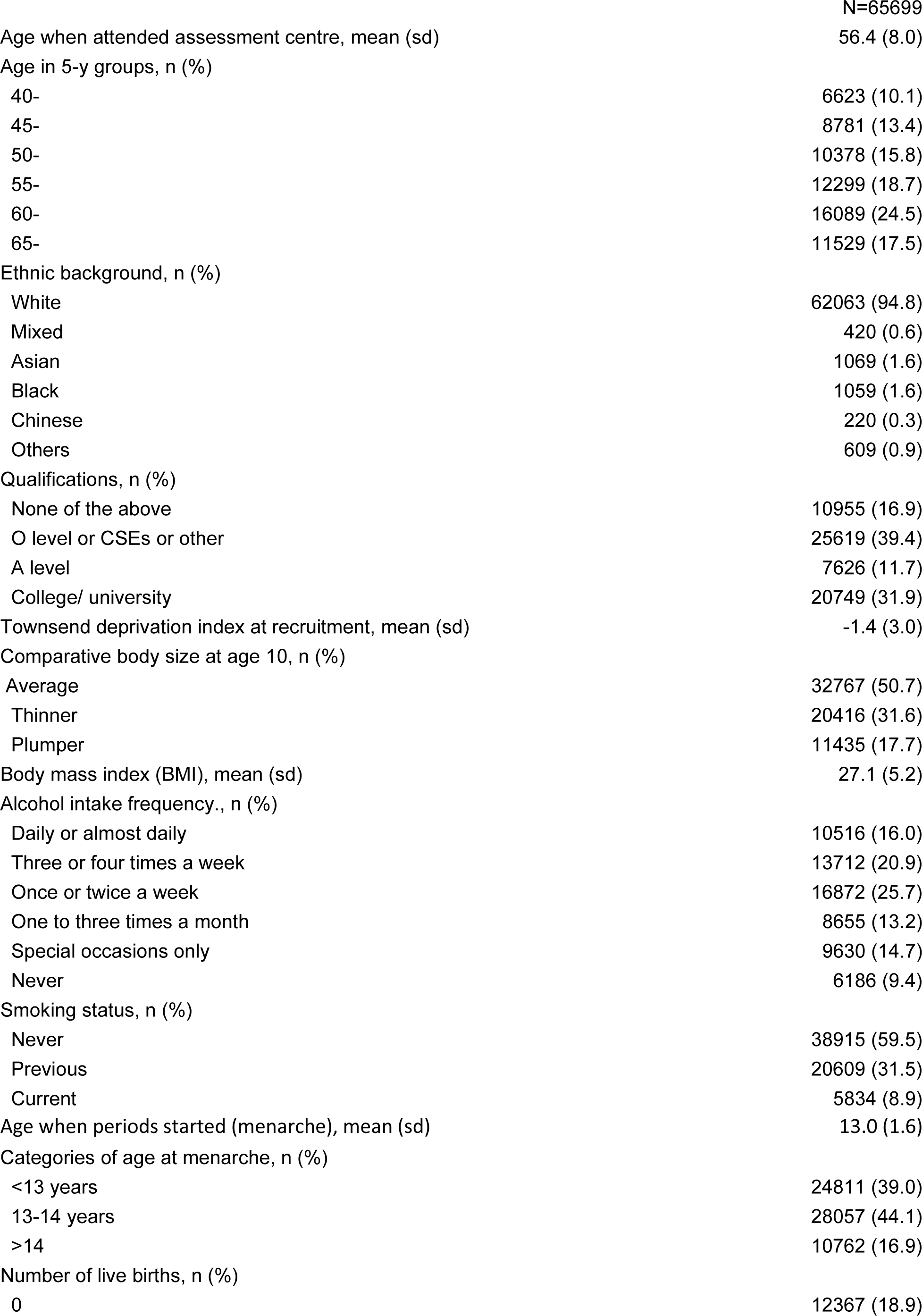
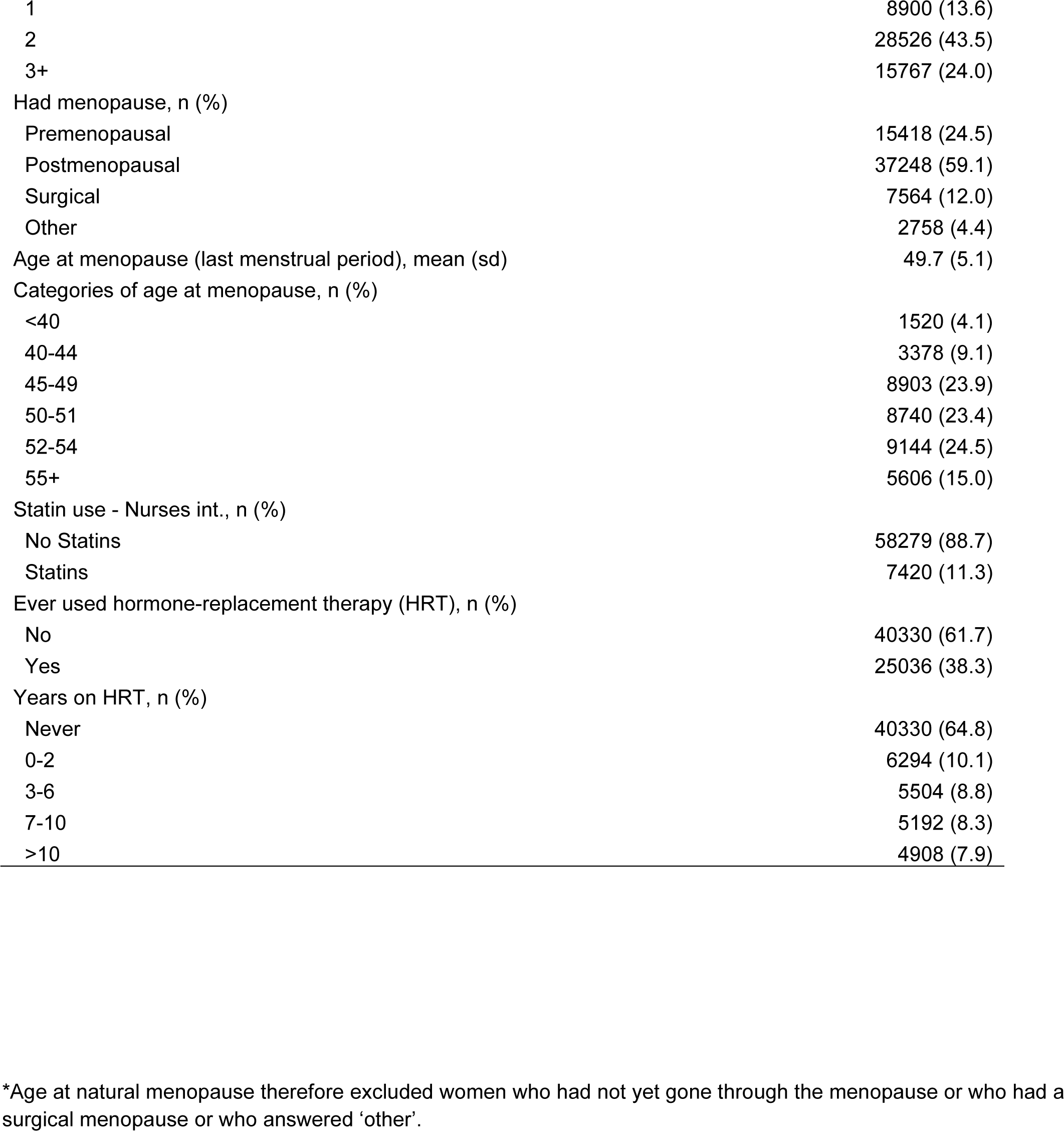
Distribution of characteristics of UK Biobank participants (females only) with NMR metabolomics data

|  |  |
| --- | --- |
|  | N=65699 |
| Age when attended assessment centre, mean (sd) | 56.4 (8.0) |
| Age in 5-y groups, n (%) |  |
| 40- | 6623 (10.1) |
| 45- | 8781 (13.4) |
| 50- | 10378 (15.8) |
| 55- | 12299 (18.7) |
| 60- | 16089 (24.5) |
| 65- | 11529 (17.5) |
| Ethnic background, n (%) |  |
| White | 62063 (94.8) |
| Mixed | 420 (0.6) |
| Asian | 1069 (1.6) |
| Black | 1059 (1.6) |
| Chinese | 220 (0.3) |
| Others | 609 (0.9) |
| Qualifications, n (%) |  |
| None of the above | 10955 (16.9) |
| O level or CSEs or other | 25619 (39.4) |
| A level | 7626 (11.7) |
| College/ university | 20749 (31.9) |
| Townsend deprivation index at recruitment, mean (sd) | -1.4 (3.0) |
| Comparative body size at age 10, n (%) |  |
| Average | 32767 (50.7) |
| Thinner | 20416 (31.6) |
| Plumper | 11435 (17.7) |
| Body mass index (BMI), mean (sd) | 27.1 (5.2) |
| Alcohol intake frequency., n (%) |  |
| Daily or almost daily | 10516 (16.0) |
| Three or four times a week | 13712 (20.9) |
| Once or twice a week | 16872 (25.7) |
| One to three times a month | 8655 (13.2) |
| Special occasions only | 9630 (14.7) |
| Never | 6186 (9.4) |
| Smoking status, n (%) |  |
| Never | 38915 (59.5) |
| Previous | 20609 (31.5) |
| Current | 5834 (8.9) |
| Age when periods started (menarche), mean (sd) | 13.0 (1.6) |
| Categories of age at menarche, n (%) |  |
| <13 years | 24811 (39.0) |
| 13-14 years | 28057 (44.1) |
| >14 | 10762 (16.9) |
| Number of live births, n (%) |  |
| 0 | 12367 (18.9) |
| 1 | 8900 (13.6) |
| 2 | 28526 (43.5) |
| 3+ | 15767 (24.0) |
| Had menopause, n (%) |  |
| Premenopausal | 15418 (24.5) |
| Postmenopausal | 37248 (59.1) |
| Surgical | 7564 (12.0) |
| Other | 2758 (4.4) |
| Age at menopause (last menstrual period), mean (sd) | 49.7 (5.1) |
| Categories of age at menopause, n (%) |  |
| <40 | 1520 (4.1) |
| 40-44 | 3378 (9.1) |
| 45-49 | 8903 (23.9) |
| 50-51 | 8740 (23.4) |
| 52-54 | 9144 (24.5) |
| 55+ | 5606 (15.0) |
| Statin use - Nurses int., n (%) |  |
| No Statins | 58279 (88.7) |
| Statins | 7420 (11.3) |
| Ever used hormone-replacement therapy (HRT), n (%) |  |
| No | 40330 (61.7) |
| Yes | 25036 (38.3) |
| Years on HRT, n (%) |  |
| Never | 40330 (64.8) |
| 0-2 | 6294 (10.1) |
| 3-6 | 5504 (8.8) |
| 7-10 | 5192 (8.3) |
| >10 | 4908 (7.9) |

\*Age at natural menopause therefore excluded women who had not yet gone through the menopause or who had a surgical menopause or who answered 'other'.

We used three approaches relying on different assumptions to explore the causal role of women’s reproductive markers on later life metabolic profile. For the first approach (‘multivariable regression’), we used linear regression models to estimate the association of reproductive markers with metabolic measures after adjusting for age at baseline, education and body composition at age 10. In sensitivity analyses, for the 55 non-derived metabolites, we categorised age at menarche, parity and age at natural menopause, tested for a linear trend and, where there was evidence of non-linearity, fit restricted cubic splines. For the second approach (‘negative control design’ – only applicable for parity), we used linear regression models to test whether number of live born children in men was associated with their metabolic measures; given men do not experience the repeated physiological stress of pregnancy, but are likely to demonstrate the same associations of confounders (eg. socioeconomic position, BMI, smoking) with number of live births, similar associations of number of live births with metabolic measures between men and women would indicate bias (e.g. due to confounding) rather than a causal effect of being exposed to the physiological stress of pregnancy on women’s metabolic profile. For the third approach (‘MR’), we selected single nucleotide polymorphisms (SNPs) as genetic instruments for each reproductive marker from previous genome-wide association studies (GWAS) and performed two-sample MR to estimate the effect of reproductive markers on metabolic measures using the standard inverse variance weighted (IVW) method. For both multivariable and MR analyses, we adopted P-value < 0.00093, which accounts for the approximate number of independent tests as detailed in ‘Statistical analyses’.

### Age at menarche

In the main multivariable regression analyses (adjusting for age at baseline, education and body composition at age 10), older age at menarche was associated with higher concentration of glutamine, glycine, albumin, apolipoprotein A1, cholines, phosphatidylcholines, and sphingomyelins but lower concentration of alanine, branched-chain amino acids (isoleucine, leucine and valine), aromatic amino acids (phenylalanine and tyrosine), fatty acids (monounsaturated fatty acids (MUFA), omega-3 polyunsaturated fatty acids (PUFA), and saturated fatty acids (SFA)), glycolysis-related metabolites (glucose, lactate, pyruvate), acetoacetate, and glycoprotein acetyls (GlycA) (P < 0.00093) (**Figure 2A** and **Supplementary Table 5**). Older age at menarche was also associated with numerous lipoprotein-related traits at P < 0.00093, particularly with higher number of particles, size, and lipid content in high-density lipoproteins (HDL) and lower number of particles, size, and lipid content in very low-density lipoproteins (VLDL) (**Figure 2A**). The associations of age at menarche with HDL-related traits were mostly due to larger HDL subclasses (i.e. medium, large and very large particles), while associations with VLDL-related traits were observed across VLDL subclasses (**Supplementary Figure 1** and **Supplementary Table 5**). In sensitivity analyses with further adjustments for BMI, smoking and alcohol status at baseline, findings for an association of older age at menarche were largely or completely attenuated towards the null for most metabolic measures with few exceptions, such as glutamine, glycine, omega-3 PUFA, pyruvate, lactate, and acetoacetate (**Supplementary Figure 2**). There was evidence of non-linearity between categories of age at menarche (<13, 13-14, >14 years) and 17 metabolites (**Supplementary Table 6** and **Supplementary Figure 3**). Restricted cubic spline models (with 3 knots at ages 11, 13, and 15 years) generally showed an increase in albumin, apolipoprotein A1, cholines, DHA, LA, and phosphatidylcholines with older age at menarche until approximately age 13, in line with our linear association, and then began to flatten and/or decrease (**Supplementary Table 7** and **Supplementary Figure 4**). Whilst older age at menarche was related to an increase in GlycA until ∼13 years and then began to flatten.

**Figure 2A.**
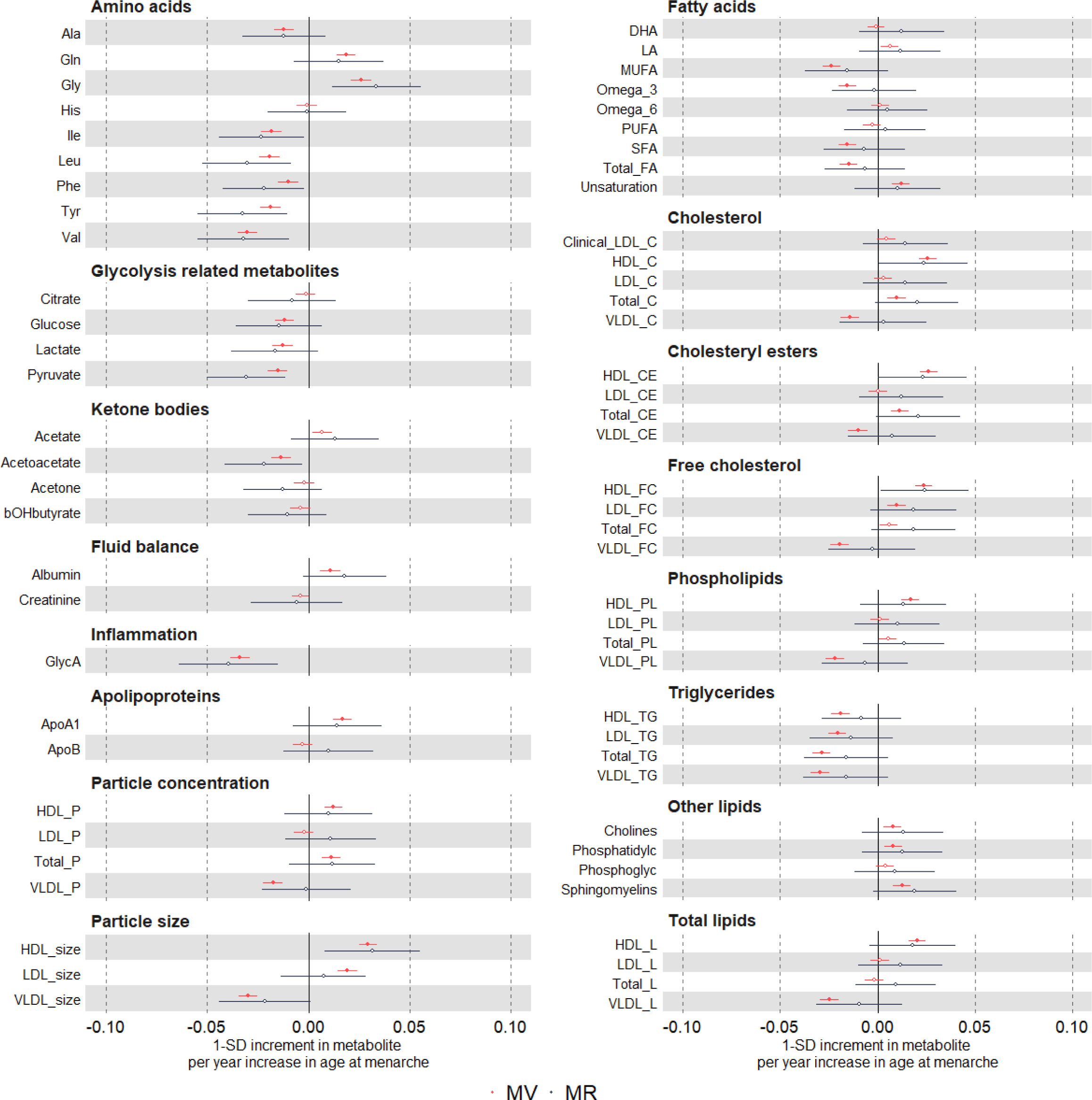
Multivariable regression and Mendelian randomization estimates for the associations between older age at menarche and metabolic measures. Results are presented as standard unit changes in metabolic measure per 1 year increase in age at menarche. Circles denote point estimates and indicate p-value < 0.00093 (closed circle) or ≥ 0.00093 (open circles). Horizontal bars denote 95% confidence intervals. Multivariable regression models were adjusted for age at recruitment, body size at age 10 and education. Mendelian randomization models were estimated using the inverse variance weighted method. Abbreviations for Figures 2A-C and 4 are given in ‘Abbreviations’ at the end of the manuscript.

**Figure 2B.**
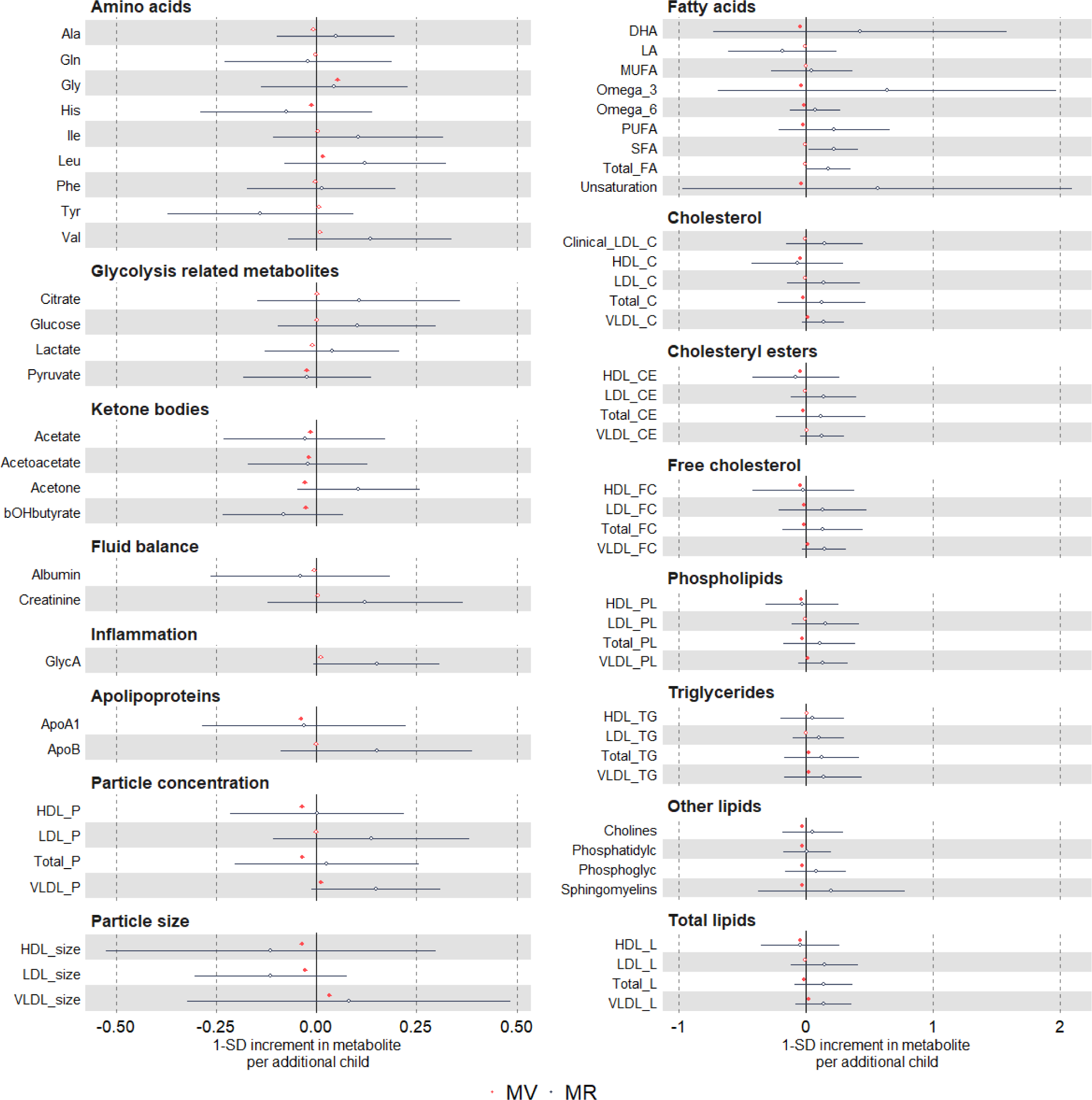
Multivariable regression and Mendelian randomization estimates for the associations between higher parity and metabolic measures. Mendelian randomization estimates for the associations between reproductive traits (i.e. older age at menarche, higher parity, and older age at natural menopause) and metabolic measures Results are presented as standard unit changes in metabolic measure per 1 additional birth. Circles denote point estimates and indicate p-value < 0.00093 (closed circle) or ≥ 0.00093 (open circles). Horizontal bars denote 95% confidence intervals. Multivariable regression models were adjusted for age at recruitment, body size at age 10 and education. Mendelian randomization models were estimated using the inverse variance weighted method. Abbreviations for Figures 2A-C and 4 are given in ‘Abbreviations’ at the end of the manuscript.

**Figure 2C.**
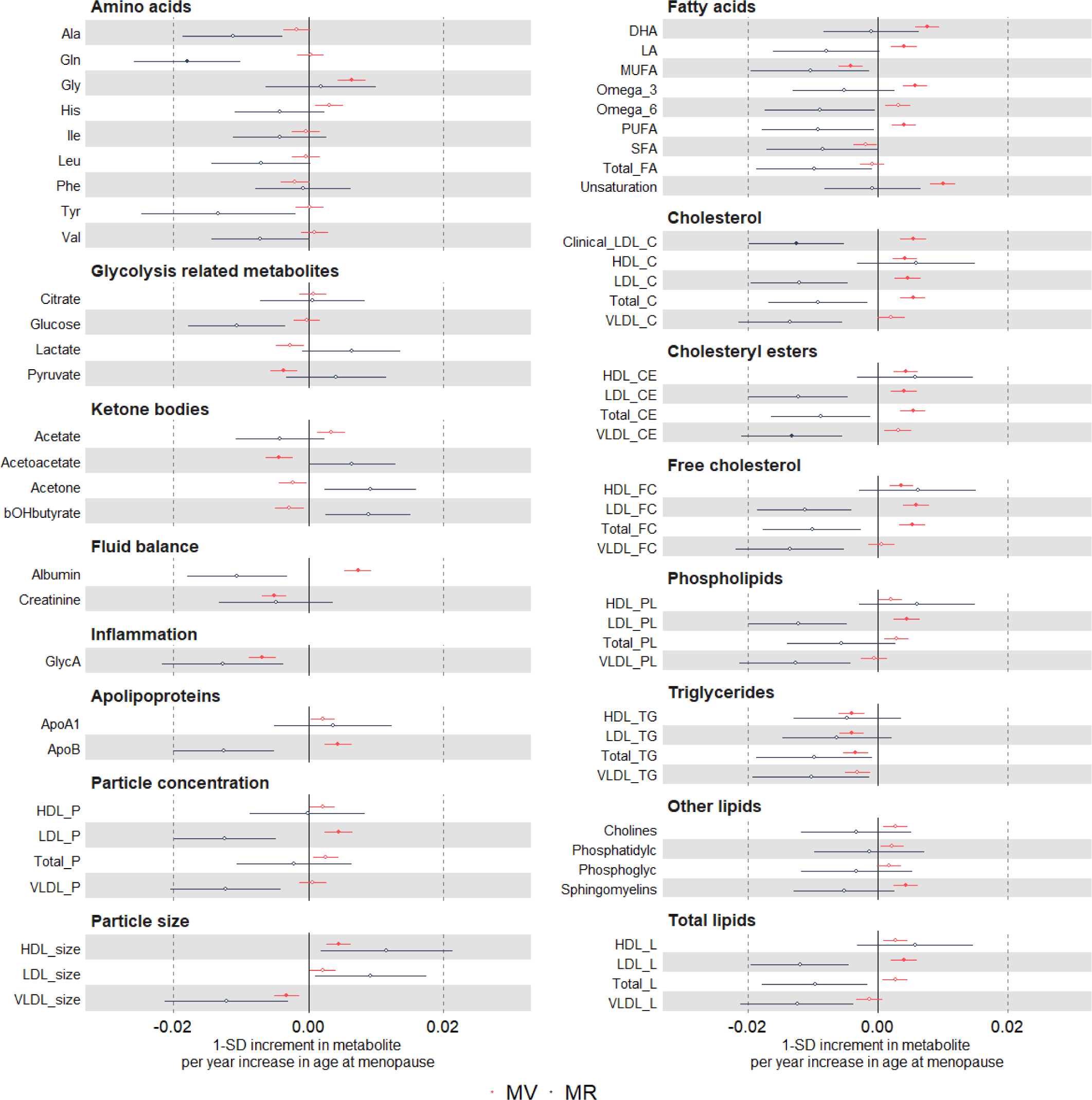
Multivariable regression and Mendelian randomization estimates for the associations between older age at natural menopause and metabolic measures. Results are presented as standard unit changes in metabolic measure per 1 year increase in age at natural menopause. Circles denote point estimates and indicate p-value < 0.00093 (closed circle) or ≥ 0.00093 (open circles). Horizontal bars denote 95% confidence intervals. Multivariable regression models were adjusted for age at recruitment, body size at age 10 and education. Mendelian randomization models were estimated using the inverse variance weighted method. Abbreviations for Figures 2A-C and 4 are given in ‘Abbreviations’ at the end of the manuscript.

For the MR analyses, we selected 389 SNPs as instruments for age at menarche, which explained 7.4% of its phenotypic variance with a corresponding mean F statistic of 63 (**Supplementary Table 8**). Overall, MR estimates using IVW were in agreement with multivariable regression estimates in direction and magnitude (**Figures 2A** and **Supplementary Figure 1**); however, due to the higher degree of uncertainty for IVW estimates, no result passed our threshold for multiple testing correction (P < 0.00093). Following reviewer’s comments, we repeated the IVW analyses for a larger sample of women (N=216,514-241,244) for the eight biomarkers assayed using clinical chemistry techniques that matched measures in the NMR metabolomics platform — i.e. albumin, apolipoprotein A1, apolipoprotein B, glucose, HDL-cholesterol, LDL-cholesterol, total cholesterol, and triglycerides. These results provided further evidence of older age at menarche being related to higher albumin, apolipoprotein A1, HDL-cholesterol, and lower triglycerides (P < 0.00093) (**Figure 3**). Given the a priori evidence of bidirectional effects between age at menarche and BMI, we also performed multivariable IVW accounting for adult BMI to estimate the direct effects of age at menarche on metabolic measures, which resulted in estimates partly or completely attenuated to the null for most metabolic measures with few exceptions, such as glutamine and glycine (**Supplementary Figure 5** and **6**).

**Figure 3.**
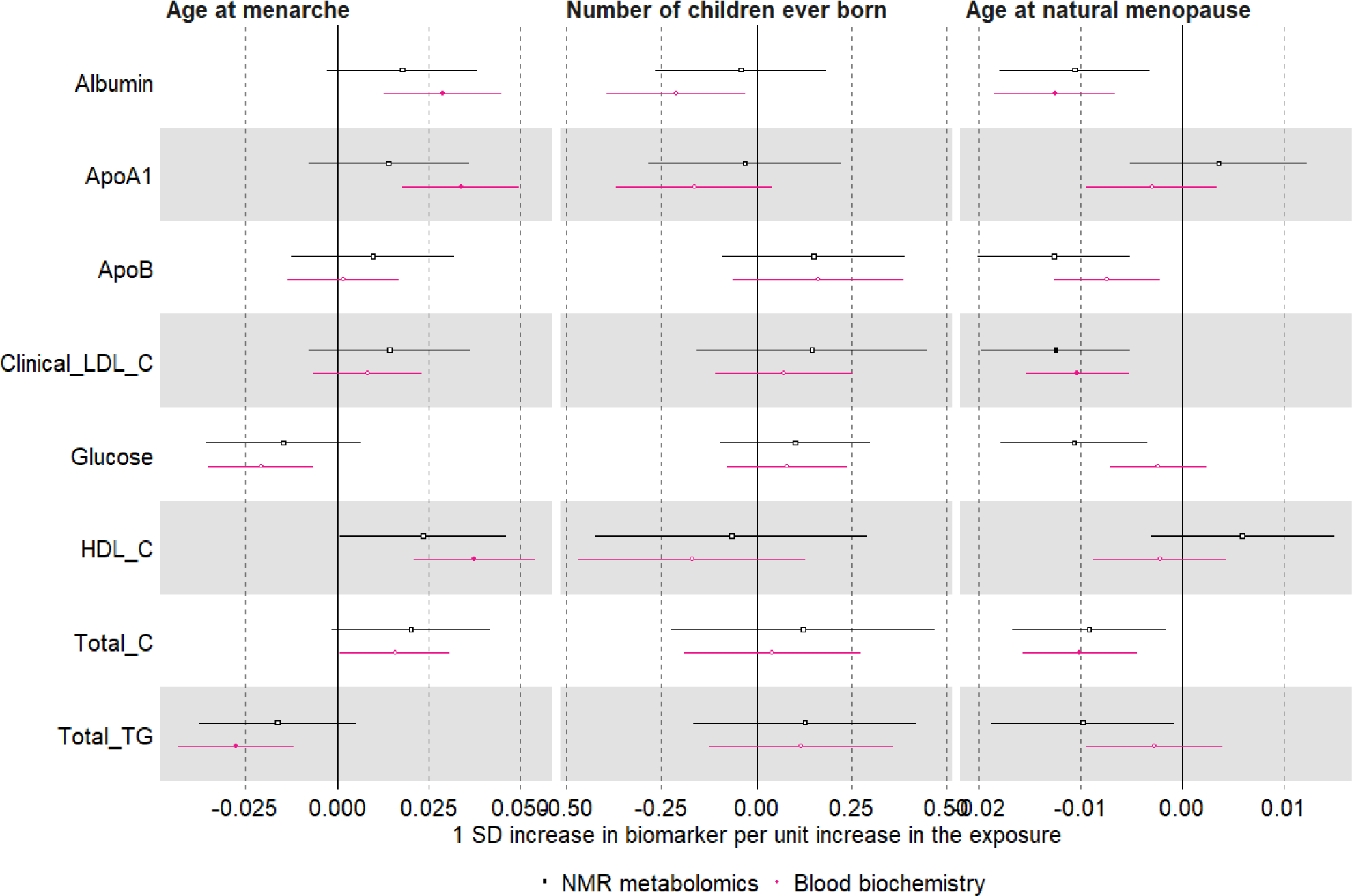
Mendelian randomization estimates for the relation between older age at menarche and metabolic measures among females measured using NMR metabolomics (black) or clinical chemistry methods (pink)

### Parity

In the main multivariable regression analyses (adjusting for age at baseline, education and body composition at age 10), higher parity was related to higher concentrations of glycine and leucine, but lower concentrations of histidine, fatty acids (docosahexaenoic acid (DHA), Omega 3, Omega 6 PUFA), pyruvate, ketone bodies (acetate, acetoacetate, acetone and β-hydroxybutyrate), and apolipoprotein A1 (P < 0.00093) (**Figures 2B** and **Supplementary Table 9**). Higher parity was also associated with numerous lipoprotein-related measures at P < 0.00093, particularly with lower and higher number of particles, size, and lipid content for HDL and VLDL, respectively, as well as lower size of LDL particles (**Figure 2B**). The associations of parity with lipoprotein-related measures were observed across most VLDL and HDL subclasses, whereas associations with LDL-related measures were mostly driven by larger LDL particles (**Supplementary Figure 7** and **Supplementary Table 9**). In sensitivity analyses with further adjustments for BMI, smoking and alcohol status at baseline, higher parity associations were consistent for glycine, histidine, fatty acids, pyruvate, ketone bodies, apolipoprotein A1, and partly attenuated towards the null for VLDL- and HDL-related traits (**Supplementary Figure 8**). There was some evidence of non-linearity between parity (0,1,2,3+) and 28 metabolites (**Supplementary Table 6** and **Supplementary Figure 9**). However, restricted cubic spline models (with knots at 1, 2, and 3) generally showed monotonic relationships for those with no to four pregnancies, consistent with the main analysis models (**Supplementary Table 10** and **Supplementary Figure 10**).

We used males as a negative control since men cannot experience the effects of being exposed to the stress test of pregnancy. Therefore, similar results between men and women would be indicative of bias, such as due to confounding by sociodemographic (e.g. education attainment) and biological (e.g. infertility) factors, rather than by an effect of repeated exposure to pregnancy. When using number of children in males as a negative control, we observed that associations for leucine, histidine, pyruvate, and ketone bodies were similar between men and women (i.e. directionally consistent, similar effect estimates and 95% confidence intervals overlapped between male and female estimates). On the other hand, association estimates for fatty acids, apolipoprotein A1, and lipoprotein-related traits were weaker or consistent with the null, and glycine was in opposite direction, in males compared to females (**Figure 4**). For the MR analyses, we selected 32 SNPs as instruments for parity, which explained 0.2% of its phenotypic variance with a corresponding mean F statistic of 31 (**Supplementary Table 8**). It is unclear whether estimates from multivariable regression and MR analyses are consistent with each other due to the high level of uncertainty in the latter (**Figures 2B** and **Supplementary Figure 7**), which persisted even when using the larger sample of women with selected biomarkers assayed by clinical chemistry (**Figure 3**).

**Figure 4.**
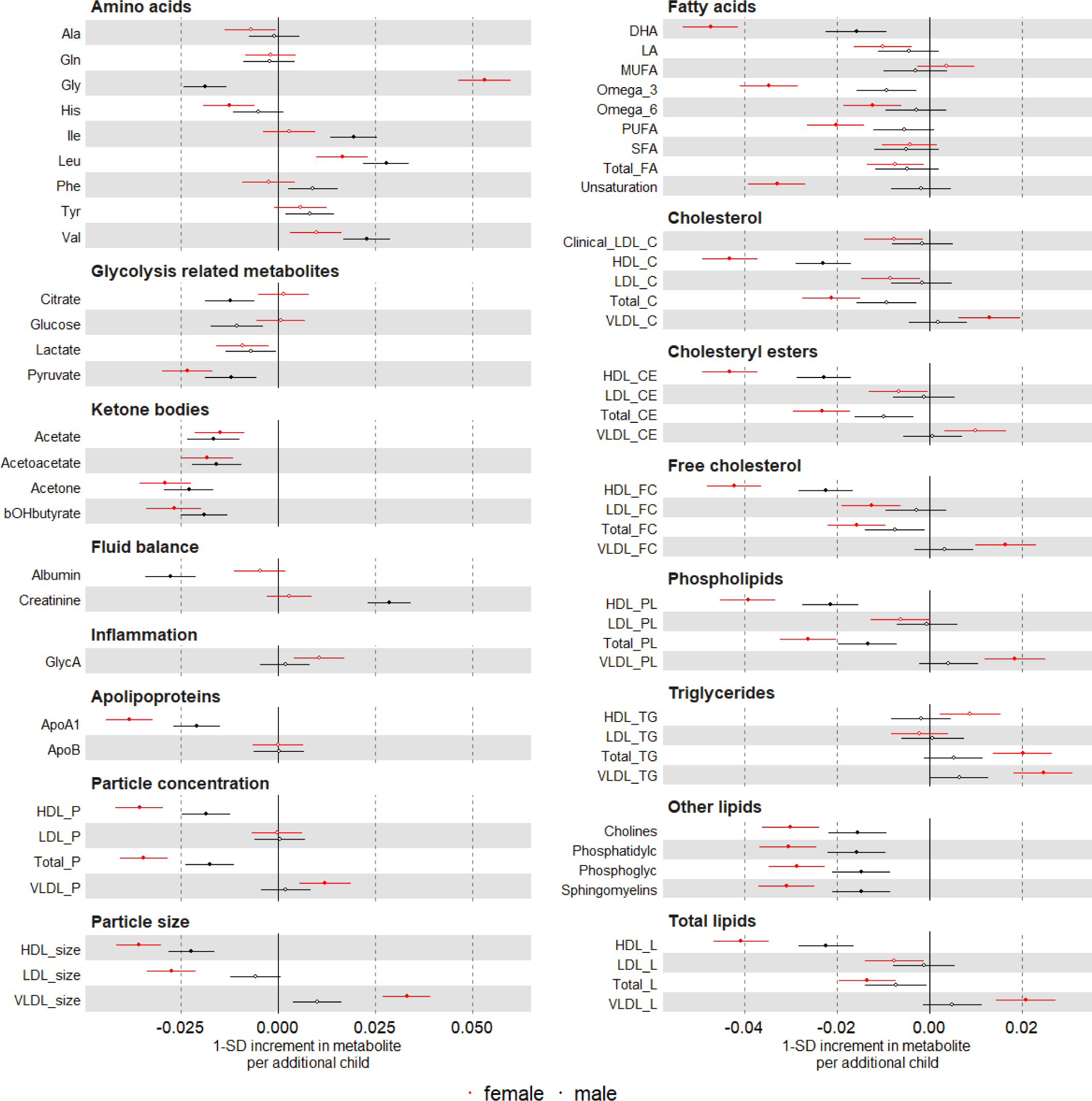
Multivariable regression estimates for the associations of parity (women) or number of children (men) with metabolic measures: negative control analyses Models adjusted for age at baseline, education, and body composition at age 10.

### Age at natural menopause

In the main multivariable regression analyses (adjusting for age at baseline, education and body composition at age 10), older age at menopause was related to higher glycine, PUFA (e.g. DHA and LA), albumin, apolipoprotein B and sphingomyelins, but lower concentration of MUFA, pyruvate, acetoacetate, creatinine and GlycA (P < 0.00093) (**Figures 2C** and **Supplementary Table 11**). Older age at menopause was also associated with numerous lipoprotein-related traits at P < 0.00093, particularly with higher number of particles and lipid content in LDL, larger size of HDL particles, and lower size of VLDL particles (**Figure 2C**). The associations between age at menopause and LDL-related traits were observed across LDL subclasses (i.e. from small to large), whereas associations with HDL-related traits were mostly driven by larger HDL particles (**Supplementary Figure 11**). In sensitivity analyses with further adjustments for BMI, smoking and alcohol status at baseline, associations between older age at natural menopause and metabolites remained similar, except for associations with HDL-related traits which were partly attenuated (**Supplementary Figure 12**). There was evidence of non-linearity across 24 metabolites (**Supplementary Table 6** and **Supplementary Figure 13**) in the multivariable regression when menopause was categorised (<49, 49-50, 51-53, >53 years). Restricted cubic spline models (with 4 knots) were generally consistent with the main analysis (assuming a linear association) until age at menopause ∼55 years when most metabolites decreased (**Supplementary Table 12** and **Supplementary Figure 14**).

For the MR analyses, we selected 290 SNPs as instruments for age at natural menopause, which explained 8.2% of its phenotypic variance with a corresponding mean F statistic of 141 (**Supplementary Table 8**). Estimates from multivariable regression and MR analyses were inconsistent in direction for many metabolic measures (**Figure 2C**). In particular, in contrast to results from multivariable regression, MR analyses indicated older age at menopause to be related to lower concentration of fatty acids (e.g. LA), albumin, apolipoprotein B, as well as lower number of particles, lipid content and size of LDL across subtypes (from small to large) (**Figure 2C** and **Supplementary Figure 11**). For some metabolites, such as GlycA and HDL-related traits, results were consistent in direction between multivariable regression and MR. For alanine, glutamine and glucose, MR analysis suggested older age of menopause to be related to lower circulating metabolite levels, which had not been observed in multivariable regression analysis (**Figures 2C** and **Supplementary Table 11**). As expected, there was more uncertainty in MR estimates and only results for glutamine and some LDL- and VLDL-related measures passed the threshold for multiple test correction (P < 0.00093). Repeating the MR analyses in the larger sample of women (N=216,514-241,244) with selected biomarkers assayed by clinical chemistry confirmed that older age at natural menopause was related to lower albumin, LDL-cholesterol, and total cholesterol at P < 0.00093 (**Figure 3**).

We performed further analyses to investigate reasons underlying discrepant findings between multivariable and MR estimates for some metabolic measures. These analyses were restricted to the eight clinical chemistry biomarkers matching measures in the NMR platform to maximise statistical power since they have been measured in the full UK Biobank sample. First, we hypothesised that discrepant findings were related to differences in the sample used for multivariable regression, which excludes women with missing data on age at menopause (hereafter ‘selected sample’), and two-sample MR, which includes women even if they are missing data on age at natural menopause (hereafter ‘full sample’). To test that, we compared estimates from multivariable regression on the selected sample to MR on both the selected sample and full sample. In agreement with our hypothesis, multivariable regression and MR estimates for LDL-cholesterol and related traits (i.e. apolipoprotein B and total cholesterol) are comparable when restricting to the selected sample. In contrast, for albumin, discrepant results were related to differences between multivariable regression and MR rather than between selected and full sample (**Supplementary Figure 15**). Second, given women with missing data at age at menopause are typically pre-menopausal and younger, we explored age-stratified multivariable and MR estimates, which revealed a strong effect modification by chronological age on the association of age at menopause with LDL-c and related traits – e.g. older age at menopause is related to substantially lower LDL-cholesterol in younger women (≤ 50 y) (e.g. MV: -0.018 SD, 95%CI: -0.021, -0.015), but slightly higher LDL-cholesterol in older women (> 63 y) (e.g. MV: 0.004 SD, 95%CI: 0.003, 0.006) (**Supplementary Figure 15**). Differences related to chronological age at baseline were also observed for other biomarkers, such as albumin. This age patterned results were largely similar when excluding women using statins at baseline or with a history of using hormone replacement therapy (HRT) (**Supplementary Figure 16**).

### Exploring the plausibility of MR assumptions

We conducted a series of sensitivity analyses to explore the plausibility of key MR assumptions, required for the method to provide a valid test of the presence of a causal effect.

First, we tested whether MR findings are likely to be biased by population stratification, assortative mating and indirect genetic effects of parents using two approaches: (i) performing two-sample MR analyses using (sex-combined) data from a recent within-siblings GWAS, and (ii) conducting two-sample MR on negative control outcomes (i.e. skin colour and skin tanning ability). Two-sample MR estimates for the effect of genetic susceptibility for older age at menarche, parity, and age at natural menopause on five available biomarkers was broadly consistent when estimated among unrelated individuals or between siblings. Results for age at menarche were slightly overestimated for triglycerides and underestimated for glycated haemoglobin in unrelated individuals, while results for a positive relation between age at natural menopause and HDL-cholesterol was supported by analyses between siblings but not among unrelated individuals (**Supplementary Figure 17**). We did not observe an association of genetically-predicted reproductive markers with skin colour or tanning (**Supplementary Table 13**). Taken together, these sensitivity analyses indicate that our main MR estimates are unlikely to be substantially biased by population stratification, assortative mating and indirect genetic effects of parents.

Second, we explored the presence of bias due to pleiotropic variants by using MR methods other than IVW: the weighted median estimator and MR-Egger. These methods can provide valid tests for the presence of a causal effect under different (and weaker) assumptions about the nature of the underlying horizontal pleiotropy compared to IVW. Estimates from IVW and weighted median were consistent in direction for most relationships between reproductive markers and metabolic measures. In most instances, estimates from MR-Egger method were uninformative given the high degree of uncertainty (**Supplementary Figures 18, 19, 20**).

Third, we assessed potential bias due to sample overlap from including UK Biobank individuals in genetic association estimates for both exposures and outcomes. This was achieved by using data from previous GWAS that did not include UK Biobank, available for age at menarche and age at natural menopause, to select SNPs (and genetic associations estimates with exposures) for two-sample MR analyses (**Supplementary Table 14**). When using SNPs selected from previous GWASes that did not include UK Biobank participants, results for of age at menarche and age at natural menopause were largely consistent, although less precise, compared to estimates from the main analyses using data with overlapping samples (**Supplementary Figure 21** and **22**).

## Discussion

Our findings indicate that reproductive markers across women’s lifespan are associated with distinct metabolic signatures in later life. Age at menarche, parity and age at natural menopause were related to numerous metabolic measures, representing multiple dimensions of metabolism, including amino acids, fatty acids, glucose, ketone bodies, and lipoprotein metabolism (see **Summary box** for key findings).

### Age at menarche

Age at menarche is frequently used as a proxy of puberty onset among females in epidemiological studies ^2,28^. Our findings for the relation of age at menarche with metabolic measures were broadly concordant between multivariable regression and MR analyses, and were supportive of older puberty onset being related to a less atherogenic metabolic profile among adult women.

Both multivariable regression and MR estimates were markedly attenuated when accounting for adult BMI for most metabolic measures with few exceptions (e.g. glutamine and glycine), suggesting that the effect of age at menarche on adult metabolites are largely explained by adult BMI. There is evidence of a bi-directional relationship between puberty timing and adiposity, where pre-pubertal adiposity influences puberty timing, which in turn influences post-pubertal adiposity ^13,28,29^. In addition, genetic variants influencing age at menarche are known to influence BMI before and after puberty ^28,29^.

The complex relationship between puberty timing and adiposity complicates inferences of the effect of age at menarche on the metabolic profile or disease risk in adulthood since the observed associations could reflect adult BMI mediating the effect of early age at menarche on metabolic measures or a confounding path from pre-puberty BMI. A previous one-sample MR study ^28^ investigating the effect of age at menarche on NMR metabolic measures reported that results were largely attenuated when accounting for BMI at 8 years old, which suggests that the estimated effect of age at menarche on the metabolic profile is largely confounded by pre-pubertal adiposity, though larger MR studies with repeat BMI and metabolic profiles before and after menarche are needed to rule out a potential causal mediated effect. In our study, accounting for self-reported adiposity in childhood in multivariable regression models did not substantially change effect estimates. This discrepancy might be related to residual confounding in our study (e.g. due to higher measurement error in our measure of childhood adiposity) or different age distributions between ours (mean=55 years) and this previous study (mean=18 years).

### Parity

Pregnant women undergo marked changes in physiology (e.g. lipid/glucose metabolism, adiposity, vascular function, hormone levels, and inflammatory response) and lifestyle (e.g. diet and physical activity ^30^), most of which return to their pre-pregnancy state after delivery ^20,30^. However, there are concerns that some of these changes might persist and accumulate over multiple pregnancies, impacting women’s cardiovascular health in the future, or that pregnancy acts as a stress test, unmasking an underlying high risk for cardiovascular disease ^30,31^. We used parity as a marker of being exposed to the physiological stress of multiple pregnancies.

In multivariable regression analyses, we found that higher parity, proxied by number of children ever born, was associated with both favorable (e.g. less LDL particles) and unfavorable (e.g. higher number of particles and lipid content in VLDL) changes in the metabolic profile. Evidence from MR analyses is uncertain due to the high imprecision in effect estimates. Using males as a negative control, we showed that the associations between number of children ever born and metabolic measures among men were largely null for lipoprotein-related measures or in opposite direction for glycine compared to females. This inconsistency between female and male findings reinforces that the metabolic signature associated with parity among females largely reflects a causal effect of parity on the metabolome rather than spurious results due to confounding or selection bias (assuming confounding structures and selection mechanisms are similar between men and women). A possible mechanism is that higher parity leads to greater insulin resistance in pregnant women and subsequently increases the production and secretion of hepatic triglycerides, which can lead to an increased lipid content in VLDL particles.

In line with our findings, other studies have reported that higher parity is related to higher cardiovascular disease risk in women ^32^. Negative control analyses (comparing associations of number of children in women and men) have been conducted previously in two UK cohorts, with one suggesting that associations with lipids and body composition in women may be due to confounding (as associations are similar in women and men) ^33,34^ and the second, the largest of these studies to date, and the only one to look at disease end points, finding evidence of a stronger association for risk factors and coronary heart disease in women than men suggesting parity itself has some influence on cardiovascular disease risk ^5^, Furthermore, studies in women only that are able to control for pre-pregnancy measures, suggest pregnancy and parity have a potentially lasting effect on adverse lipid profiles ^35,36^.

### Age at natural menopause

Previous conventional observational studies suggested older age at natural menopause to be associated with lower risk of cardiometabolic diseases ^37^ over and above the underlying age trajectory ^21,38^. In our study, estimates for VLDL- and LDL-related traits were inconsistent between multivariable regressions and MR; the former suggests older age at menopause to be related to a more atherogenic profile, while the latter indicates the opposite. On the other hand, both multivariable regression and MR estimates suggested older age at menopause to be related to higher lipid content in HDL particles and lower systemic inflammation, as proxied by GlycA. Consistent with Auro et al ^39^, a recent multivariable analysis of 218 Finnish women going through the menopause transition found that menopause was similarly associated with a higher lipid content in HDL particles and lower systemic inflammation ^40^. Whilst a recent longitudinal study of up to 3892 women with up to 12 CVD risk factors measured as they went through the menopausal transitional also found higher HDL and non-HDL associations, stronger effects of chronological rather than reproductive aging were observed ^41^.

Findings from our multivariable regression analyses for age at natural menopause should be interpreted with caution given 40% of women were excluded from these analyses as they had not experienced a natural menopause and 7% of the women had experienced menopause less than two years before study recruitment (when blood samples for NMR metabolomics were collected). In follow-up analyses, we have shown that discrepancy in findings between multivariable and MR for LDL-related traits were related to the exclusion of younger pre-menopausal women in multivariable regression. In addition, age-stratified analyses revealed that age at menopause is related to lower LDL-cholesterol in younger women but slightly higher LDL-cholesterol in older women. These results did not seem to be explained by higher intake of statins or HRT among older women, although such analyses should be interpreted with caution given the potential for collider stratification bias. Previous longitudinal studies indicated that LDL-cholesterol ^41^ and related traits increase sharply through the menopause transition and early postmenopausal years and then plateau with increasing postmenopausal years ^42^. In our cross-sectional analyses, we observed a non-linear pattern for several metabolites, such that mean metabolite levels increase linearly with age at menopause until 50-55 years old and then decline. Taken together, we speculate that these findings explain the pattern by chronological age in the association between timing of menopause and LDL-related traits. However, larger longitudinal studies with longer follow-up are needed to tease apart the complex nature, and possible time-varying, effect of reproductive aging on the metabolome.

The largest two-sample MR analysis to date indicate that older age at menopause is related to lower risk of type 2 diabetes in females, but no difference in risk of cardiovascular disease or dyslipidemia (data combining males and females) ^43^. This is in agreement with our analyses suggesting older age at natural menopause is related to lower glucose, and with evidence from randomized controlled trials of estrogen therapy pointing to a protective effect on type 2 diabetes but no change in risk of cardiovascular diseases ^44-46^. The mechanisms underlying the putative protective effect of older menopause on the risk of metabolic diseases in MR studies is unclear, but might reflect an effect of prolonged exposure to sex hormones or of slower cell aging, given genetic variants associated with age at natural menopause are highly enriched for genes in DNA damage response pathways ^43,47^. The consistent results between MR of age at menopause and randomized controlled trials of estrogen therapy for type 2 diabetes indicates that prolonged exposure to sex hormones is likely to be involved. Moreover, the lipid metabolism is regulated by estrogen, meaning that lower levels of estrogen during menopause can cause an increase in lipids, particularly LDL, HDL, and triglycerides.

### Strengths and limitations

To the best of our knowledge, this is the largest study to examine the long-term impact of key events in reproductive life on the multiple metabolic measures in women. The use of large-scale metabolomics data and the integration of multiple analytical approaches are key strengths of our study as these allowed us to strengthen the inference of the causal impact of these reproductive markers on the metabolic health of females.

It is important to note that the validity of our findings rely on the plausibility of the assumptions underlying each analytical approach. For multivariable regression, we cannot exclude the possibility of bias due to residual confounding, especially given we were unable to adjust for key confounders in multivariable regression as measures of these were not available at or before the exposure to reproductive factors. For the use of negative controls, we rely on the unverifiable assumption that residual confounding and selection bias is similar in females and males analyses. It is plausible that factors relating to metabolites, such as age, ethnicity, socioeconomic position, and BMI, relate similarly to number of children in females and males and hence that confounding structures are similar. For MR, we have conducted extensive sensitivity analyses supporting the validity of our results; however, we cannot rule out the possibility of bias due to violations of the core instrumental variable assumptions. In addition, MR analyses for parity was uninformative given the low proportion of phenotypic variance explained by the genetic instruments.

When assessing non-linearity, our multivariable regression results were generally consistent between the main analysis model (assuming a linear relationship) and categories for most metabolites. For metabolites that showed evidence of non-linearity, many seemed to plateau and decrease with older ages of menarche and menopause and similarly with higher parity (however, this was also where we had the least amount of data, which could be driving some of the non-linearity). We were unable to fit non-linear associations in an MR framework given this would require much larger sample sizes; future studies with larger sample sizes should be better powered to examine potential non-linear effects using MR and contrast those found in the multivariable regression.

Whilst some key sources of bias may remain in each method, a key strength of our study is exploring and focusing on results that are consistent across the different methods. As the sources of bias differ between the methods causal inference is strengthened where there is consistency, as we see for example in associations between multivariable regression and MR for age at menarche, multivariable regression and negative control analyses for parity and multivariable regression and MR for age at natural menopause in relation to HDL-related measures and GlycA (but not LDL-related and other measures).

Across all analytical approaches, we cannot discard the presence of selection biases from using UK Biobank data given the low recruitment rate of the study (5%) and inclusion of healthier/wealthier individuals compared to the general UK population ^48^. In addition, the metabolic traits measured by the NMR metabolomics platform cover a limited set of metabolic pathways^49^, and, therefore, future studies including data from more sensitive metabolomics techniques, such as mass spectrometry, may improve coverage of the metabolome and provide insights into additional biological processes related to reproductive events. Triangulating results across different methods is useful for causal inference and where there are discrepant results it is important to explore these. We have found that the discrepant results between MR and multivariable regression for the association of age at menopause with some of the metabolites (notably LDL-c and related metabolites) are due to the exclusion of women with missing data on age at menopause in multivariable regression and a potential effect modification by chronological age in the association between age at menopause and some metabolic measures. However, we acknowledge we lack power to fully explore the mechanisms for this given the current number of UK Biobank participants with NMR data.

## Conclusions

Overall, older age at menarche/menopause were related to a more favorable metabolic profile, while a mixed pattern was observed for higher parity. Evidence supporting a relation between later pubertal timing and a less atherogenic metabolic profile was largely explained by adult BMI, while findings supporting a relation between slower reproductive aging and a less atherogenic metabolic profile was mostly observed among younger women. These results could contribute to identifying novel markers for the prevention of adverse cardiometabolic outcomes in women and/or methods for accurate risk prediction.

## Methods

### Study participants

UK Biobank is a population-based cohort consisting of approximately 500,000 men and women recruited between 2006 and 2010 from across the UK (age range at recruitment: 38 years to 73 years old) ^50^. UK Biobank participants have provided a range of information via questionnaires and interviews, including on sociodemographic, lifestyle, health, and reproductive factors; as well as biological samples and physical measures (data available at www.ukbiobank.ac.uk). A subset of approximately 20,000 were selected for repeat assessment between 2012 and 2013. A full description of the study design, participants and quality control (QC) methods have been described in detail previously ^51^. UK Biobank received ethical approval from the Research Ethics Committee (REC reference for UK Biobank is 11/NW/0382). The current work was approved under UK Biobank Project 30418 and 81499.

### Reproductive traits

Women were asked a detailed set of questions about their reproductive health via a self-reported questionnaire. Parity was based on the number of live births reported whilst in men number of children were reported. Age at menarche and age at natural menopause were reported in years. Age at natural menopause therefore excluded women who had not yet gone through the menopause or who had a surgical menopause. Those who had not gone through a natural menopause (N=25,70) had either (i) not yet gone through the menopause (15,418, 60%) (ii) had a surgical menopause or other (10,322, 40%) (**Table 1, Supplementary Table 3**).

### NMR metabolic measures

Metabolic traits were measured using a targeted high-throughput NMR metabolomics (Nightingale Health Ltd; biomarker quantification version 2020)^52^. This platform provides simultaneous quantification of 249 metabolic measures, consisting of concentrations of 165 metabolic measures and 84 derived ratios, encompassing routine lipids, lipoprotein subclass profiling (including lipid composition within 14 subclasses), fatty acid composition, and various low-molecular weight metabolites such as amino acids, ketone bodies and glycolysis metabolites. Technical details and epidemiological applications have been previously reviewed ^18,53^. Pre-release data from a random subset of 126,846 non-fasting plasma samples collected at baseline or first repeat assessment were made available to early access analysts. 121,577 samples were retained for analyses after removing duplicates and observations not passing quality control (QC) (i.e. sample QC flag “Low protein”, biomarker QC flag “Technical error”, or samples with insufficient material). All metabolic measures were standardised and normalised prior to analyses using rank-based inverse normal transformation.

### Clinical chemistry measures

We used data on the eight biomarkers assayed using clinical chemistry techniques, as previously described ^54^, that matched measures in the NMR metabolomics platform — i.e. albumin, apolipoprotein A1, apolipoprotein B, glucose, HDL-cholesterol, LDL-cholesterol, total cholesterol, and triglycerides. These measures are available in most UK Biobank participants and were used in Mendelian randomization analyses, as described under ‘Statistical analyses’, to increase statistical power and check agreement with results from NMR metabolic measures. All biomarkers were standardised and normalised prior to analyses using rank-based inverse normal transformation.

### Summary data on genetic associations with metabolic measures

Genotype data was available for 488,377 UK Biobank participants, of which 49,979 were genotyped using the UK BiLEVE array and 438,398 using the UK Biobank axiom array. Pre-imputation QC, phasing and imputation are described elsewhere ^55^. Genotype imputation was performed using IMPUTE2 algorithms ^56^ to a reference set combining the UK10K haplotype and HRC reference panels ^57^. Post-imputation QC was performed as described in the “UK Biobank Genetic Data: MRC-IEU Quality Control” documentation ^58^. Genetic association data for metabolic measures was generated using the MRC IEU UK Biobank GWAS pipeline ^59^. Briefly, we restricted the sample to individuals of ‘European’ ancestry as defined by the largest cluster in an in-house k-means cluster analysis performed using the first 4 principal components provided by UK Biobank in the statistical software environment R (n=464,708). Genome-wide association analysis (GWAS) was conducted using linear mixed model (LMM) association method as implemented in BOLT-LMM (v2.3) ^60^. Population structure was modelled using 143,006 directly genotyped SNPs (MAF > 0.01; genotyping rate > 0.015; Hardy-Weinberg equilibrium p-value < 0.0001 and LD pruning to an r2 threshold of 0.1 using PLINKv2.00). Models were adjusted for genotyping array and fasting time and were restricted to the subsample of women.

### Covariables

For multivariable analyses, confounders were defined a priori based on them being known or plausible causal factors for reproductive traits and cardiovascular risk via higher/lower metabolites. A minimal set of adjustments were made in the main multivariable regression analyses as most confounders were not assessed prior to or around when the reproductive traits occurred. Specifically, we adjusted for education as a categorical variable (University, A-levels, O levels (or equivalent) or other), age at baseline and retrospectively reported body size at age 10 (average, thinner, plumper) in all regression analyses. In additional analyses we also partially adjusted for the full set of defined confounders using baseline measurements (mostly after the occurrence of exposures) as correlates of the before exposure measures (see below in statistical analyses).

### Statistical analyses

We used multiple approaches (i.e. multivariable regression, negative control and MR) relying on different assumptions to explore the causal role of reproductive traits on later life metabolic profile. All analysis was conducted using Stata16 (StataCorp, College Station, TX) or R 4.1.1 (R Foundation for Statistical Computing, Vienna, Austria) and results presented as differences in means for each metabolic trait in standard deviation (SD) units per 1 child difference for number of children and per 1 year difference for age at menarche and age at menopause, facilitating the comparison of results from different methods.

For both multivariable and MR analyses, we corrected for multiple testing using the Bonferroni method considering 3*18=54 independent tests (*P*=0.05/54≈0.00093). This was based on the three exposures included in our analyses (i.e. age at menarche, parity, and age at natural menopause) and the 18 independent features explaining over 95% of variance in the highly correlated NMR metabolic measures in our dataset as estimated by principal component analysis ^61^.

#### (a) Multivariable regression

In the main analyses we used linear regression, with three sets of models: (1) no adjustments, (2) adjusted for education, age at baseline and body composition at age 10 and (3) model (2) additionally adjusted for baseline variables collected at the first assessment at (mean) age 56 years (SD=8) including BMI, smoking and alcohol status. By adjusting for the baseline variables at the first assessment we can either block the confounding path or create bias if these variables are mediators. If the results change between model (2) and (3) it is hard to distinguish whether its correct adjustment for confounding or whether it is a mediated path. Because of this we considered model (2) to be the best causal estimate and present models (1) and (3) in supplementary material. For age at menarche, education will have been measured after the exposure. However, as it is influenced by parental education, income and occupation (occurring before menarche) unlikely to be determined by age at menarche, we a priori considered a proxy of early life ^62^. In sensitivity analyses we assessed whether there was a non-linear relationship between each reproductive trait and 55 non-derived metabolites. For ease of presentation, we excluded measures that were derived (eg ratios) or related to lipoprotein subfractions as these are highly correlated with one or more of the 55 non-derived metabolites. We compared the categorised reproductive trait entered into the model as a categorical variable and as a continuous variable using a likelihood ratio test. Age at menarche and age at menopause were categorised into tertiles (<13, 13-14, >14 years) and quartiles (<49, 49-50, 51-53, >53 years), respectively. Parity was categorised as 0, 1, 2, and 3+. Results were plotted against the first reference category and the p-value for linear trend reported. For any metabolites that showed evidence of non-linearity, restricted cubic splines (with either 3, 4, or 5 knots placed at percentiles as suggested by Harrell^63^ for each reproductive trait) were fit and compared to the main analysis model (assuming a linear association) using AIC (BIC and root mean square error also shown).

#### (b) Negative control analyses

Negative control analyses aim to emulate a condition that cannot involve the hypothesized causal mechanism but is likely to have similar sources of bias that may have been present in the association of interest ^5,27^. We used males as negative controls to assess potential biases in the association between parity (proxied by number of live births) and metabolic measures in women. If associations between number of live births and metabolic measures in women reflect a causal effect of parity on women’s metabolic health, one would expect number of live births to be associated with metabolic measures in women but not in men given men do not experience pregnancy. Similar to the multivariable regression analyses, we test the association between number of children (men) and their measured metabolites and present three sets of models: (1) with no adjustments, (2) adjusted for education, age at baseline and retrospectively self-reported body composition at age 10 and (3) model (2) additionally adjusted for baseline variables collected at the first assessment at (mean) age 56 years (SD=8) including BMI, smoking and alcohol status.

#### (c) Mendelian randomization

We used two-sample MR to explore the effect of older age at menarche, higher parity, and older age at natural menopause on women’s metabolic profile. Publicly available GWAS summary data were used for SNP-reproductive traits associations (sample 1) and UK Biobank summary GWAS data for SNP-metabolite associations (sample 2). This approach does not require all participants to have data on both exposure and outcome, and, therefore, allows us to retain the largest possible sample sizes, meaning that power to detect a causal effect is increased ^64^.

### Selection of genetic instruments

#### Age at menarche

Genetic instruments were selected from a GWAS of age at menarche, which included 329,345 women of European ancestry (**Supplementary Table 14**) ^65^. Linear regression models were used to estimate the association between genetic variants and age at menarche (in years) adjusting for age at study visit and study-specific covariables. For our analyses, we selected the 389 independent SNPs reported by the GWAS to be strongly associated with age at menarche (P-value < 5*10^-8^) in the discovery metanalyses. Given the age at menarche GWAS included UK Biobank participants (maximum estimated sample overlap: ∼20%), we have also selected an additional set of age at menarche-associated genetic variants (N = 68 SNPs) using data from a previous GWAS that did not including UK Biobank (details in ‘Sensitivity analyses’ below and **Supplementary Table 14**) ^66^.

#### Parity

Genetic instruments were selected from a GWAS of number of children ever born, as a proxy of parity, which included 785,604 men and women of European ancestry from 45 studies (**Supplementary Table 14**) ^67^. Number of children ever born was treated as a continuous measure and included both parous and nulliparous women. Linear regression models were used to estimate the association between genetic variants and number of children ever born adjusting for principal components of ancestry, birth year, its square and cubic, to control for non-linear birth cohort effects. Family-based studies controlled for family structure or excluded relatives. The sex-combined metanalysis also included interactions of birth year and its polynomials with sex. For our analyses, we used the 32 independent SNPs reported by the GWAS to be strongly associated with number of children ever born (P-value < 5*10^-8^) in either the sex-combined (28 SNPs) or female-specific (4 SNPs) metanalyses and summary association data from the female-specific metanalyses. The GWAS included UK Biobank (maximum estimated sample overlap: 14%).

#### Age at natural menopause

Genetic instruments were selected from a GWAS of age at natural menopause conducted in 201,323 women of European ancestry (**Supplementary Table 14**) ^17^. Linear regression models were used to estimate the association between genetic variants and age at natural menopause (in years). For our analyses, we selected 290 SNPs reported by the GWAS to be strongly associated with age at natural menopause (P-value < 5*10^-8^). Where available, we used association data from the sample combining discovery and replication stages (N = 496,151). Given the age at menarche GWAS included UK Biobank participants (maximum estimated sample overlap: 13% considering the GWAS combined discovery and replication samples), we have also selected an additional set of age at natural menopause-associated genetic variants (N = 42 SNPs) using data from a previous GWAS that did not include UK Biobank (details in ‘Sensitivity analyses’ below and **Supplementary Table 14**) ^68^.

### Main analyses

We used a standard two-sample MR method, the inverse variance weighted (IVW) estimator, to explore the effect of age at menarche, parity and age at natural menopause on women’s metabolic profile by combining genetic association estimates for reproductive traits (extracted from published GWASes data) with genetic association estimates for the metabolic measures (generated from UK Biobank data). Given a priori evidence of a potential bidirectional relationship between age at menarche and BMI, we also used multivariable IVW to test the effect of age at menarche on metabolic measures accounting for adult BMI. For multivariable IVW analysis, apart from the data previously described, we used summary genetic association data for BMI extracted from the 2015 metanalysis by the GIANT consortium (N = 339,224 individuals not including UK Biobank participants) ^69^.

### Sensitivity analyses

Several sensitivity analyses were conducted to explore the plausibility of the three core MR assumptions, which are required for the method to provide a valid test of the presence of a causal effect.

#### Assumption 1: the genetic instrument must be associated with the reproductive trait

We selected genetic variants reported to be strongly associated with reproductive in the largest available GWAS and estimated the proportion of phenotypic variance explained (R^2^) and F-statistics for the association of SNPs with reproductive traits among females as an indicator of instrument strength.

#### Assumption 2: the association between genetic instrument and outcome is unconfounded

One of the main motivations for using MR is to avoid unmeasured confounding. However, there is growing evidence that, in some instances, MR studies can be confounded when using data from unrelated individuals due to population stratification, assortative mating and indirect genetic effects of parents ^70,71^. We used two approaches to explore whether these were likely to bias our main results. First, we used sex-combined data from a recent within-sibship GWAS, including up to 159,701 siblings from 17 cohorts, to test the effect of genetic susceptibility to higher age at menarche, parity and age at menopause on metabolic markers (i.e. LDL-cholesterol, triglycerides, HDL-cholesterol, C-reactive protein, and glycated haemoglobin) ^70^. C-reactive protein and glycated haemoglobin were used as proxies for inflammation and hyperglycaemia, respectively, given GlycA and glucose were not available. Within-sibling MR designs control for variation in parental genotypes, and so should not be affected by population stratification, assortative mating and indirect genetic effects of parents ^70-72^. Second, we performed IVW on negative control outcomes (i.e. skin colour and skin tanning ability) since these could not conceivably be affected by the exposures and any evidence for an association between reproductive traits and, these negative control outcomes would be indicative of residual population stratification in the exposure GWAS ^73^.

#### Assumption 3: the genetic instrument does not affect the outcome except through its possible effect on the exposure

A key violation of this assumption is known as horizontal pleiotropy, where genetic variants influence the outcome through pathways that are not mediated by the exposure ^74^. We explored the presence of bias due to horizontal pleiotropy by using other MR methods: the weighted median estimator and MR-Egger. These methods can provide valid tests of a causal effect under different (and weaker) assumptions about the nature of the underlying horizontal pleiotropy. The weighted median estimator requires that at least 50% of the weight in the analysis stems from valid instruments. The MR-Egger estimator assumes that the instrument strength is independent of its the direct effects on the outcome (i.e. INSIDE assumption).

In addition to the core assumptions, the two-sample MR approach assumes that genetic associations with exposure and outcome were estimated from two comparable but non-overlapping samples. We restricted our analyses to European adult individuals to ensure that samples were comparable. We assessed potential bias due to sample overlap by conducting MR using SNPs selected from previous GWAS of age at menarche and age at natural menopause that did not include UK Biobank (**Supplementary Table 14**).

## Data Availability

Access to UK Biobank data can be obtained under registration and application to the Access Management System (AMS) (https://www.ukbiobank.ac.uk/enable-your-research/register). GWAS summary data for reproductive traits is publicly available as detailed in Supplementary table 4.

## Acknowledgements

We are extremely grateful to Nightingale for the use of their data and for their helpful discussions throughout. We want to acknowledge participants and investigators from UK Biobank and the multiple large-scale GWAS consortia which made summary data available. This work used the computational facilities of the Advanced Computing Research Centre, University of Bristol - http://www.bristol.ac.uk/acrc/. The current analysis was approved under UK Biobank Project 30418 and 81499. We are also grateful to Professor Kate Tilling (University of Bristol), Prof Zoltan Kutalik, and Leona Knusel (University of Lausanne) who helped us with additional analyses undertaken to explore discrepant results between multivariable regression and two sample MR for the association of age at natural menopause with biomarkers.

## Availability of data and materials

Analysis scripts and the analysis plan can be found on the following GitHub page: https://github.com/gc13313/nmr_repro.

## Ethical approval and consent to participate

UK Biobank received ethical approval from the Research Ethics Committee (REC reference for UK Biobank is 11/NW/0382). The current analysis was approved under UK Biobank Project 30418 and 81499.

## Funding

This research is supported by the University of Bristol and UK Medical Research Council (MRC) (MC_UU_00011/6, all authors), the European Union’s Horizon 2020 research and innovation programme under grant agreement No 733206 LifeCycle (GLC and DAL), a University of Bristol Vice-Chancellor’s Fellowship (MCB), the British Heart Foundation (AA/18/7/34219, MCB and DAL and CH/F/20/90003, DAL) and the UK National Institute of Health Research (NF-0616-10102, DAL).

The funders had no role in study design, data collection and analysis, decision to publish, or preparation of the manuscript. For the purpose of Open Access, the author has applied a CC BY public copyright licence to any Author Accepted Manuscript version arising from this submission.

This publication is the work of the authors and all authors will serve as guarantors for the contents of this paper.

## Author contributions

DAL conceived the study. DAL, MCB and GLC designed the study. MCB and GLC performed the analyses. MCB and GLC wrote the original draft of the manuscript with input from DAL. All authors were involved in the interpretation of results, helped refine the manuscript, and approved its final version.

## Abbreviations

Ala: Alanine
ApoA1: Apolipoprotein A1
ApoB: Apolipoprotein B
ApoB_by_ApoA1: Ratio of apolipoprotein B to apolipoprotein A1 Cholines Total cholines
Clinical_LDL_C: Clinical LDL cholesterol
DHA: Docosahexaenoic acid
DHA_pct: Ratio of docosahexaenoic acid to total fatty acids
Gln: Glutamine
Gly: Glycine
GlycA: Glycoprotein acetyls
HDL_C: HDL cholesterol
HDL_CE: Cholesteryl esters in HDL
HDL_FC: Free cholesterol in HDL
HDL_L: Total lipids in HDL
HDL_P: Concentration of HDL particles
HDL_PL: Phospholipids in HDL
HDL_TG: Triglycerides in HDL
HDL_size: Average diameter for HDL particles
His: Histidine
IDL_C: Cholesterol in IDL
IDL_CE: Cholesteryl esters in IDL
IDL_CE_pct: Cholesteryl esters to total lipids ratio in IDL
IDL_C_pct: Cholesterol to total lipids ratio in IDL
IDL_FC: Free cholesterol in IDL
IDL_FC_pct: Free cholesterol to total lipids ratio in IDL
IDL_L: Total lipids in IDL
IDL_P: Concentration of IDL particles
IDL_PL: Phospholipids in IDL
IDL_PL_pct: Phospholipids to total lipids ratio in IDL
IDL_TG: Triglycerides in IDL
IDL_TG_pct: Triglycerides to total lipids ratio in IDL
Ile: Isoleucine
LA: Linoleic acid
LA_pct: Ratio of linoleic acid to total fatty acids
LDL_C: LDL cholesterol
LDL_CE: Cholesteryl esters in LDL
LDL_FC: Free cholesterol in LDL
LDL_L: Total lipids in LDL
LDL_P: Concentration of LDL particles
LDL_PL: Phospholipids in LDL
LDL_TG: Triglycerides in LDL
LDL_size: Average diameter for LDL particles
L_HDL_C: Cholesterol in large HDL
L_HDL_CE: Cholesteryl esters in large HDL
L_HDL_CE_pct: Cholesteryl esters to total lipids ratio in large HDL
L_HDL_C_pct: Cholesterol to total lipids ratio in large HDL
L_HDL_FC: L_HDL_L Total lipids in large HDL
L_HDL_P: Concentration of large HD particles
L_HDL_PL: Phospholipids in large HDL
L_HDL_PL_pct: Phospholipids to total lipids ratio in large HDL
L_HDL_TG: Triglycerides in large HDL
L_HDL_TG_pct: Triglycerides to total lipids ratio in large
L_LDL_C: Cholesterol in large LDL
L_LDL_CE: Cholesteryl esters in large LDL
L_LDL_CE_pct: Cholesteryl esters to total lipids ratio in large LDL
L_LDL_C_pct: Cholesterol to total lipids ratio in large LDL
L_LDL_FC: Free cholesterol in large LDL
L_LDL_FC_pct: Free cholesterol to total lipids ratio in large LDL
L_LDL_L: Total lipids in large LDL
L_LDL_P: Concentration of large LDL particles
L_LDL_PL: Phospholipids in large LDL
L_LDL_PL_pct: Phospholipids to total lipids ratio in large LDL
L_LDL_TG: Triglycerides in large LDL
L_LDL_TG_pct: Triglycerides to total lipids ratio in large LDL
L_VLDL_C: Cholesterol in large VLDL
L_VLDL_CE: Cholesteryl esters in large VLDL
L_VLDL_CE_pct: Cholesteryl esters to total lipids ratio in large VLDL
L_VLDL_C_pct: Cholesterol to total lipids ratio in large VLDL
L_VLDL_FC: Free cholesterol in large VLDL
L_VLDL_FC_pct: Free cholesterol to total lipids ratio in large VLDL
L_VLDL_L: Total lipids in large VLDL
L_VLDL_P: Concentration of large VLDL particles
L_VLDL_PL: Phospholipids in large VLDL
L_VLDL_PL_pct: Phospholipids to total lipids ratio in large VLDL
L_VLDL_TG: Triglycerides in large VLDL
L_VLDL_TG_pct: Triglycerides to total lipids ratio in large VLDL
Leu: Leucine
MUFA: Monounsaturated fatty acids
MUFA_pct: Ratio of monounsaturated fatty acids to total fatty acids
M_HDL_C: Cholesterol in medium HDL
M_HDL_CE: Cholesteryl esters in medium HDL
M_HDL_CE_pct: Cholesteryl esters to total lipids ratio in medium HDL
M_HDL_C_pct: Cholesterol to total lipids ratio in medium HDL
M_HDL_FC: Free cholesterol in medium HDL
M_HDL_FC_pct: Free cholesterol to total lipids ratio in medium HDL
M_HDL_L: Total lipids in medium HDL
M_HDL_P: Concentration of medium HDL particles
M_HDL_PL: Phospholipids in medium HDL
M_HDL_PL_pct: Phospholipids to total lipids ratio in medium HDL
M_HDL_TG: Triglycerides in medium HDL
M_HDL_TG_pct: Triglycerides to total lipids ratio in medium HDL
M_LDL_C: Cholesterol in medium LDL
M_LDL_CE: Cholesteryl esters in medium LDL
M_LDL_CE_pct: Cholesteryl esters to total lipids ratio in medium LDL
M_LDL_C_pct: Cholesterol to total lipids ratio in medium LDL M_LDL_FC Free cholesterol in medium LDL
M_LDL_FC_pct: Free cholesterol to total lipids ratio in medium LDL M_LDL_L Total lipids in medium LDL
M_LDL_P: Concentration of medium LDL particles
M_LDL_PL: Phospholipids in medium LDL
M_LDL_PL_pct: Phospholipids to total lipids ratio in medium LDL M_LDL_TG Triglycerides in medium LDL
M_LDL_TG_pct: Triglycerides to total lipids ratio in medium LDL M_VLDL_C Cholesterol in medium VLDL
M_VLDL_CE: Cholesteryl esters in medium VLDL
M_VLDL_CE_pct: Cholesteryl esters to total lipids ratio in medium VLDL M_VLDL_C_pct Cholesterol to total lipids ratio in medium VLDL M_VLDL_FC Free cholesterol in medium VLDL
M_VLDL_FC_pct: Free cholesterol to total lipids ratio in medium VLDL M_VLDL_L Total lipids in medium VLDL
M_VLDL_P: Concentration of medium VLDL particles
M_VLDL_PL: Phospholipids in medium VLDL
M_VLDL_PL_pct: Phospholipids to total lipids ratio in medium VLDL
M_VLDL_TG: Triglycerides in medium VLDL
M_VLDL_TG_pct: Triglycerides to total lipids ratio in medium VLDL
Omega_3: Omega-3 fatty acids
Omega_3_pct: Ratio of omega-3 fatty acids to total fatty acids
Omega_6: Omega-6 fatty acids
Omega_6_by_Omega_3: Ratio of omega-6 fatty acids to omega-3 fatty acids
Omega_6_pct: Ratio of omega-6 fatty acids to total fatty acids
PUFA: Polyunsaturated fatty acids
PUFA_by_MUFA: Ratio of polyunsaturated fatty acids to monounsaturated fatty acids
PUFA_pct: Ratio of polyunsaturated fatty acids to total fatty acids
Phe: Phenylalanine
Phosphatidylc: Phosphatidylcholines
Phosphoglyc: Phosphoglycerides
Remnant_C: Remnant cholesterol (non-HDL, non-LDL -cholesterol)
SFA: Saturated fatty acids
SFA_pct: Ratio of saturated fatty acids to total fatty acids
S_HDL_C: Cholesterol in small HDL
S_HDL_CE: Cholesteryl esters in small HDL
S_HDL_CE_pct: Cholesteryl esters to total lipids ratio in small HDL
S_HDL_C_pct: Cholesterol to total lipids ratio in small HDL
S_HDL_FC: Free cholesterol in small HDL
S_HDL_FC_pct: Free cholesterol to total lipids ratio in small HDL
S_HDL_L: Total lipids in small HDL
S_HDL_P: Concentration of small HDL particles
S_HDL_PL: Phospholipids in small HDL
S_HDL_PL_pct: Phospholipids to total lipids ratio in small HDL
S_HDL_TG: Triglycerides in small HDL
S_HDL_TG_pct: Triglycerides to total lipids ratio in small HDL
S_LDL_C: Cholesterol in small LDL
S_LDL_CE: Cholesteryl esters in small LDL
S_LDL_CE_pct: Cholesteryl esters to total lipids ratio in small LDL
S_LDL_C_pct: Cholesterol to total lipids ratio in small LDL
S_LDL_FC: Free cholesterol in small LDL
S_LDL_FC_pct: Free cholesterol to total lipids ratio in small LDL
S_LDL_L: Total lipids in small LDL
S_LDL_P: Concentration of small LDL particles
S_LDL_PL: Phospholipids in small LDL
S_LDL_PL_pct: Phospholipids to total lipids ratio in small LDL
S_LDL_TG: Triglycerides in small LDL
S_LDL_TG_pct: Triglycerides to total lipids ratio in small LDL
S_VLDL_C: Cholesterol in small VLDL
S_VLDL_CE: Cholesteryl esters in small VLDL
S_VLDL_CE_pct: Cholesteryl esters to total lipids ratio in small VLDL
S_VLDL_C_pct: Cholesterol to total lipids ratio in small VLDL
S_VLDL_FC: Free cholesterol in small VLDL
S_VLDL_FC_pct: Free cholesterol to total lipids ratio in small VLDL
S_VLDL_L: Total lipids in small VLDL
S_VLDL_P: Concentration of small VLDL particles
S_VLDL_PL: Phospholipids in small VLDL
S_VLDL_PL_pct: Phospholipids to total lipids ratio in small VLDL
S_VLDL_TG: Triglycerides in small VLDL
S_VLDL_TG_pct: Triglycerides to total lipids ratio in small VLDL TG_by_PG Ratio of triglycerides to phosphoglycerides
Total_BCAA: Total concentration of branched-chain amino acids (leucine + isoleucine + valine)
Total_C: Total cholesterol
Total_CE: Total esterified cholesterol
Total_FA: Total fatty acids
Total_FC: Total free cholesterol
Total_L: Total lipids in lipoprotein particles
Total_P: Total concentration of lipoprotein particles
Total_PL: Total phospholipids in lipoprotein particles
Total_TG: Total triglycerides
Tyr: Tyrosine
Unsaturation: Degree of unsaturation
VLDL_C: VLDL cholesterol
VLDL_CE: Cholesteryl esters in VLDL
VLDL_FC: Free cholesterol in VLDL
VLDL_L: Total lipids in VLDL
VLDL_P: Concentration of VLDL particles
VLDL_PL: Phospholipids in VLDL
VLDL_TG: Triglycerides in VLDL
VLDL_size: Average diameter for VLDL particles
Val: Valine
XL_HDL_C: Cholesterol in very large HDL
XL_HDL_CE: Cholesteryl esters in very large HDL
XL_HDL_CE_pct: Cholesteryl esters to total lipids ratio in very large
HDL: XL_HDL_C_pct Cholesterol to total lipids ratio in very large HDL
XL_HDL_FC: Free cholesterol in very large HDL
XL_HDL_FC_pct: Free cholesterol to total lipids ratio in very large HDL
XL_HDL_L: Total lipids in very large HDL
XL_HDL_P: Concentration of very large HDL particles
XL_HDL_PL: Phospholipids in very large HDL
XL_HDL_PL_pct: Phospholipids to total lipids ratio in very large HDL
XL_HDL_TG: Triglycerides in very large HDL
XL_HDL_TG_pct: Triglycerides to total lipids ratio in very large HDL
XL_VLDL_C: Cholesterol in very large VLDL
XL_VLDL_CE: Cholesteryl esters in very large VLDL
XL_VLDL_CE_pct: Cholesteryl esters to total lipids ratio in very large VLDL
XL_VLDL_C_pct: Cholesterol to total lipids ratio in very large VLDL
XL_VLDL_FC: Free cholesterol in very large VLDL
XL_VLDL_FC_pct: Free cholesterol to total lipids ratio in very large VLDL
XL_VLDL_L: Total lipids in very large VLDL
XL_VLDL_P: Concentration of very large VLDL particles
XL_VLDL_PL: Phospholipids in very large VLDL
XL_VLDL_PL_pct: Phospholipids to total lipids ratio in very large VLDL
XL_VLDL_TG: Triglycerides in very large VLDL
XL_VLDL_TG_pct: Triglycerides to total lipids ratio in very large VLDL
XS_VLDL_C: Cholesterol in very small VLDL
XS_VLDL_CE: Cholesteryl esters in very small VLDL
XS_VLDL_CE_pct: Cholesteryl esters to total lipids ratio in very small VLDL
XS_VLDL_C_pct: Cholesterol to total lipids ratio in very small VLDL
XS_VLDL_FC: Free cholesterol in very small VLDL
XS_VLDL_FC_pct: Free cholesterol to total lipids ratio in very small VLDL
XS_VLDL_L: Total lipids in very small VLDL
XS_VLDL_P: Concentration of very small VLDL particles
XS_VLDL_PL: Phospholipids in very small VLDL
XS_VLDL_PL_pct: Phospholipids to total lipids ratio in very small VLDL
XS_VLDL_TG: Triglycerides in very small VLDL
XS_VLDL_TG_pct: Triglycerides to total lipids ratio in very small VLDL
XXL_VLDL_C: Cholesterol in chylomicrons and extremely large VLDL
XXL_VLDL_CE: Cholesteryl esters in chylomicrons and extremely large VLDL
XXL_VLDL_CE_pct: Cholesteryl esters to total lipids ratio in chylomicrons and extremely large VLDL
XXL_VLDL_C_pct: Cholesterol to total lipids ratio in chylomicrons and extremely large VLDL
XXL_VLDL_FC: Free cholesterol in chylomicrons and extremely large VLDL
XXL_VLDL_FC_pct: Free cholesterol to total lipids ratio in chylomicrons and extremely large VLDL
XXL_VLDL_L: Total lipids in chylomicrons and extremely large VLDL
XXL_VLDL_P: Concentration of chylomicrons and extremely large VLDL particles
XXL_VLDL_PL: Phospholipids in chylomicrons and extremely large VLDL
XXL_VLDL_PL_pct: Phospholipids to total lipids ratio in chylomicrons and extremely large VLDL
XXL_VLDL_TG: Triglycerides in chylomicrons and extremely large VLDL
XXL_VLDL_TG_pct: Triglycerides to total lipids ratio in chylomicrons and extremely large VLDL
bOHbutyrate: 3-Hydroxybutyrate
non_HDL_C: Total cholesterol minus HDL-C

## References

1 Ness, R. B., Schotland, H. M., Flegal, K. M. & Shofer, F. S. Reproductive History and Coronary Heart Disease Risk in Women. Epidemiologic Reviews 16, 298–314 (1994). https://doi.org:10.1093/oxfordjournals.epirev.a036155

2 Canoy, D. et al. Age at Menarche and Risks of Coronary Heart and Other Vascular Diseases in a Large UK Cohort. Circulation 131, 237–244 (2015). https://doi.org:10.1161/CIRCULATIONAHA.114.010070

3 Cooper, G. S. et al. Menstrual and Reproductive Risk Factors for Ischemic Heart Disease. Epidemiology 10, 255–259 (1999).

4 Green, A., Beral, V. & Moser, K. Mortality in women in relation to their childbearing history. British Medical Journal 297, 391 (1988). https://doi.org:10.1136/bmj.297.6645.391

5 Lawlor Debbie, A., et al. Is the Association Between Parity and Coronary Heart Disease Due to Biological Effects of Pregnancy or Adverse Lifestyle Risk Factors Associated With Child-Rearing? Circulation 107, 1260–1264 (2003). https://doi.org:10.1161/01.CIR.0000053441.43495.1A

6 Colditz, G. A. et al. A PROSPECTIVE STUDY OF AGE AT MENARCHE, PARITY, AGE AT FIRST BIRTH, AND CORONARY HEART DISEASE IN WOMEN. American Journal of Epidemiology 126, 861–870 (1987). https://doi.org:10.1093/oxfordjournals.aje.a114723

7 Kelsey, J. L., Gammon, M. D. & John, E. M. Reproductive Factors and Breast Cancer. Epidemiologic Reviews 15, 36–47 (1993). https://doi.org:10.1093/oxfordjournals.epirev.a036115

8 Lambe, M. et al. Parity, age at first and last birth, and risk of breast cancer: A population-based study in Sweden. Breast Cancer Research and Treatment 38, 305–311 (1996). https://doi.org:10.1007/BF01806150

9 Gong, T.-T., Wu, Q.-J., Vogtmann, E., Lin, B. & Wang, Y.-L. Age at menarche and risk of ovarian cancer: a meta-analysis of epidemiological studies. Int J Cancer 132, 2894–2900 (2013). https://doi.org:10.1002/ijc.27952

10 Whiteman, D. C., Siskind, V., Purdie, D. M. & Green, A. C. Timing of Pregnancy and the Risk of Epithelial Ovarian Cancer. Cancer Epidemiology Biomarkers & Prevention 12, 42 (2003).

11 Adami, H. O. et al. Parity, age at first childbirth, and risk of ovarian cancer. The Lancet 344, 1250–1254 (1994). https://doi.org:https://doi.org/10.1016/S0140-6736(94)90749-8

12 Dossus, L. et al. Reproductive risk factors and endometrial cancer: the European Prospective Investigation into Cancer and Nutrition. Int J Cancer 127, 442–451 (2010). https://doi.org:https://doi.org/10.1002/ijc.25050

13 Burgess, S. et al. Dissecting Causal Pathways Using Mendelian Randomization with Summarized Genetic Data: Application to Age at Menarche and Risk of Breast Cancer. Genetics 207, 481–487 (2017). https://doi.org:10.1534/genetics.117.300191

14 Mumby, H. S. et al. Mendelian Randomisation Study of Childhood BMI and Early Menarche. Journal of Obesity 2011, 180729 (2011). https://doi.org:10.1155/2011/180729

15 Mills, M. C. et al. Identification of 370 loci for age at onset of sexual and reproductive behaviour, highlighting common aetiology with reproductive biology, externalizing behaviour and longevity. bioRxiv, 2020.2005.2006.081273 (2020). https://doi.org:10.1101/2020.05.06.081273

16 Barban, N. et al. Genome-wide analysis identifies 12 loci influencing human reproductive behavior. Nature genetics 48, 1462–1472 (2016). https://doi.org:10.1038/ng.3698

17 Ruth, K. S. et al. Genetic insights into the biological mechanisms governing human ovarian ageing. medRxiv, 2021.2001.2011.20248322 (2021). https://doi.org:10.1101/2021.01.11.20248322

18 Wurtz, P. et al. Quantitative Serum Nuclear Magnetic Resonance Metabolomics in Large-Scale Epidemiology: A Primer on -Omic Technologies. Am J Epidemiol 186, 1084–1096 (2017). https://doi.org:10.1093/aje/kwx016

19 Würtz, P. et al. Metabolite profiling and cardiovascular event risk: a prospective study of 3 population-based cohorts. Circulation 131, 774–785 (2015). https://doi.org:10.1161/CIRCULATIONAHA.114.013116

20 Wang, Q. et al. Metabolic profiling of pregnancy: cross-sectional and longitudinal evidence. BMC Medicine 14, 205 (2016). https://doi.org:10.1186/s12916-016-0733-0

21 Wang, Q. et al. Metabolic characterization of menopause: cross-sectional and longitudinal evidence. BMC medicine 16, 17–17 (2018). https://doi.org:10.1186/s12916-018-1008-8

22 Wang, Q. et al. Effects of hormonal contraception on systemic metabolism: cross-sectional and longitudinal evidence. International journal of epidemiology 45, 1445–1457 (2016). https://doi.org:10.1093/ije/dyw147

23 Lane, A. N., Higashi, R. M. & Fan, T. W. M. NMR and MS-based Stable Isotope-Resolved Metabolomics and Applications in Cancer Metabolism. Trends Analyt Chem 120, 115322 (2019). https://doi.org:10.1016/j.trac.2018.11.020

24 Johnson, K. E. et al. The relationship between circulating lipids and breast cancer risk: A Mendelian randomization study. PLoS medicine 17, e1003302–e1003302 (2020). https://doi.org:10.1371/journal.pmed.1003302

25 Chen, W., Wang, S., Lv, W. & Pan, Y. Causal associations of insulin resistance with coronary artery disease and ischemic stroke: a Mendelian randomization analysis. BMJ Open Diabetes Research & Care 8, e001217 (2020). https://doi.org:10.1136/bmjdrc-2020-001217

26 Xu, L., Borges, M. C., Hemani, G. & Lawlor, D. A. The role of glycaemic and lipid risk factors in mediating the effect of BMI on coronary heart disease: a two-step, two-sample Mendelian randomisation study. Diabetologia 60, 2210–2220 (2017). https://doi.org:10.1007/s00125-017-4396-y

27 Lawlor, D. A., Tilling, K. & Davey Smith, G. Triangulation in aetiological epidemiology. International Journal of Epidemiology 45, 1866–1886 (2016). https://doi.org:10.1093/ije/dyw314

28 Bell, J. A. et al. Influence of puberty timing on adiposity and cardiometabolic traits: A Mendelian randomisation study. PLoS medicine 15, e1002641–e1002641 (2018). https://doi.org:10.1371/journal.pmed.1002641

29 Kirkpatrick, R. M., McGue, M., Iacono, W. G., Miller, M. B. & Basu, S. Results of a “GWAS Plus:” General Cognitive Ability Is Substantially Heritable and Massively Polygenic. PLOS ONE 9, e112390 (2014). https://doi.org:10.1371/journal.pone.0112390

30 Rich-Edwards, J. W., Fraser, A., Lawlor, D. A. & Catov, J. M. Pregnancy Characteristics and Women’s Future Cardiovascular Health: An Underused Opportunity to Improve Women’s Health? Epidemiologic Reviews 36, 57–70 (2014). https://doi.org:10.1093/epirev/mxt006

31 Sattar, N. & Greer, I. A. Pregnancy complications and maternal cardiovascular risk: opportunities for intervention and screening? BMJ (Clinical research ed.) 325, 157–160 (2002). https://doi.org:10.1136/bmj.325.7356.157

32 Li, W., Ruan, W., Lu, Z. & Wang, D. Parity and risk of maternal cardiovascular disease: A dose–response meta-analysis of cohort studies. European Journal of Preventive Cardiology 26, 592–602 (2019). https://doi.org:10.1177/2047487318818265

33 Hardy, R., Lawlor, D. A., Black, S., Wadsworth, M. E. J. & Kuh, D. Number of children and coronary heart disease risk factors in men and women from a British birth cohort. BJOG: An International Journal of Obstetrics & Gynaecology 114, 721–730 (2007). https://doi.org:https://doi.org/10.1111/j.1471-0528.2007.01324.x

34 Bridger Staatz, C. & Hardy, R. Number of children and body composition in later life among men and women: Results from a British birth cohort study. PLOS ONE 14, e0209529 (2019). https://doi.org:10.1371/journal.pone.0209529

35 Gunderson, E. P. et al. Childbearing is associated with higher incidence of the metabolic syndrome among women of reproductive age controlling for measurements before pregnancy: the CARDIA study. American Journal of Obstetrics and Gynecology 201, 177.e171–177.e179 (2009). https://doi.org:https://doi.org/10.1016/j.ajog.2009.03.031

36 Markovitz, A. R. et al. Does pregnancy alter life-course lipid trajectories? Evidence from the HUNT Study in Norway. Journal of lipid research 59, 2403–2412 (2018). https://doi.org:10.1194/jlr.P085720

37 Okoth, K. et al. Association between the reproductive health of young women and cardiovascular disease in later life: umbrella review. BMJ 371, m3502 (2020). https://doi.org:10.1136/bmj.m3502

38 de Kat, A. C. et al. Unraveling the associations of age and menopause with cardiovascular risk factors in a large population-based study. BMC Medicine 15, 2 (2017). https://doi.org:10.1186/s12916-016-0762-8

39 Auro, K. et al. A metabolic view on menopause and ageing. Nature Communications 5, 4708 (2014). https://doi.org:10.1038/ncomms5708

40 Karppinen, J. E. et al. Menopause modulates the circulating metabolome: evidence from a prospective cohort study. medRxiv, 2021.2012.2017.21266891 (2021). https://doi.org:10.1101/2021.12.17.21266891

41 Clayton, G. L. et al. Cardiovascular health in the menopause transition: a longitudinal study of up to 3892 women with up to four repeated measures of risk factors. BMC Medicine 20, 299 (2022). https://doi.org:10.1186/s12916-022-02454-6

42 Matthews, K. A. et al. Are Changes in Cardiovascular Disease Risk Factors in Midlife Women Due to Chronological Aging or to the Menopausal Transition? Journal of the American College of Cardiology 54, 2366–2373 (2009). https://doi.org:https://doi.org/10.1016/j.jacc.2009.10.009

43 Ruth, K. S. et al. Genetic insights into biological mechanisms governing human ovarian ageing. Nature 596, 393–397 (2021). https://doi.org:10.1038/s41586-021-03779-7

44 Salpeter, S. R. et al. Meta-analysis: effect of hormone-replacement therapy on components of the metabolic syndrome in postmenopausal women. Diabetes, Obesity and Metabolism 8, 538–554 (2006). https://doi.org:https://doi.org/10.1111/j.1463-1326.2005.00545.x

45 Manson, J. E. et al. Menopausal Hormone Therapy and Health Outcomes During the Intervention and Extended Poststopping Phases of the Women’s Health Initiative Randomized Trials. JAMA 310, 1353–1368 (2013). https://doi.org:10.1001/jama.2013.278040

46 Gartlehner, G. et al. U.S. Preventive Services Task Force Evidence Syntheses, formerly Systematic Evidence Reviews (Agency for Healthcare Research and Quality (US), Rockville (MD), 2017).

47 Day, F. R. et al. Large-scale genomic analyses link reproductive aging to hypothalamic signaling, breast cancer susceptibility and BRCA1-mediated DNA repair. Nature Genetics 47, 1294–1303 (2015). https://doi.org:10.1038/ng.3412

48 Munafò, M. R., Tilling, K., Taylor, A. E., Evans, D. M. & Davey Smith, G. Collider scope: when selection bias can substantially influence observed associations. International Journal of Epidemiology 47, 226–235 (2018). https://doi.org:10.1093/ije/dyx206

49 Soininen, P., Kangas, A. J., Würtz, P., Suna, T. & Ala-Korpela, M. Quantitative Serum Nuclear Magnetic Resonance Metabolomics in Cardiovascular Epidemiology and Genetics. Circulation: Cardiovascular Genetics 8, 192–206 (2015). https://doi.org:10.1161/CIRCGENETICS.114.000216

50 Allen, N. E., Sudlow, C., Peakman, T., Collins, R. & Biobank, U. K. UK biobank data: come and get it. Sci Transl Med 6, 224ed224 (2014). https://doi.org:10.1126/scitranslmed.3008601

51 Collins, R. What makes UK Biobank special? Lancet 379, 1173–1174 (2012). https://doi.org:10.1016/S0140-6736(12)60404-8

52 Julkunen, H., Cichonska, A., Slagboom, P. E., Wurtz, P. & Nightingale Health, U. K. B. I. Metabolic biomarker profiling for identification of susceptibility to severe pneumonia and COVID-19 in the general population. Elife 10 (2021). https://doi.org:10.7554/eLife.63033

53 Soininen, P., Kangas, A. J., Wurtz, P., Suna, T. & Ala-Korpela, M. Quantitative serum nuclear magnetic resonance metabolomics in cardiovascular epidemiology and genetics. Circ Cardiovasc Genet 8, 192–206 (2015). https://doi.org:10.1161/CIRCGENETICS.114.000216

54 Daniel Fry, R. A., Stewart Moffat, Mark Gordon & Parmesher Singh. UK Biobank Biomarker Project Companion Document to Accompany Serum Biomarker Data, <https://biobank.ndph.ox.ac.uk/showcase/showcase/docs/serum_biochemistry.pdf> (2019).

55 Bycroft, C. et al. The UK Biobank resource with deep phenotyping and genomic data. Nature 562, 203–209 (2018). https://doi.org:10.1038/s41586-018-0579-z

56 Howie, B., Marchini, J. & Stephens, M. Genotype imputation with thousands of genomes. G3 (Bethesda) 1, 457-470 (2011). https://doi.org:10.1534/g3.111.001198

57 Huang, J. et al. Improved imputation of low-frequency and rare variants using the UK10K haplotype reference panel. Nat Commun 6, 8111 (2015). https://doi.org:10.1038/ncomms9111

58 Mitchell, R., Hemani, G., Dudding, T., Corbin, L., Harrison, S., Paternoster, L. UK Biobank Genetic Data: MRC-IEU Quality Control, version 2 -Datasets -data.bris. data.bris. (2018). https://doi.org:10.5523/bris.1ovaau5sxunp2cv8rcy88688v

59 Mitchell R, E. B., Raistrick CA, Paternoster L, Hemani G, Gaunt TR. MRC IEU UK Biobank GWAS pipeline version 2. (2019). https://doi.org:10.5523/bris.pnoat8cxo0u52p6ynfaekeigi

60 Loh, P. R. et al. Efficient Bayesian mixed-model analysis increases association power in large cohorts. Nat Genet 47, 284–290 (2015). https://doi.org:10.1038/ng.3190

61 Kettunen, J. et al. Genome-wide study for circulating metabolites identifies 62 loci and reveals novel systemic effects of LPA. Nat Commun 7, 11122 (2016). https://doi.org:10.1038/ncomms11122

62 Bruna, G., Mary, S., Debbie, A. L., John, W. L. & George Davey, S. Indicators of socioeconomic position (part 1). Journal of Epidemiology and Community Health 60, 7 (2006). https://doi.org:10.1136/jech.2004.023531

63 Harrell, J. F. E. & SpringerLink. Regression Modeling Strategies: With Applications to Linear Models, Logistic and Ordinal Regression, and Survival Analysis. 1 online resource (XXV, 582 p. 157 illus., 553 illus. in color.) (2015).

64 Lawlor, D. A. Commentary: Two-sample Mendelian randomization: opportunities and challenges. Int J Epidemiol 45, 908–915 (2016). https://doi.org:10.1093/ije/dyw127

65 Day, F. R. et al. Genomic analyses identify hundreds of variants associated with age at menarche and support a role for puberty timing in cancer risk. Nat Genet 49, 834–841 (2017). https://doi.org:10.1038/ng.3841

66 Perry, J. R. et al. Parent-of-origin-specific allelic associations among 106 genomic loci for age at menarche. Nature 514, 92–97 (2014). https://doi.org:10.1038/nature13545

67 Mathieson, I. et al. Genome-wide analysis identifies genetic effects on reproductive success and ongoing natural selection at the <em>FADS</em> locus. bioRxiv, 2020.2005.2019.104455 (2020). https://doi.org:10.1101/2020.05.19.104455

68 Day, F. R. et al. Large-scale genomic analyses link reproductive aging to hypothalamic signaling, breast cancer susceptibility and BRCA1-mediated DNA repair. Nat Genet 47, 1294–1303 (2015). https://doi.org:10.1038/ng.3412

69 Locke, A. E. et al. Genetic studies of body mass index yield new insights for obesity biology. Nature 518, 197–206 (2015). https://doi.org:10.1038/nature14177

70 Howe, L. J. et al. Within-sibship GWAS improve estimates of direct genetic effects. bioRxiv, 2021.2003.2005.433935 (2021). https://doi.org:10.1101/2021.03.05.433935

71 Davies, N. M. et al. Within family Mendelian randomization studies. Hum Mol Genet 28, R170–R179 (2019). https://doi.org:10.1093/hmg/ddz204

72 Morris, T. T., Davies, N. M., Hemani, G. & Smith, G. D. Population phenomena inflate genetic associations of complex social traits. Sci Adv 6, eaay0328 (2020). https://doi.org:10.1126/sciadv.aay0328

73 Sanderson, E., Richardson, T. G., Hemani, G. & Davey Smith, G. The use of negative control outcomes in Mendelian randomization to detect potential population stratification. Int J Epidemiol (2021). https://doi.org:10.1093/ije/dyaa288

74 Zheng, J. et al. Recent Developments in Mendelian Randomization Studies. Curr Epidemiol Rep 4, 330–345 (2017). https://doi.org:10.1007/s40471-017-0128-6

