## Supplementary figures for "From menarche to menopause: the impact of reproductive factors on the metabolic profile of over 65,000 women"

Suppl Fig 1. Multivariable regression (red) and Mendelian randomization (black) estimates for the relation between older age at menarche and metabolic measures among females.
Suppl Fig 2. Multivariable regression estimates for the relation between older age at menarche and metabolic measures among females (comparing different model adjustments)
Suppl Fig 3. Multivariable regression estimates for the relation between age at menarche (<13 years, 13-14 years, >14 years) and metabolic measures among females
Suppl Fig 4. Multivariable regression estimates for the relation between age at menarche (restricted cubic splines with knots placed at ages 11, 13, and 15) and metabolic measures among females
Suppl Fig 5. Univariable and multivariable Mendelian randomization estimates for the relation between older age at menarche and NMR metabolomics measures among females
Suppl Fig 6. Univariable and multivariable Mendelian randomization estimates for the relation between older age at menarche and clinical chemistry biomarkers among females
Suppl Fig 7. Multivariable regression (red) and Mendelian randomization (black) estimates for the relation between higher parity and metabolic measures among females.
Suppl Fig 8. Multivariable regression estimates for the relation between higher parity and metabolic measures among females (comparing different model adjustments)
Suppl Fig 9. Multivariable regression estimates for the relation between parity (0, 1, 2, 3+) and metabolic measures among females
Suppl Fig 10. Multivariable regression estimates for the relation between parity (restricted cubic splines with knots placed at 1 2 and 3 births) and metabolic measures among females
Suppl Fig 11 Multivariable regression (red) and Mendelian randomization (black) estimates for the relation between older age at natural menopause and metabolic measures among females
Suppl Fig 12. Multivariable regression estimates for the relation between older age at natural menopause and metabolic measures among females (comparing different model adjustments)
Suppl Fig 13. Multivariable regression estimates for the relation between age at menopause (<49, 49-50, 51-53, >53) and metabolic measures among females
Suppl Fig 14. Multivariable regression estimates for the relation between age at menopause (restricted cubic splines with knots placed at ages 40, 49, 52, and 56) and metabolic measures among females
Suppl Fig 15. Age-combined and age-stratified estimates for the association between age at natural menarche and clinical chemistry biomarkers using multivariable regression (red) and Mendelian randomization (blue) restricted to women with data on age at menopause or Mendelian randomization (black) using data from all women.

Suppl Fig 16A. Age-combined and age-stratified estimates for the association between age at natural menarche and clinical chemistry biomarkers excluding users of statins at baseline estimated using multivariable regression (red) and Mendelian randomization (blue) restricted to women with data on age at menopause or Mendelian randomization (black) using data from all women.

Suppl Fig 16B. Age-combined and age-stratified estimates for the association between age at natural menarche and clinical chemistry biomarkers excluding users of hormone replacement therapy (HRT) at baseline estimated using multivariable regression (red) and Mendelian randomization (blue) restricted to women with data on age at menopause or Mendelian randomization (black) using data from all women.

Suppl Fig 17. Mendelian randomization estimates for the relation between reproductive markers and conventional biomarkers among unrelated individuals and within siblings.

Suppl Fig 18. Mendelian randomization estimates for the relation between older age at menarche and metabolic measures among females (comparing different Mendelian randomization methods)

Suppl Fig 19. Mendelian randomization estimates for the relation between higher parity and metabolic measures among females (comparing different Mendelian randomization methods)

Suppl Fig 20. Mendelian randomization estimates for the relation between older age at natural menopause and metabolic measures among females (compare different Mendelian randomization methods)

Suppl Fig 21. Mendelian randomization estimates for the relation between older age at menarche and metabolic measures among females (comparing different SNP sets)

Suppl Fig 22. Mendelian randomization estimates for the relation between older age at natural menopause and metabolic measures among females (comparing different SNP sets)

Suppl Fig 1. Multivariable regression (red) and Mendelian randomization (black) estimates for the relation between older age at menarche and metabolic measures among females. Footnote: MV=multivariable; MR=Mendelian Randomisation. Multivariable regression model is based on adjustments of age at baseline, education, and body composition at age 10.

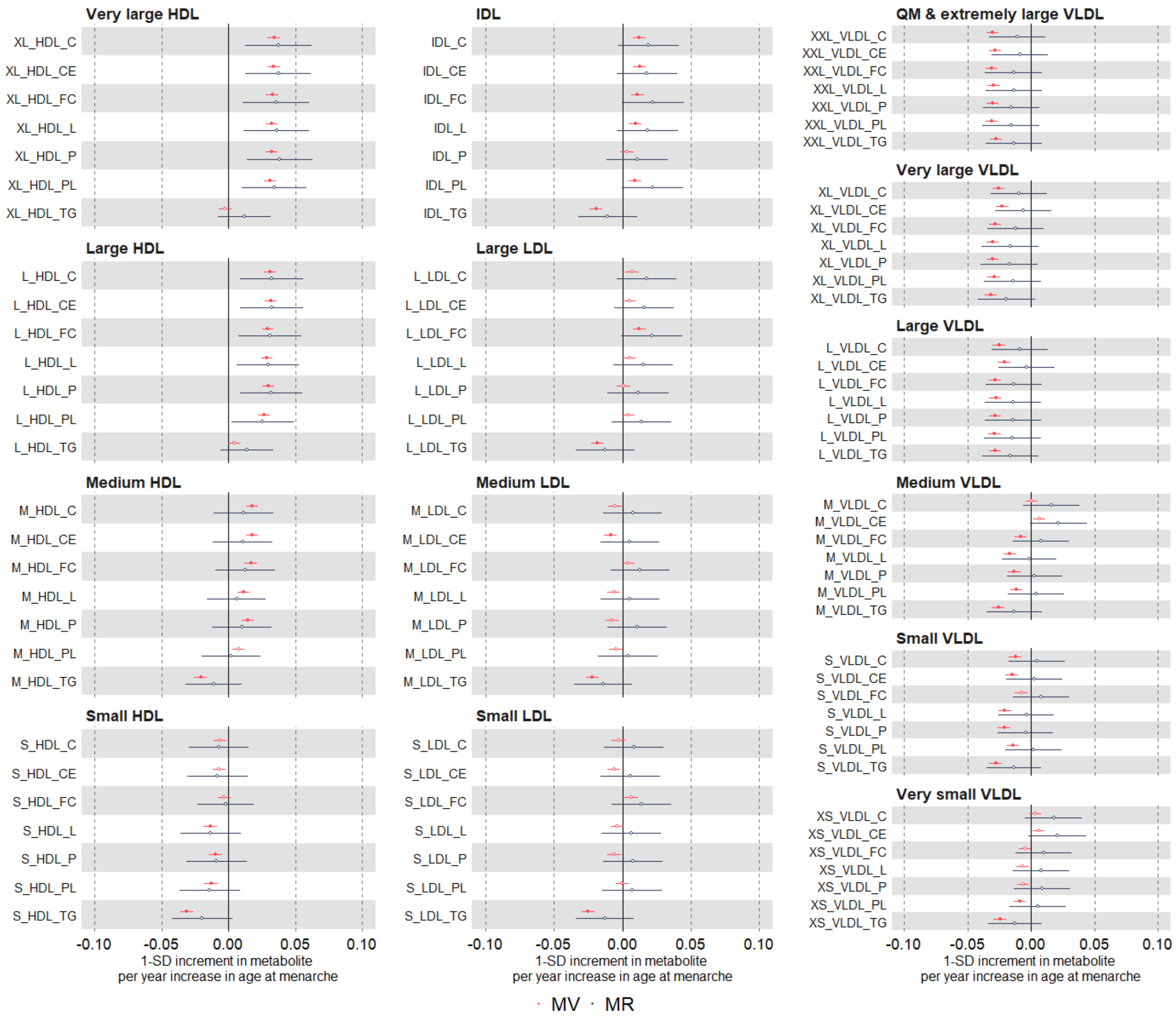

Suppl Fig 1. Continued

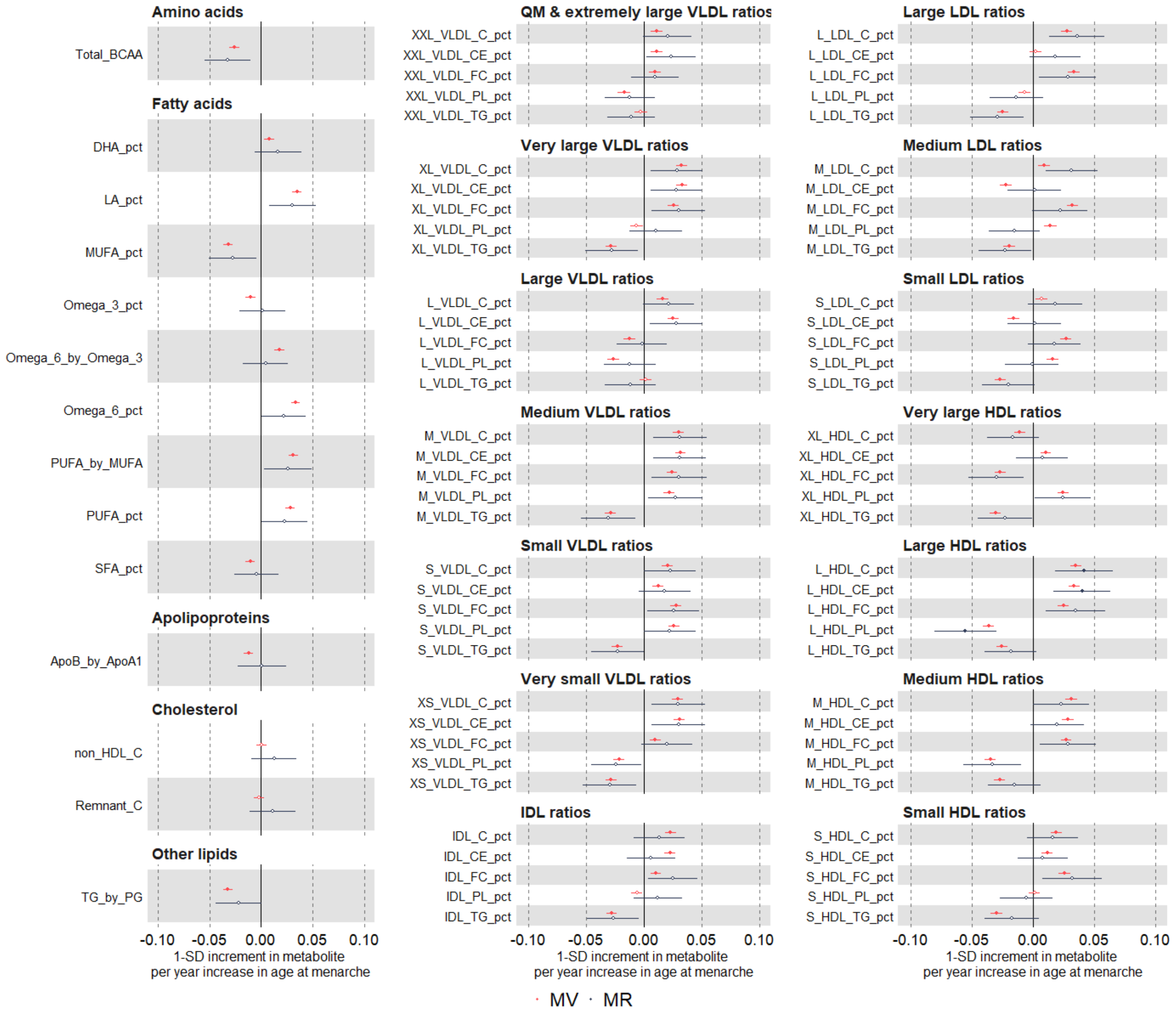

Suppl Fig 2. Multivariable regression estimates for the relation between older age at menarche and metabolic measures among females (comparing different model adjustments). Footnote: model 1 (unadjusted) (red); model 2 (main model, age at baseline, education, and body composition at age 10) (green); model 3 (additionally adjusted for BMI, smoking ad alcohol status at baseline) (black).

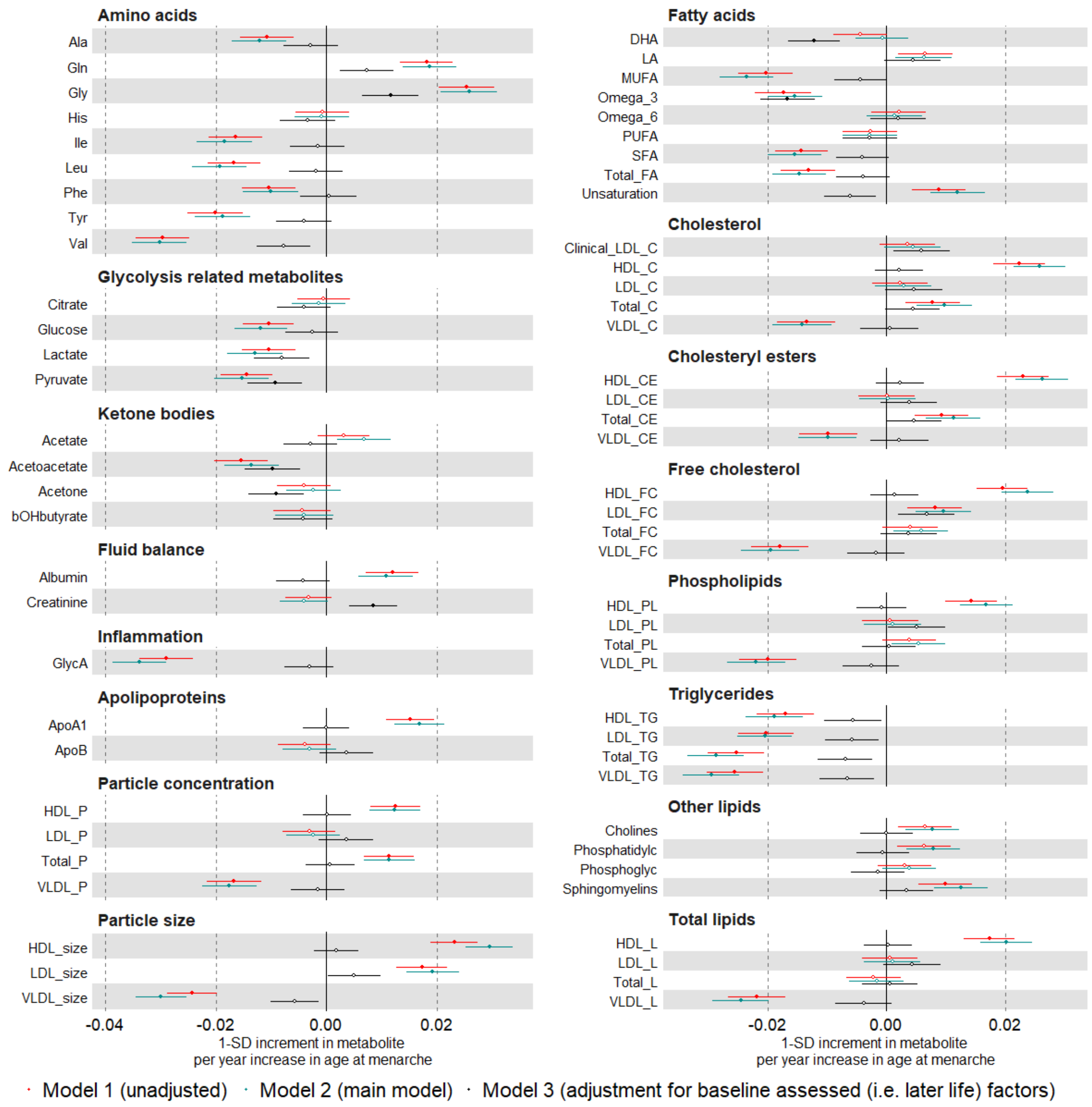

Suppl Fig 2. Continued

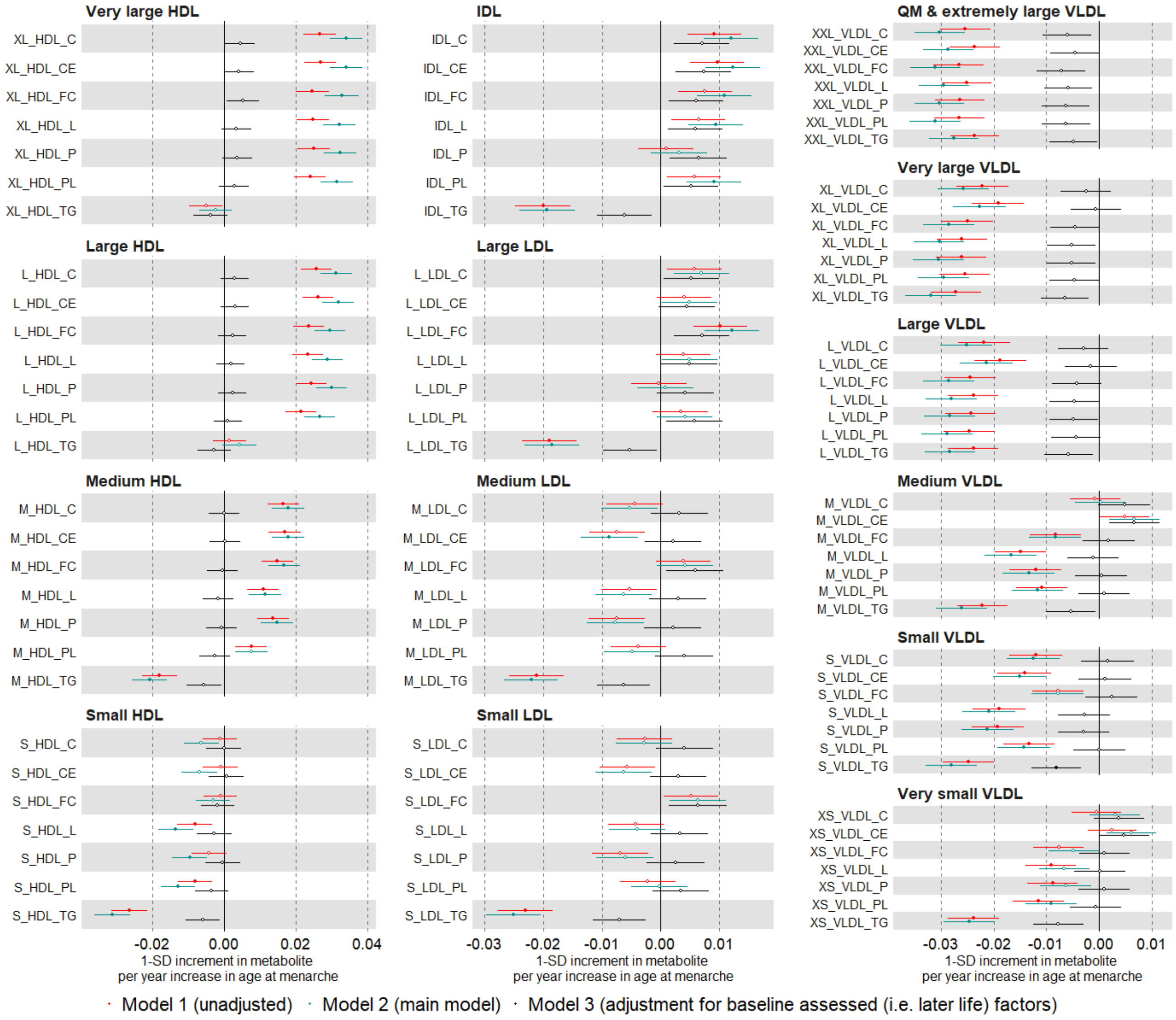

Suppl Fig 2. Continued

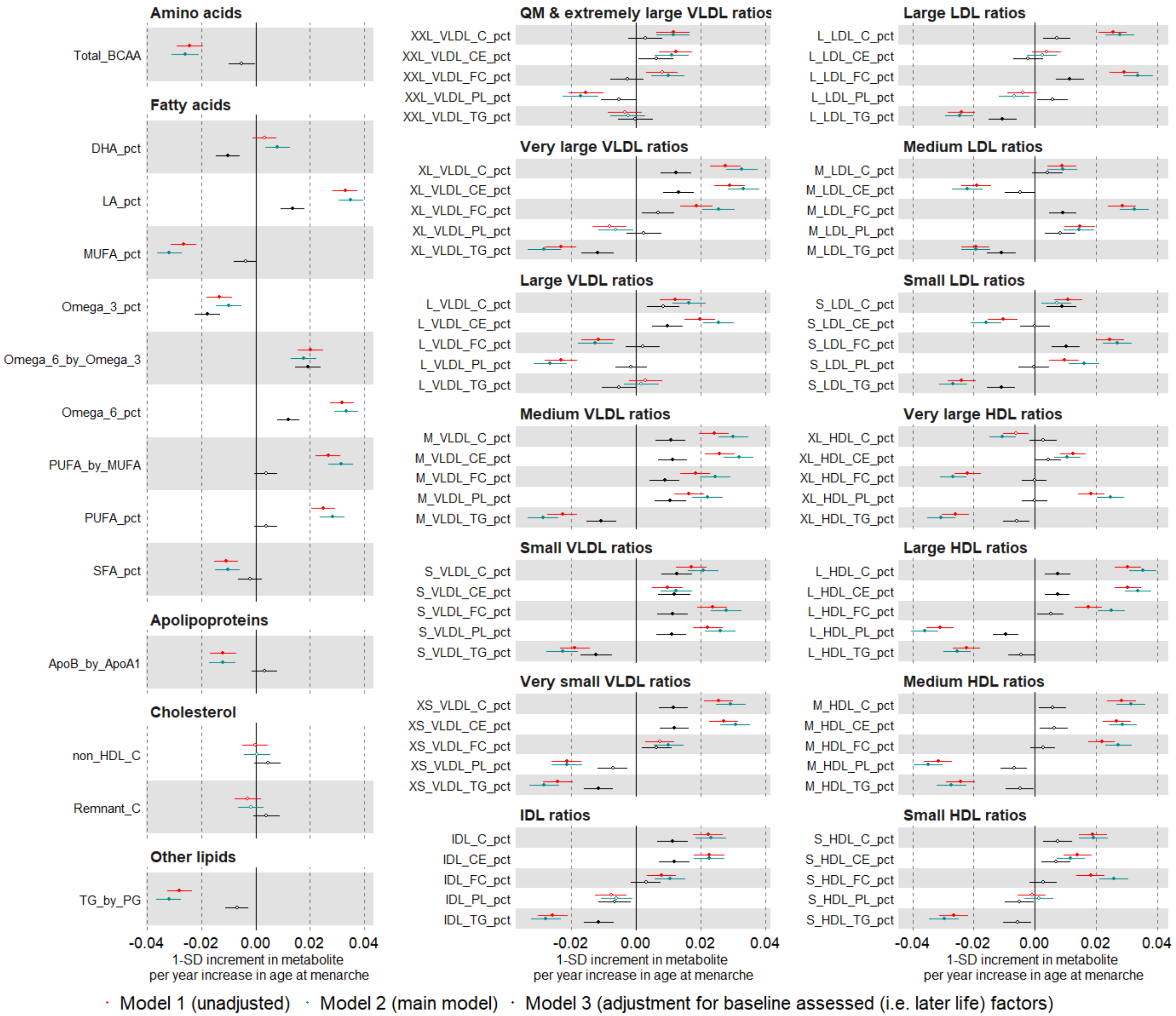

Suppl Fig 3. Multivariable regression estimates for the relation between age at menarche (categorised: <13 years (reference), 13-14 years, >14 years) and metabolic measures among females. Model adjusted for age at baseline, education, and body composition at age 10.

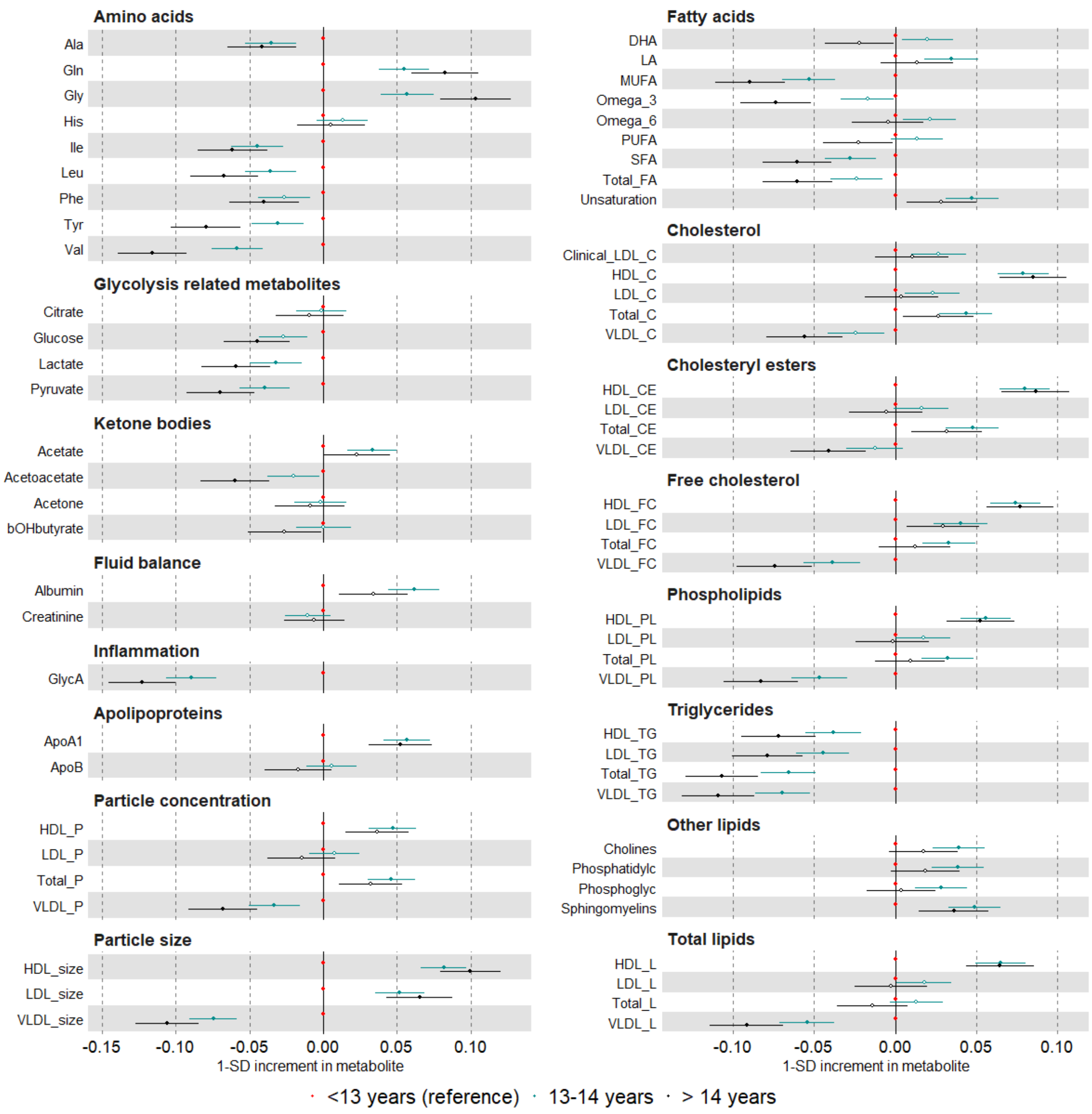

Suppl Fig 3. Continued

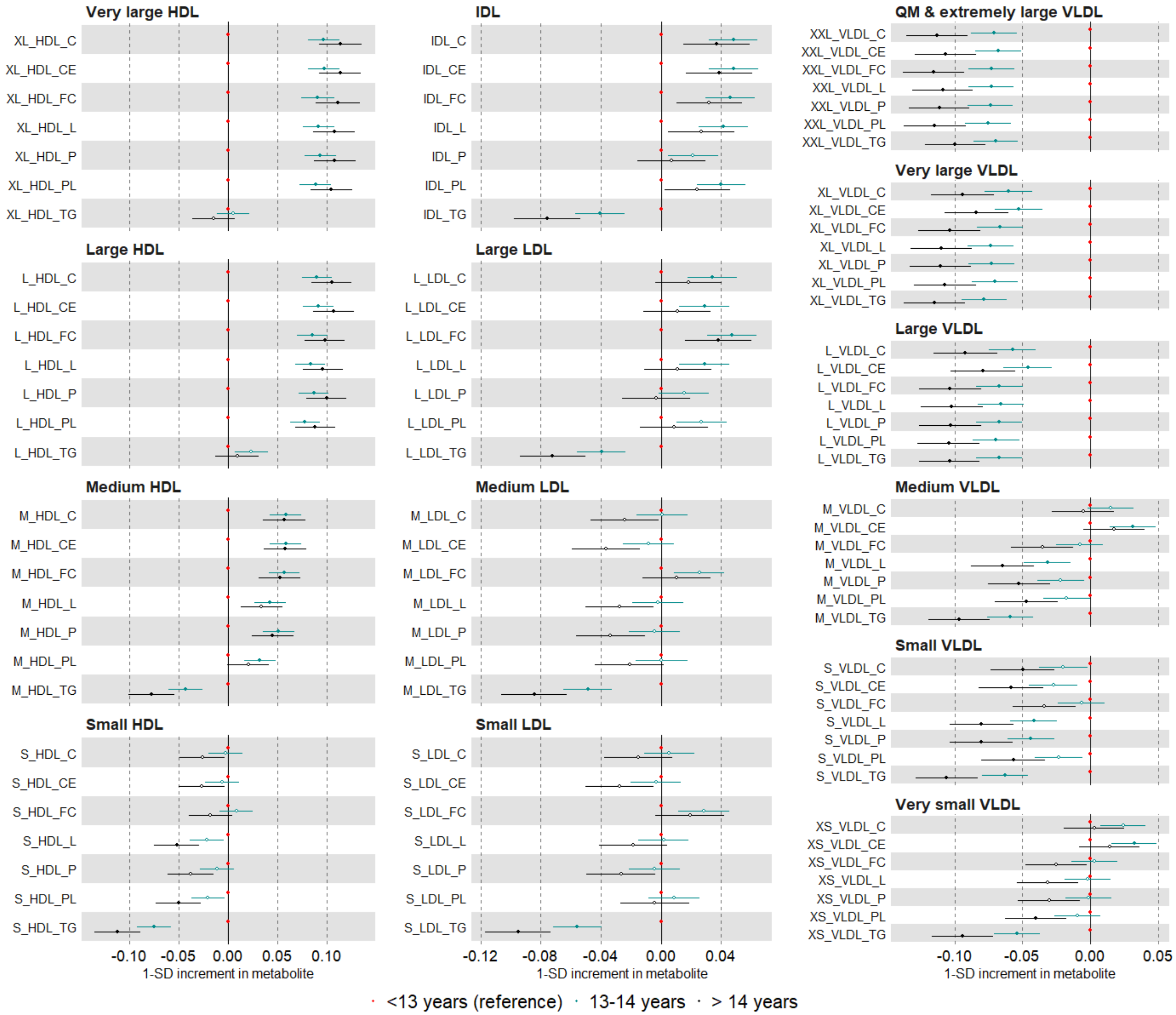

Suppl Fig 3. Continued

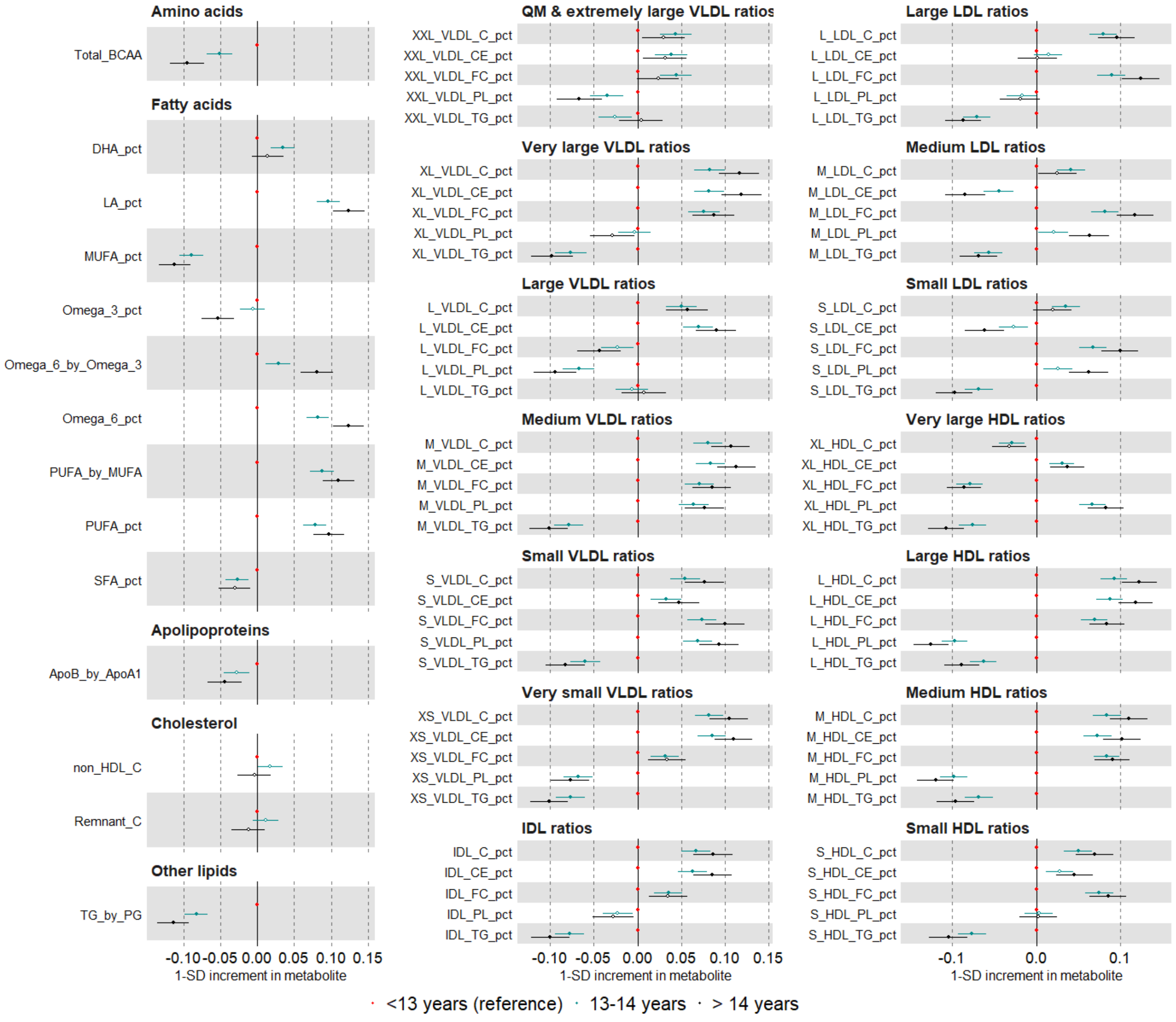

Suppl Fig 4. Multivariable regression estimates for the relation between age at menarche (restricted cubic splines with knots placed at ages 11, 13, and 15) and metabolic measures among females (Mean predicted outcome levels at different menarche ages for a women who is 60 years old, had an average body size at age 10 and is educated to college or university level, restricted cubic splines plotted in blue and a linear association from our main model in red)

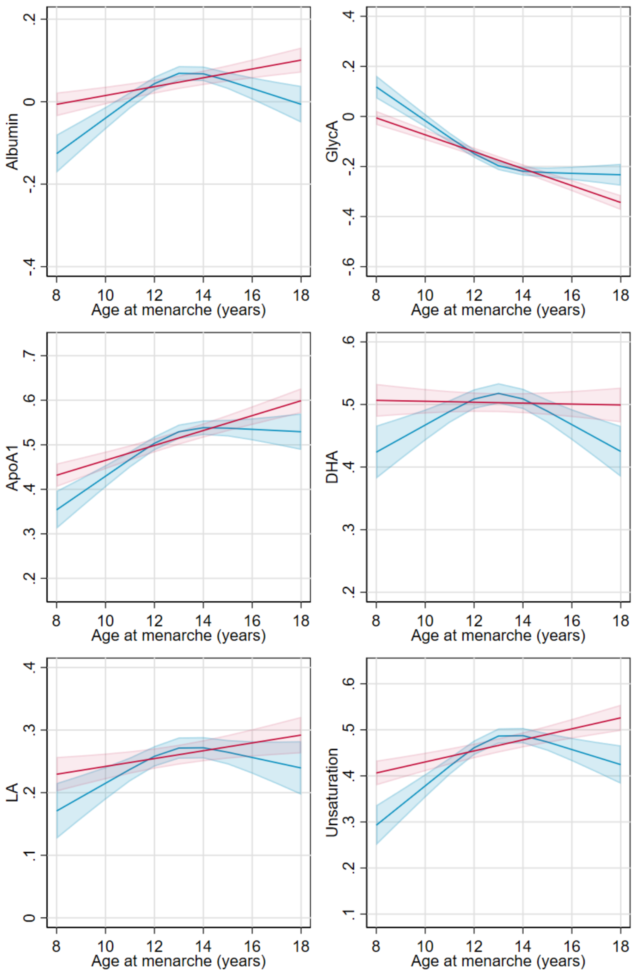

Suppl Fig 4.continued

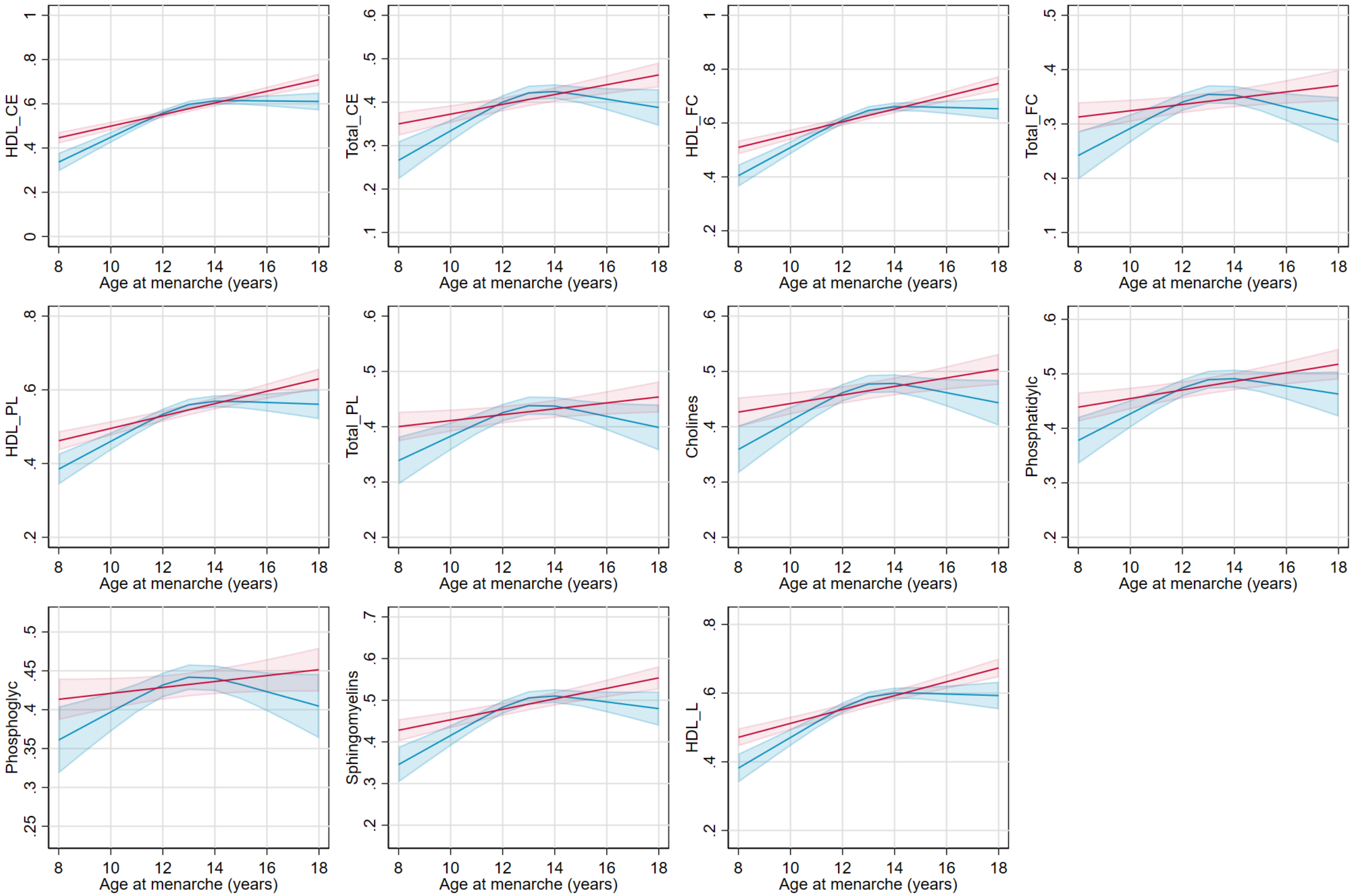

Suppl Fig 5. Univariable and multivariable Mendelian randomization estimates for the relation between older age at menarche and NMR metabolomics measures among females

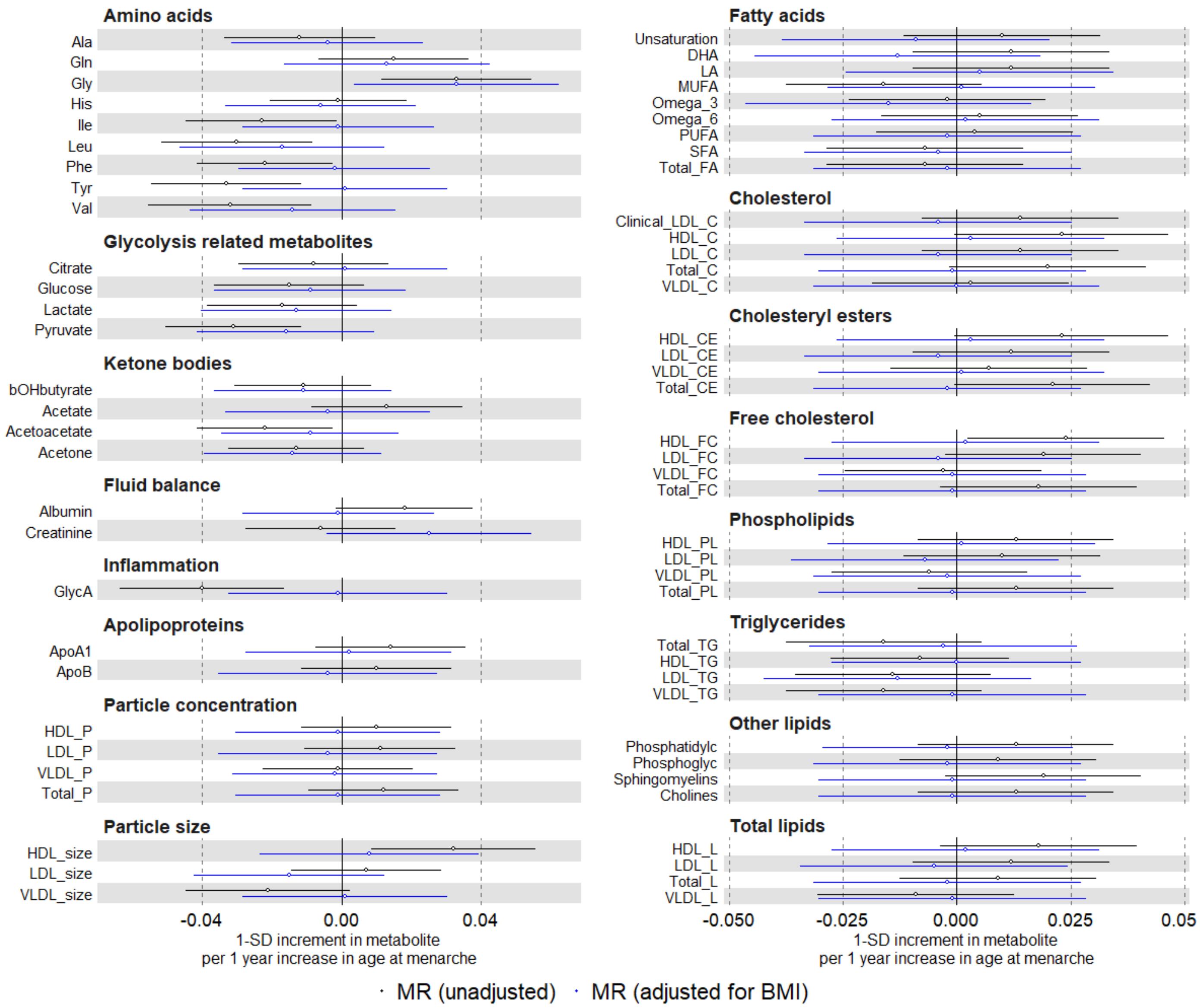

Suppl Fig 5. Continued

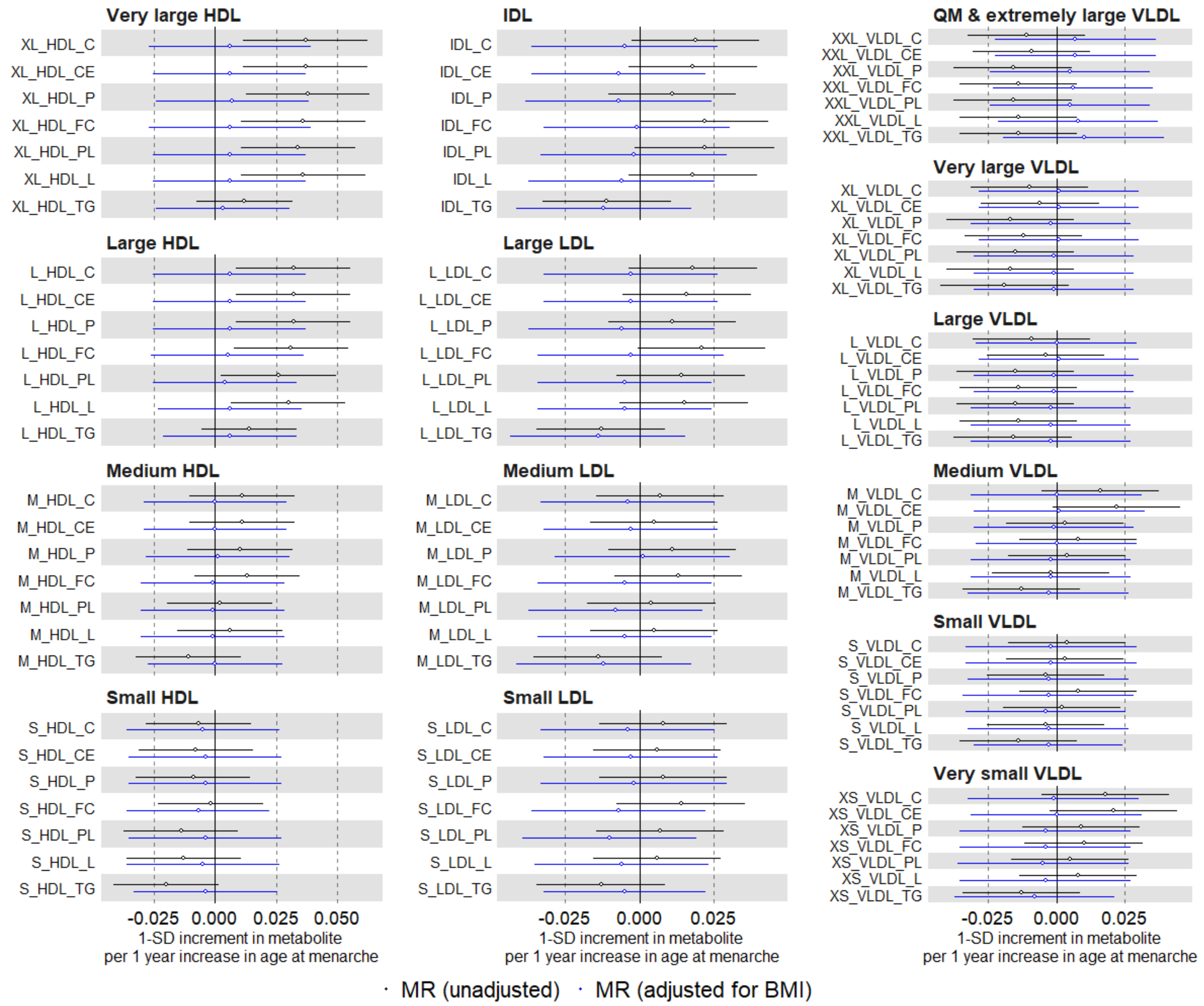

Suppl Fig 5. continued

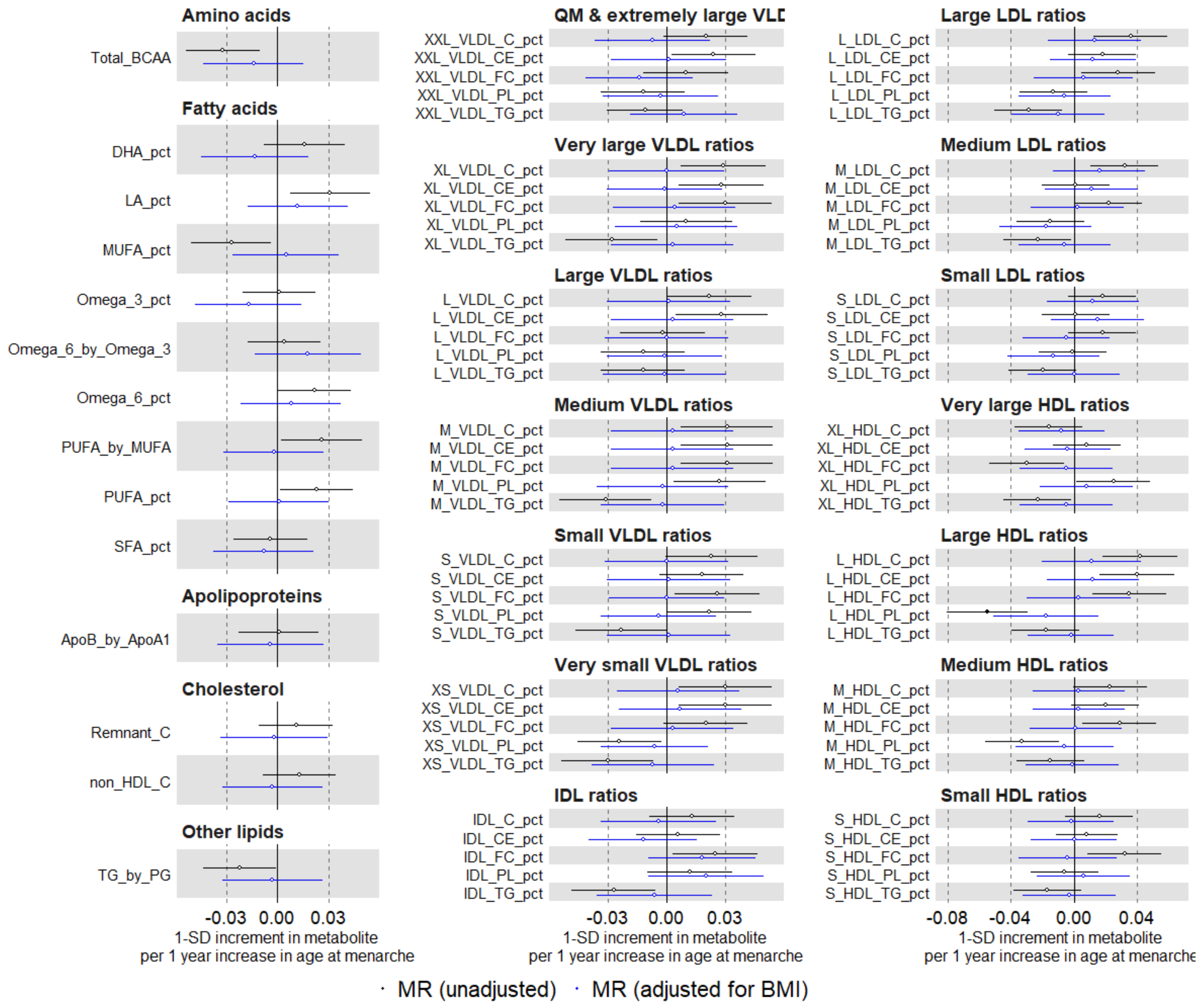

Suppl Fig 6. Univariable and multivariable Mendelian randomization estimates for the relation between older age at menarche and clinical chemistry biomarkers among females

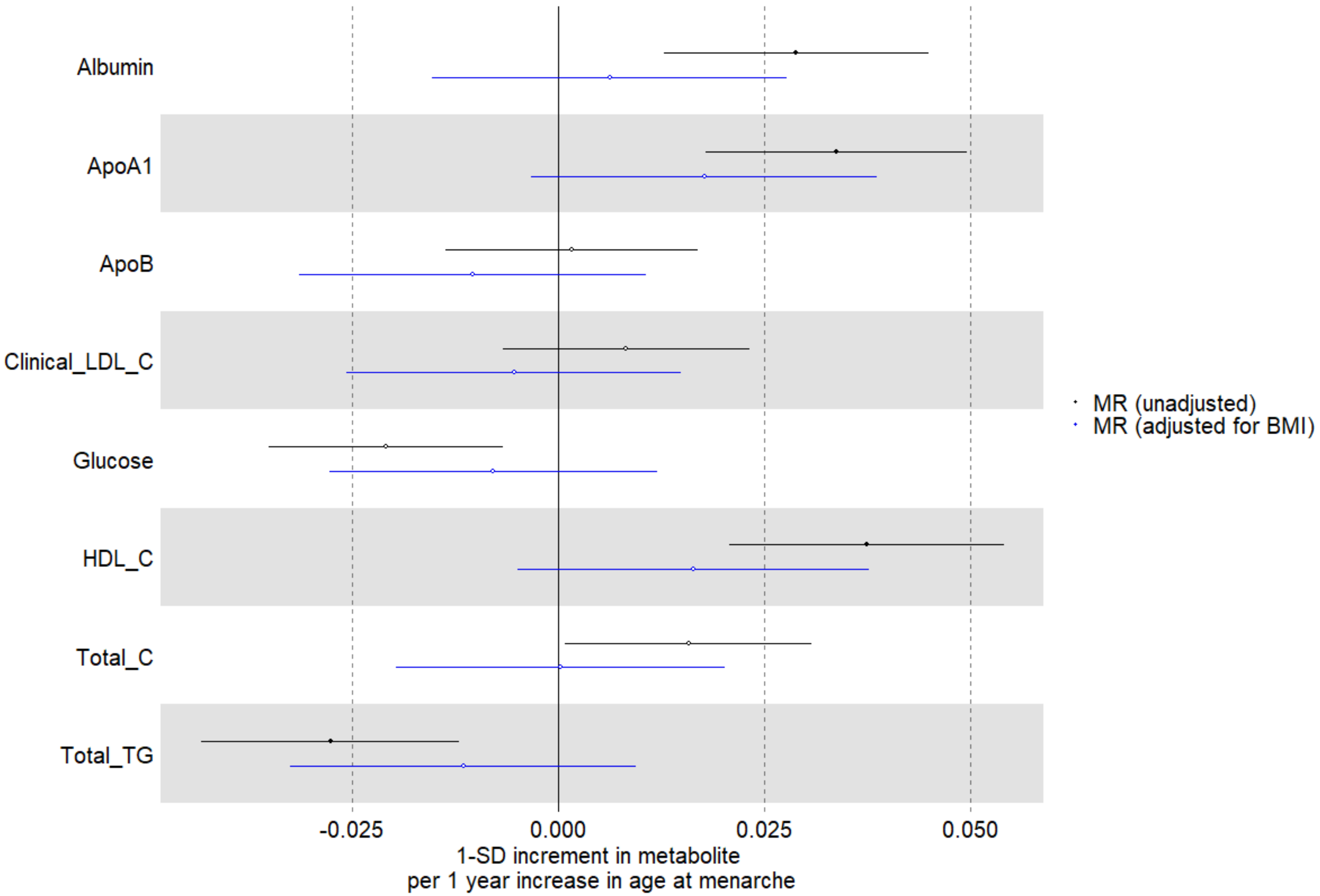

Suppl Fig 7. Multivariable regression (red) and Mendelian randomization (black) estimates for the relation between higher parity and metabolic measures among females. Footnote: MV=multivariable; MR=Mendelian Randomisation. Multivariable regression model is based on adjustments of age at baseline, education, and body composition at age 10.

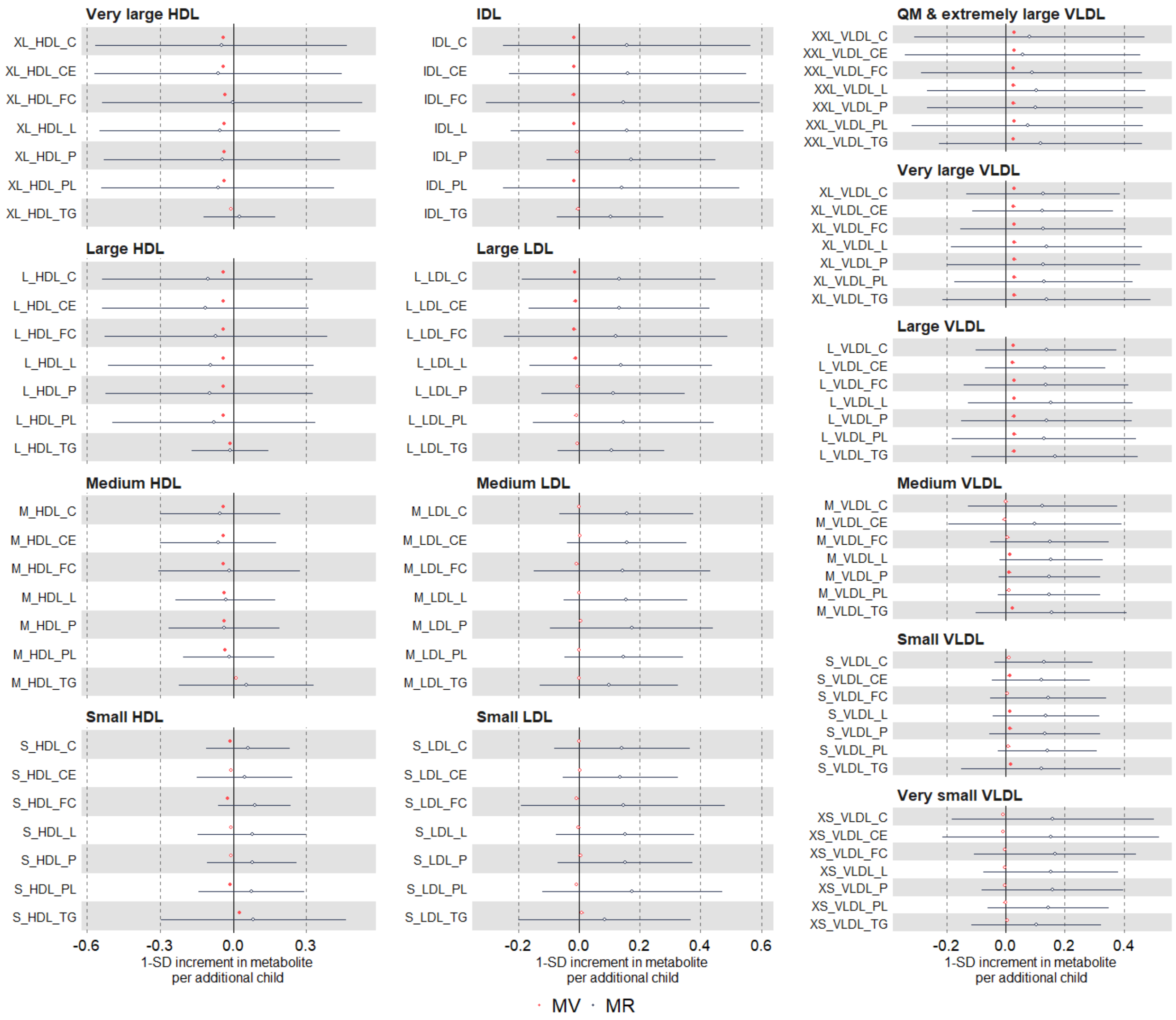

Suppl Fig 7 Continued

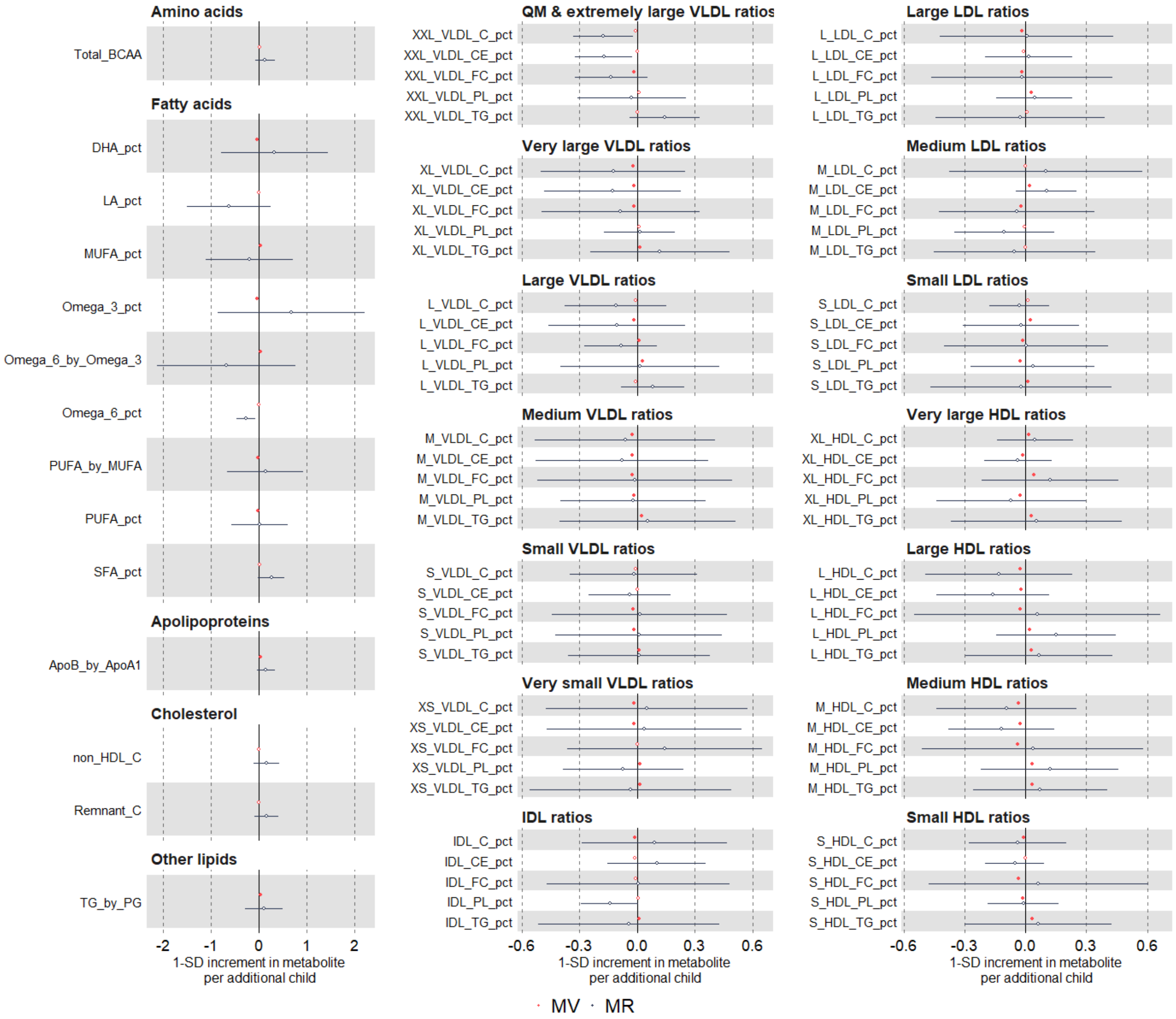

Suppl Fig 8. Multivariable regression estimates for the relation between higher parity and metabolic measures among females (comparing different model adjustments). Footnote: model 1 (unadjusted) (red); model 2 (main model, age at baseline, education, and body composition at age 10) (green); model 3 (additionally adjusted for BMI, smoking ad alcohol status at baseline) (black).

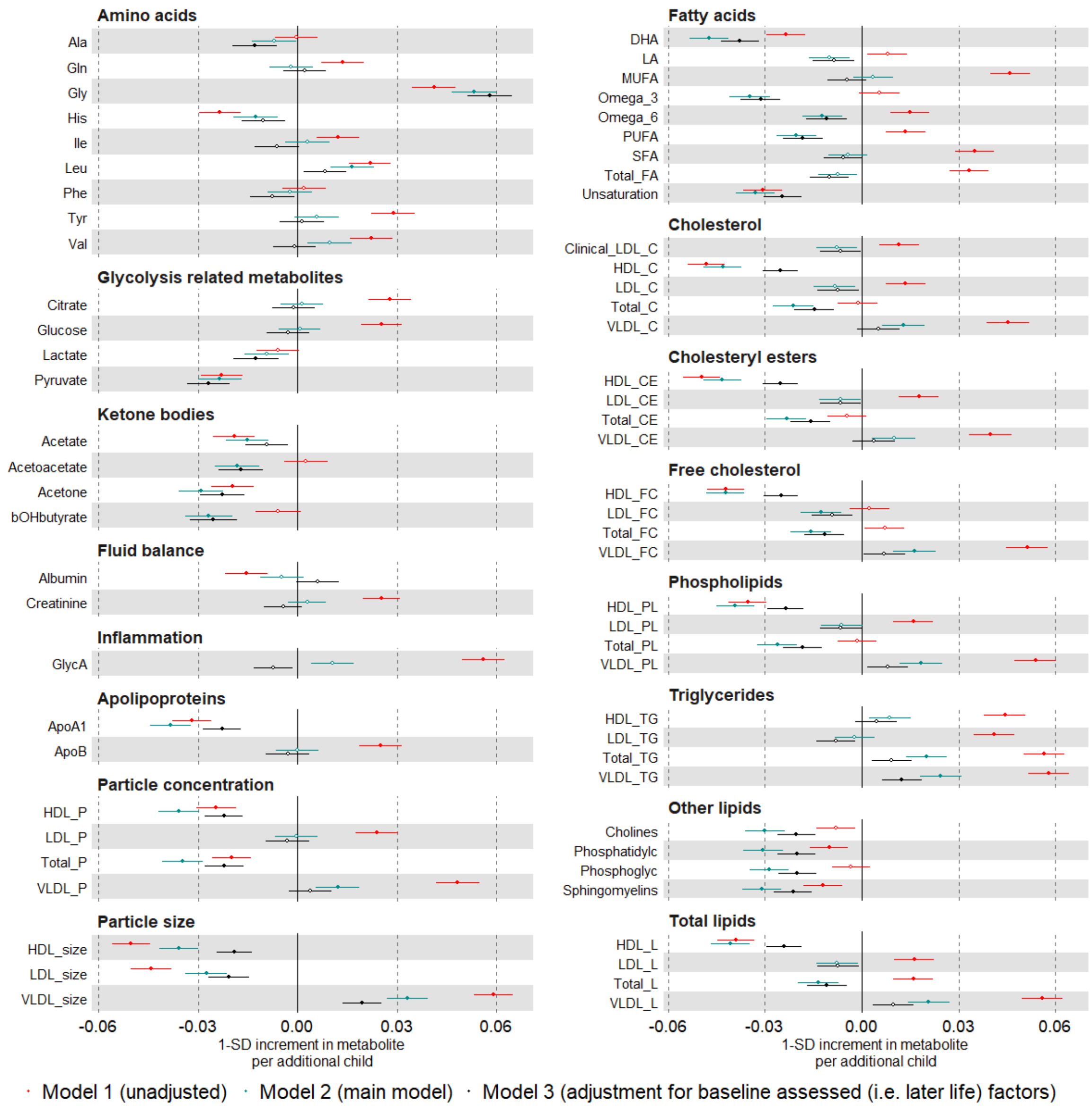

Suppl Fig 8. Continued

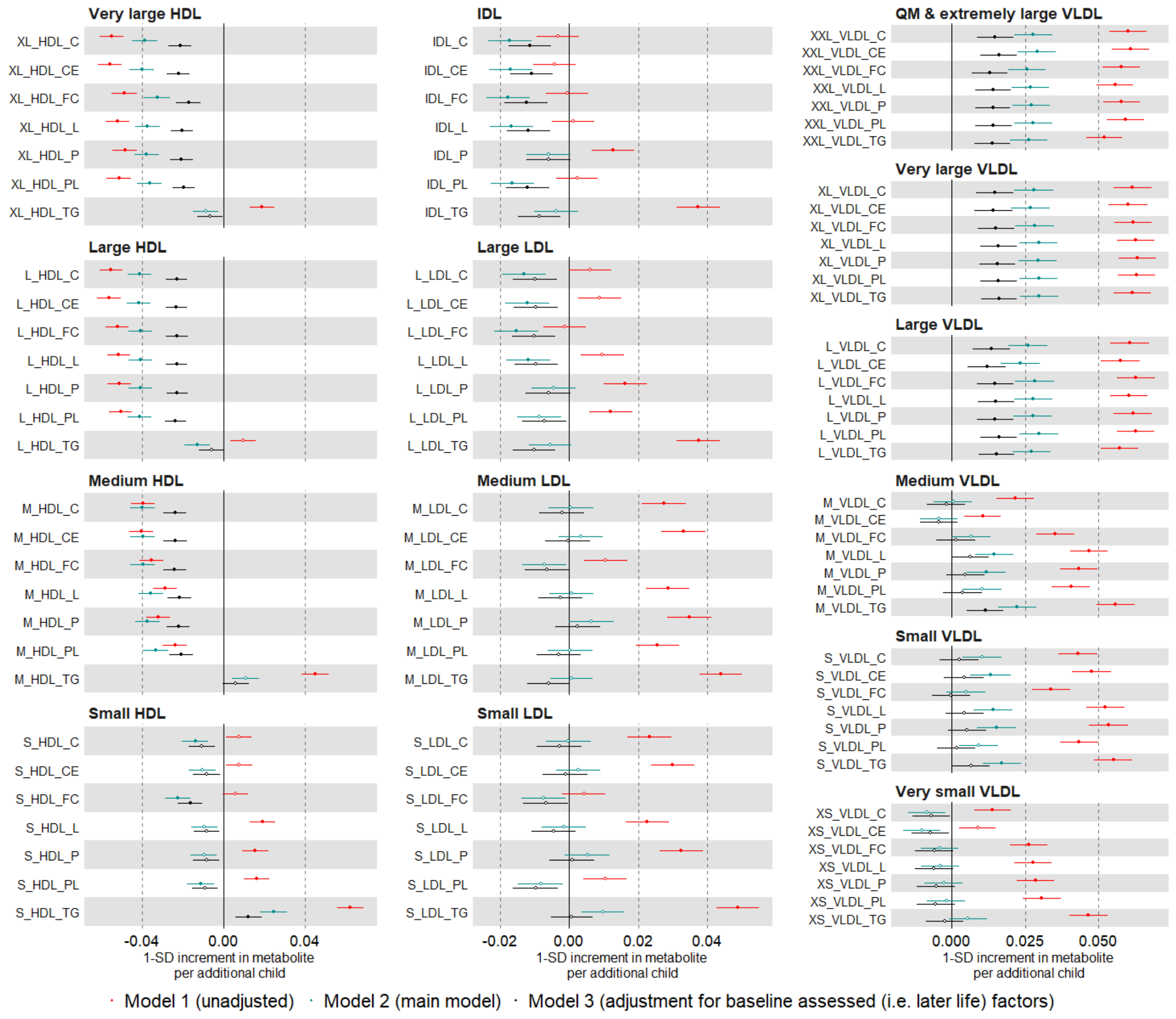

Suppl Fig 8. continued

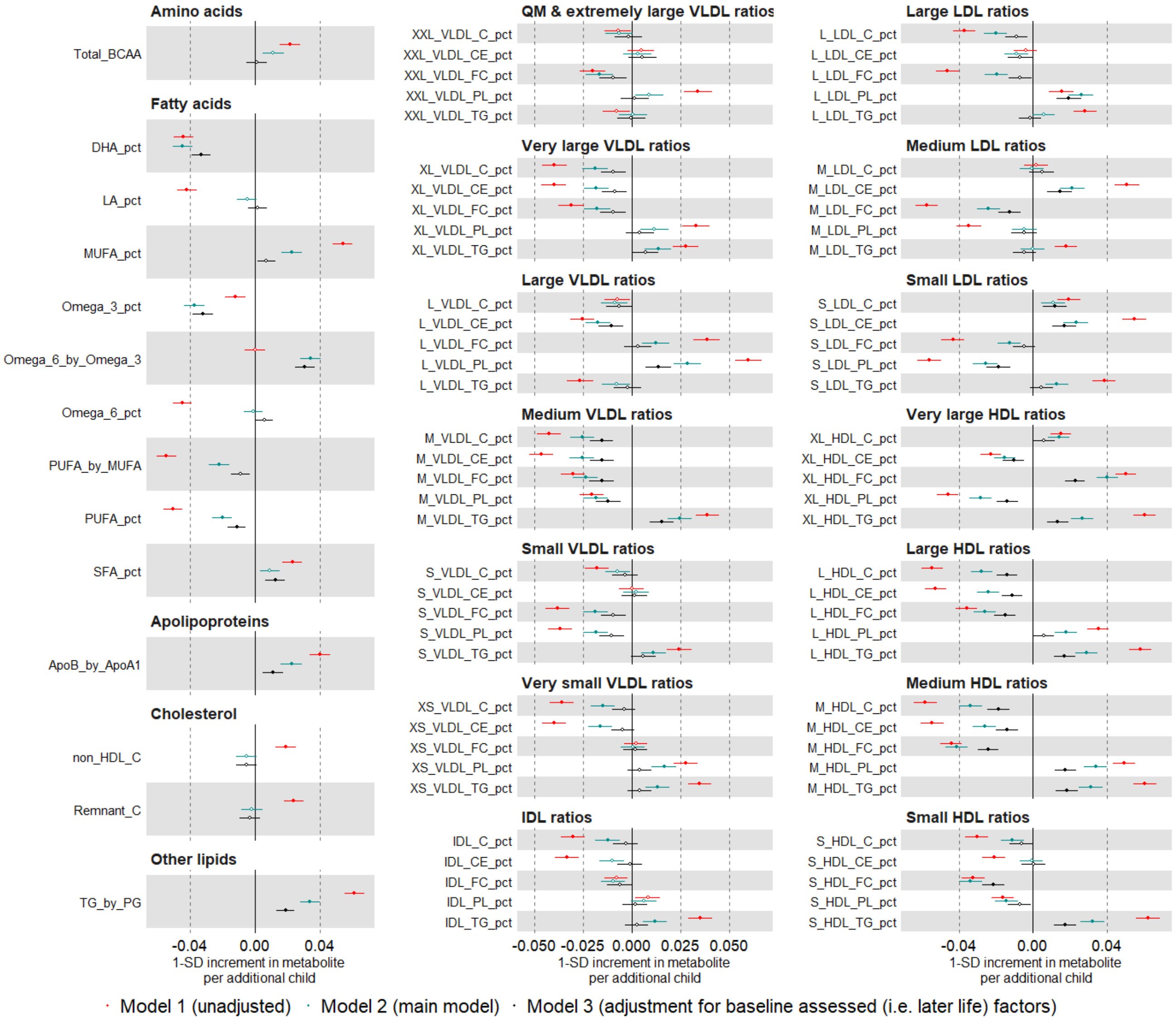

Suppl Fig 9. Multivariable regression estimates for the relation between parity (categorised: 0 (reference), 1, 2, 3+) and metabolic measures among females. Model adjusted for age at baseline, education, and body composition at age 10.

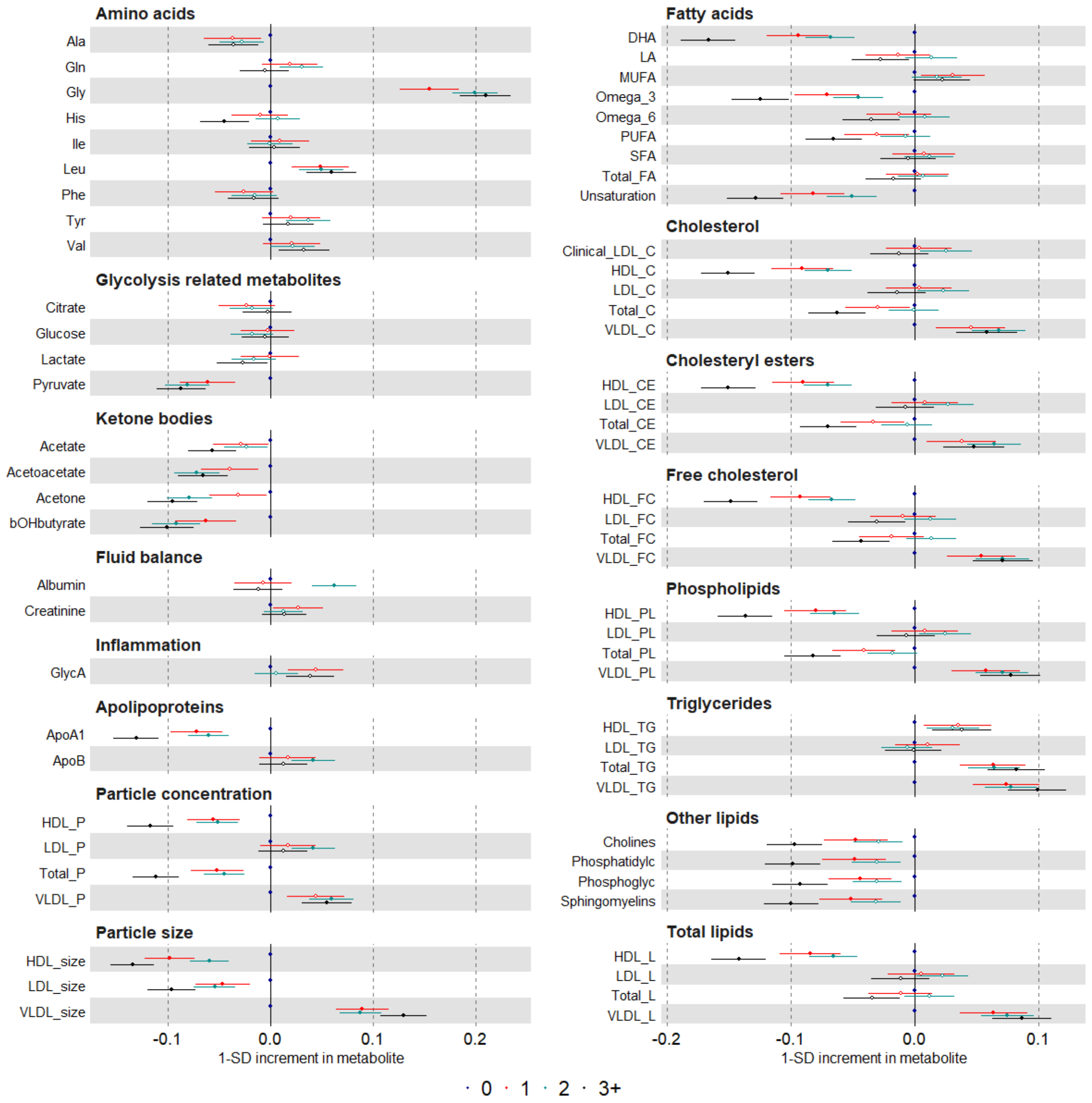

Suppl Fig 9. Continued

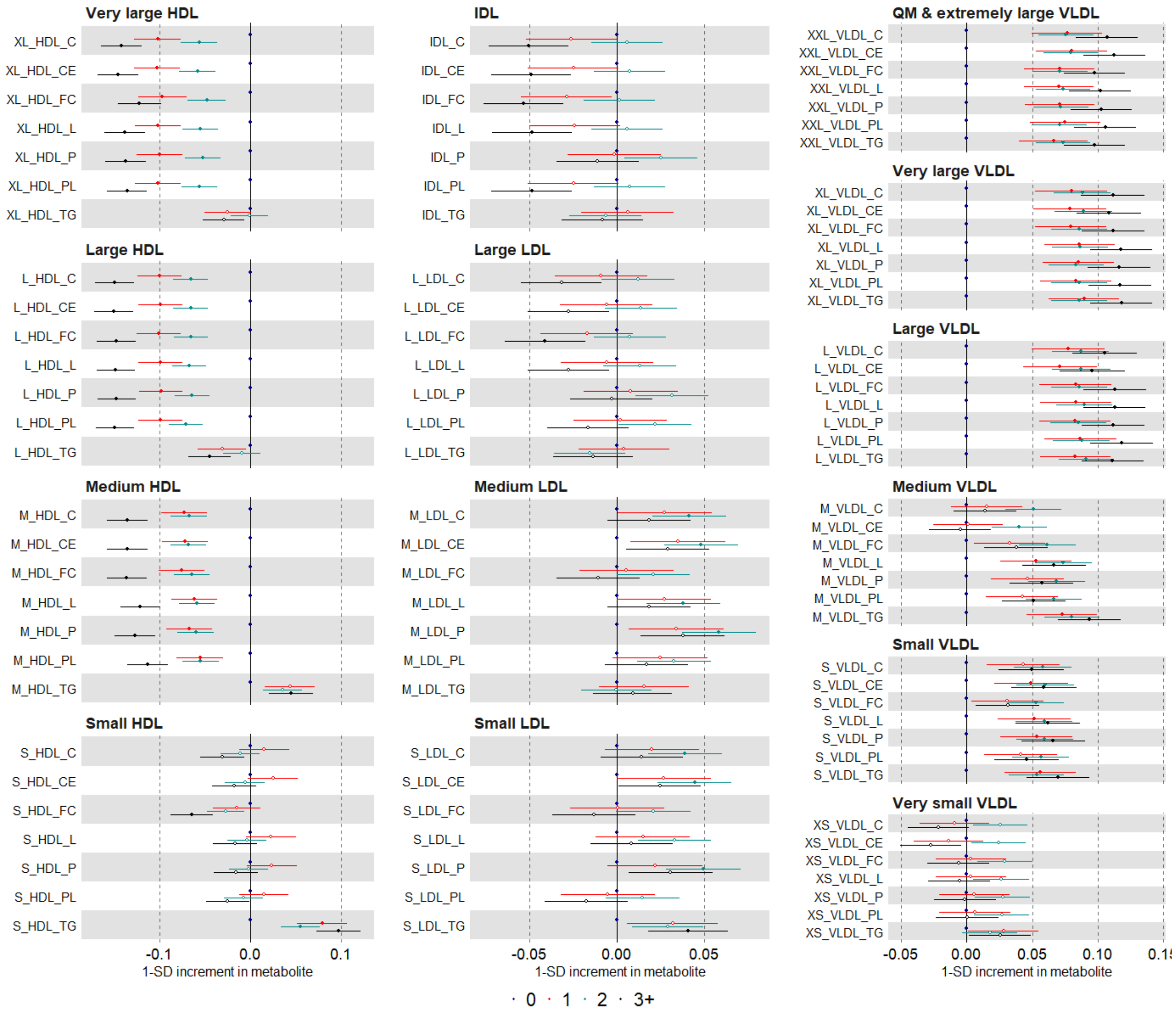

Suppl Fig 9. Continued

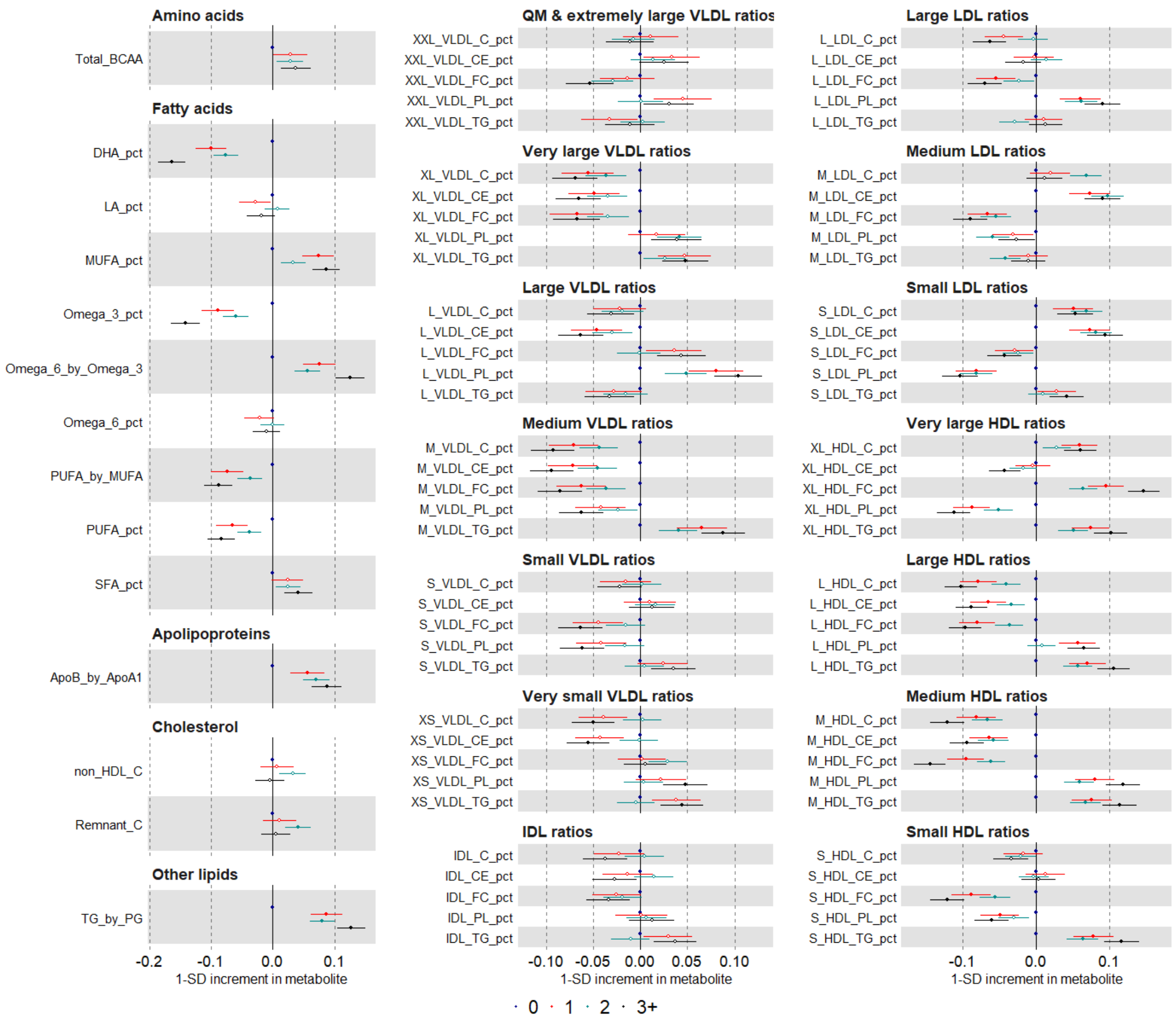

Suppl Fig 10. Multivariable regression estimates for the relation between parity (restricted cubic splines with knots placed at 1 2 and 3) and metabolic measures among females (Mean predicted outcome levels at different levels of parity for a women who is 60 years old, had an average body size at age 10 and is educated to college or university level)

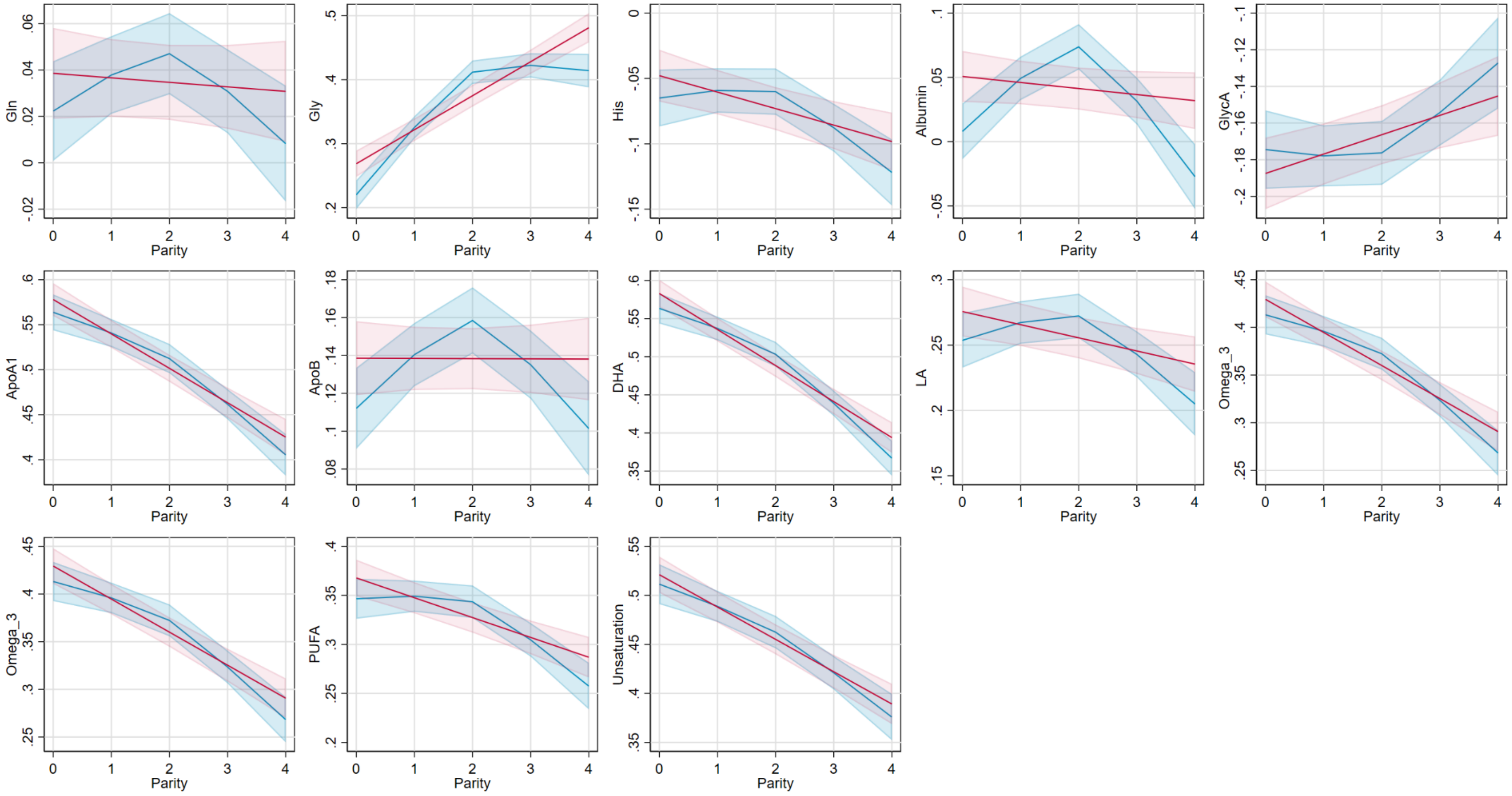

Suppl Fig 10 .continued

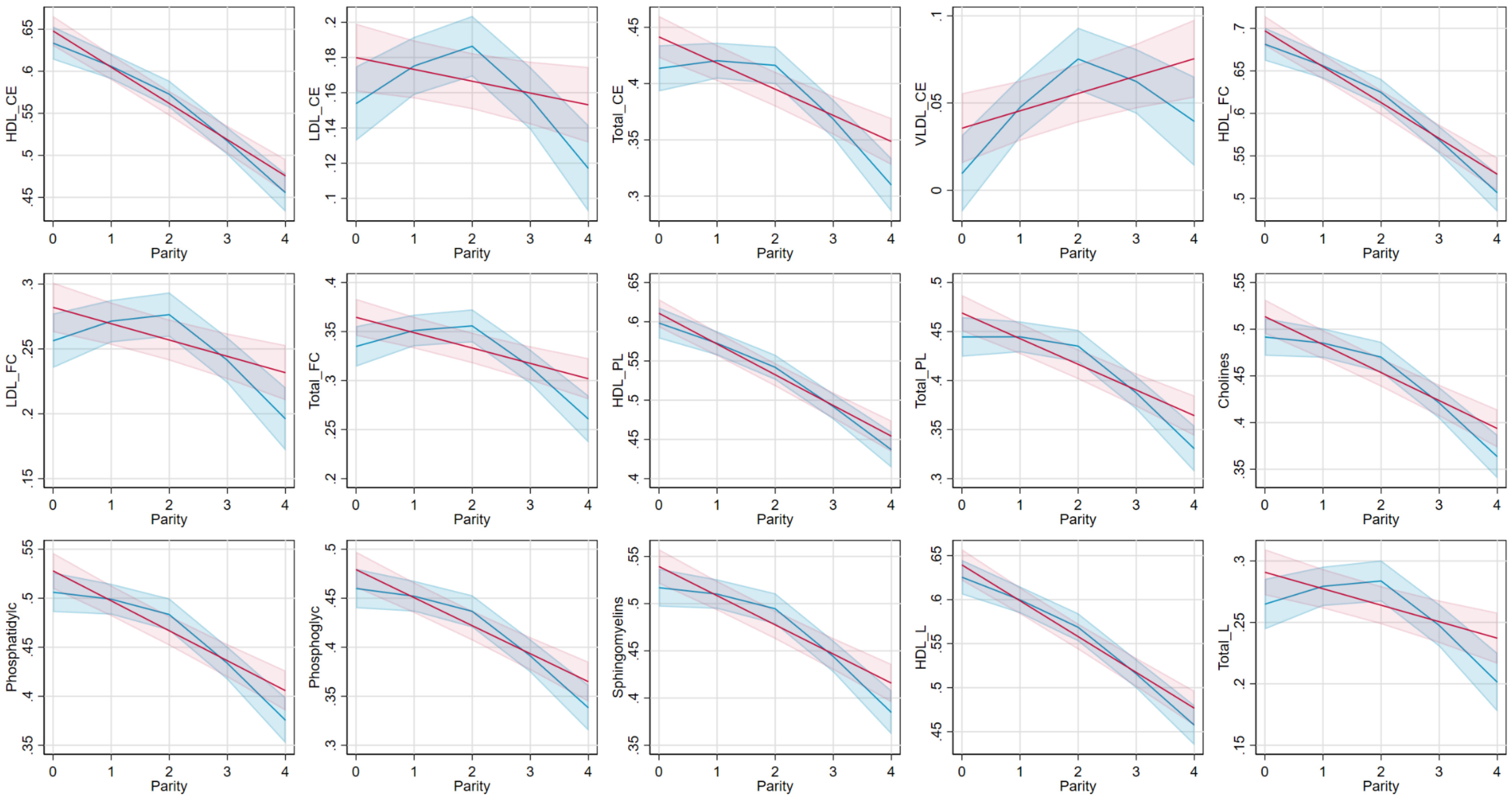

Suppl Fig 11. Multivariable regression (red) and Mendelian randomization (black) estimates for the relation between older age at natural menopause and metabolic measures among females. Footnote: MV=multivariable; MR=Mendelian Randomisation. Multivariable regression model is based on adjustments of age at baseline, education, and body composition at age 10.

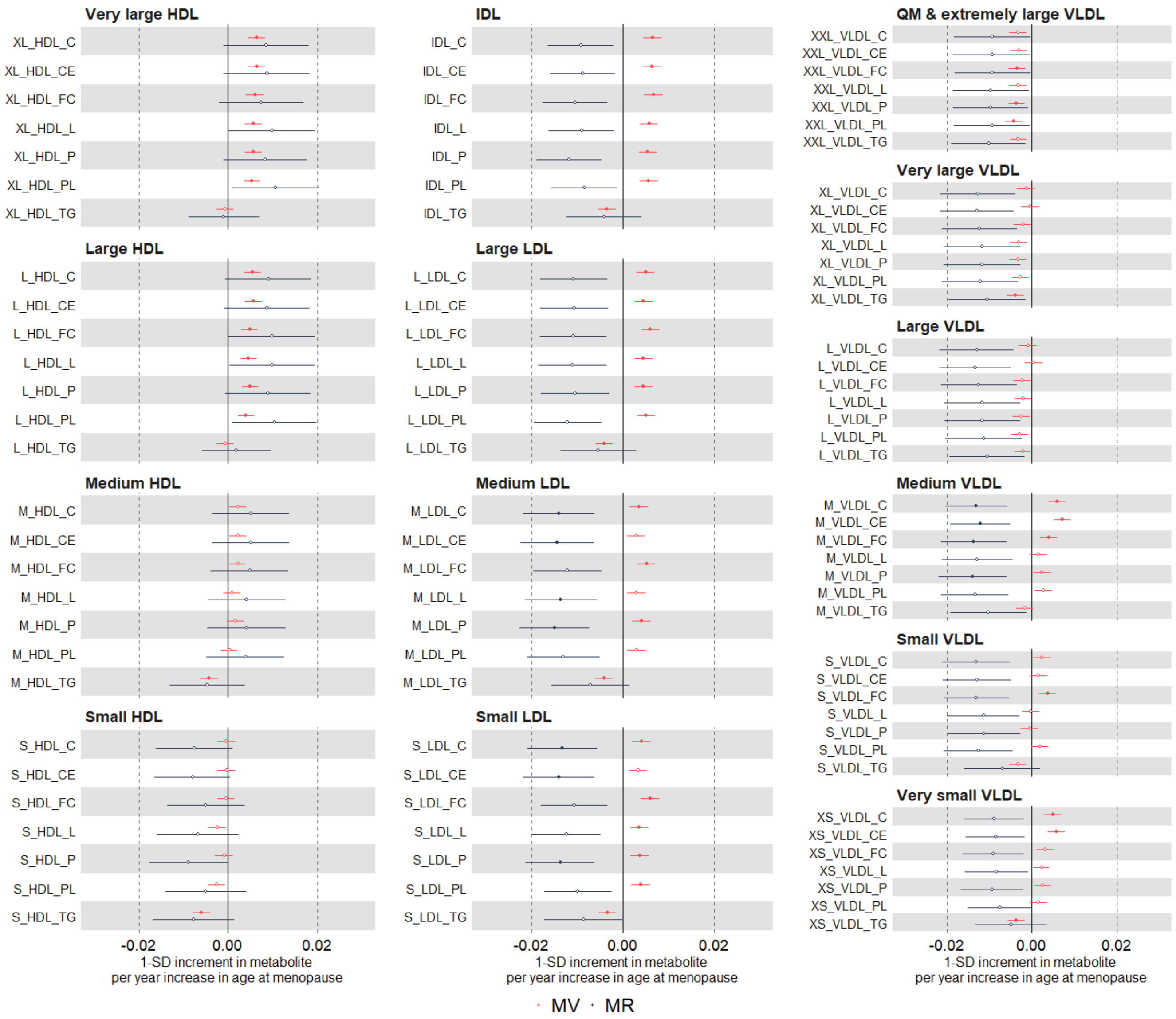

Suppl Fig 11. Continued

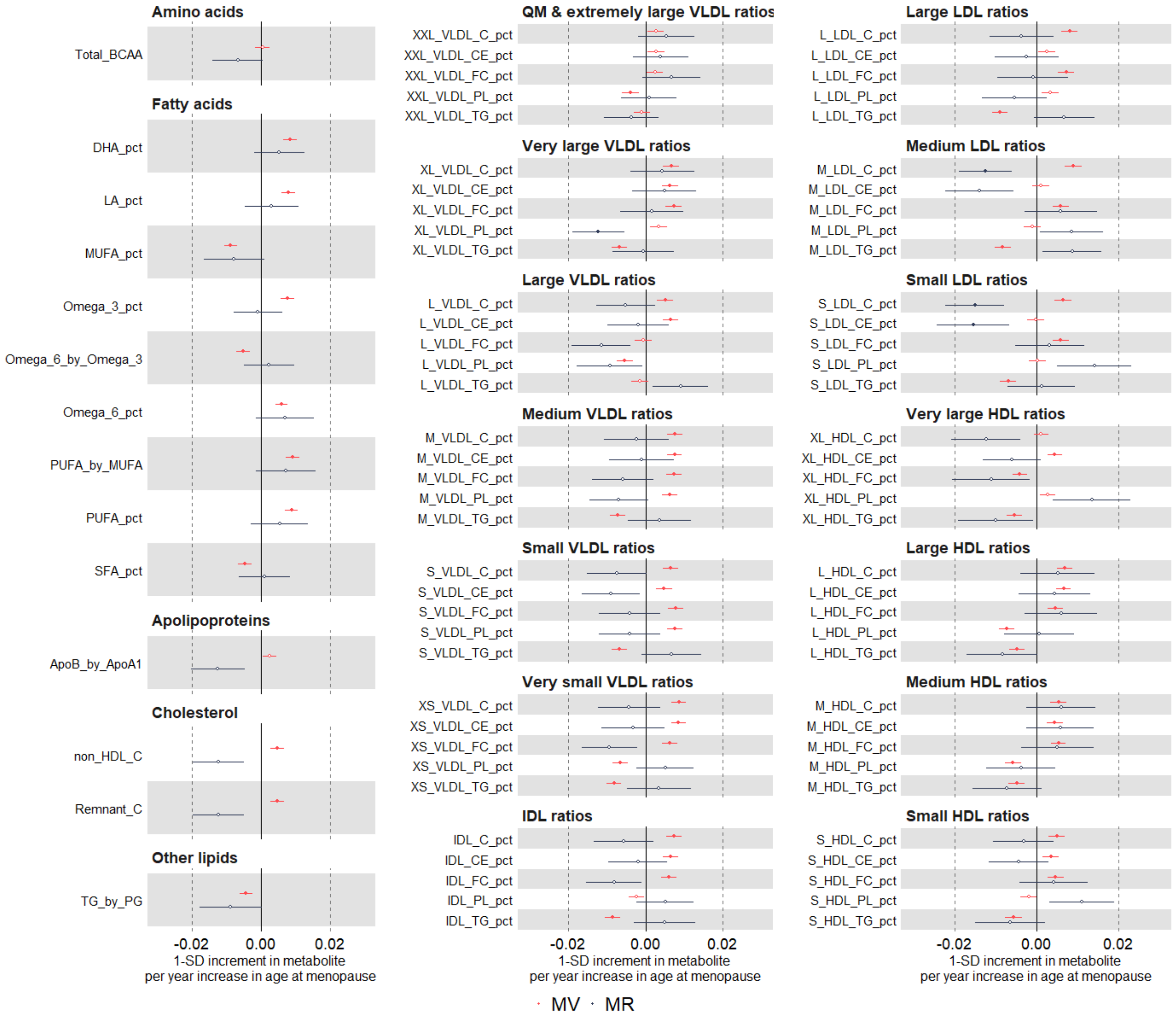

Suppl Fig 12. Multivariable regression estimates for the relation between older age at natural menopause and metabolic measures among females (comparing different model adjustments). Footnote: model 1 (unadjusted) (red); model 2 (main model, age at baseline, education, and body composition at age 10) (green); model 3 (additionally adjusted for BMI, smoking ad alcohol status at baseline) (black).

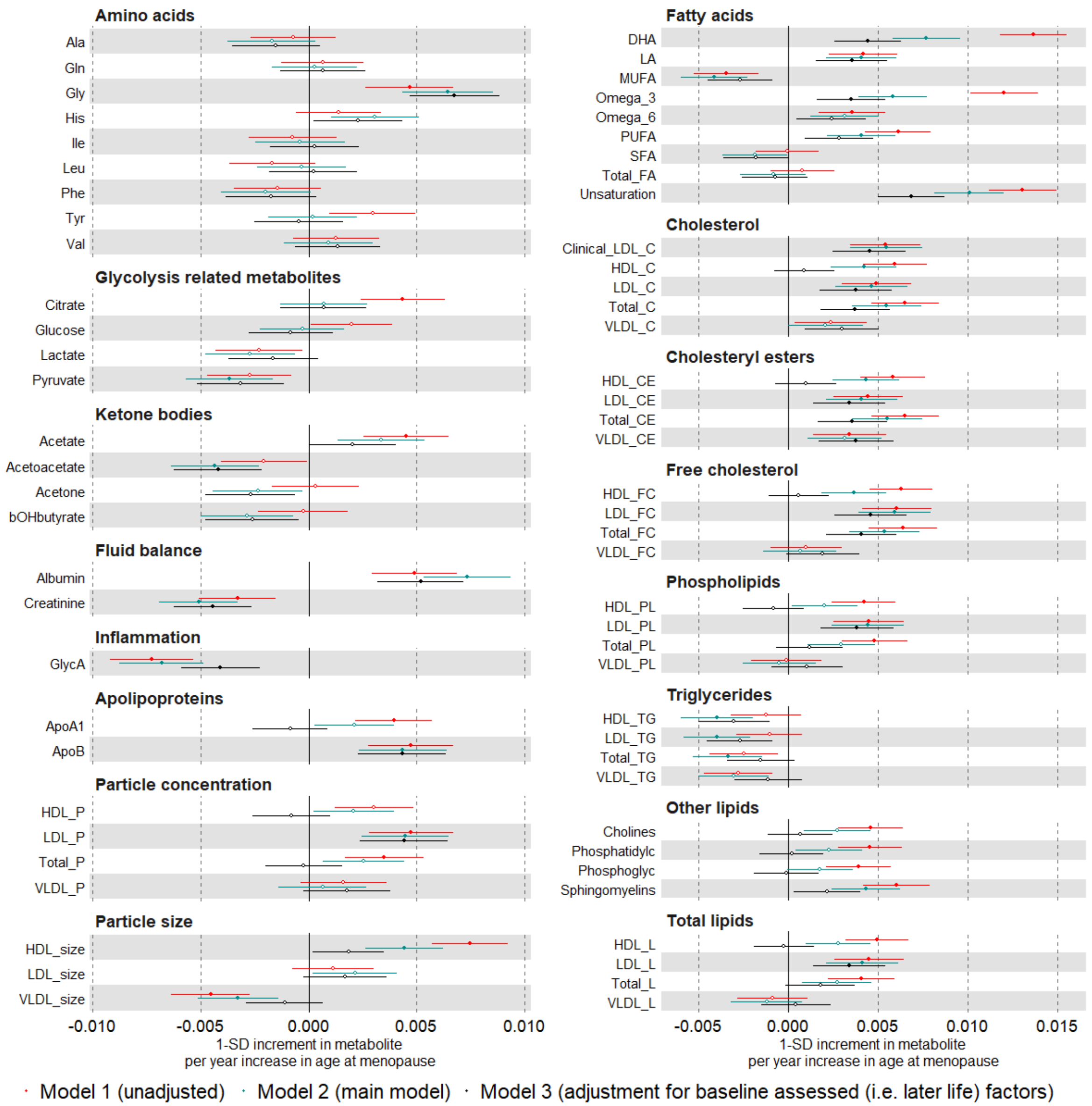

Suppl Fig 12. Continued

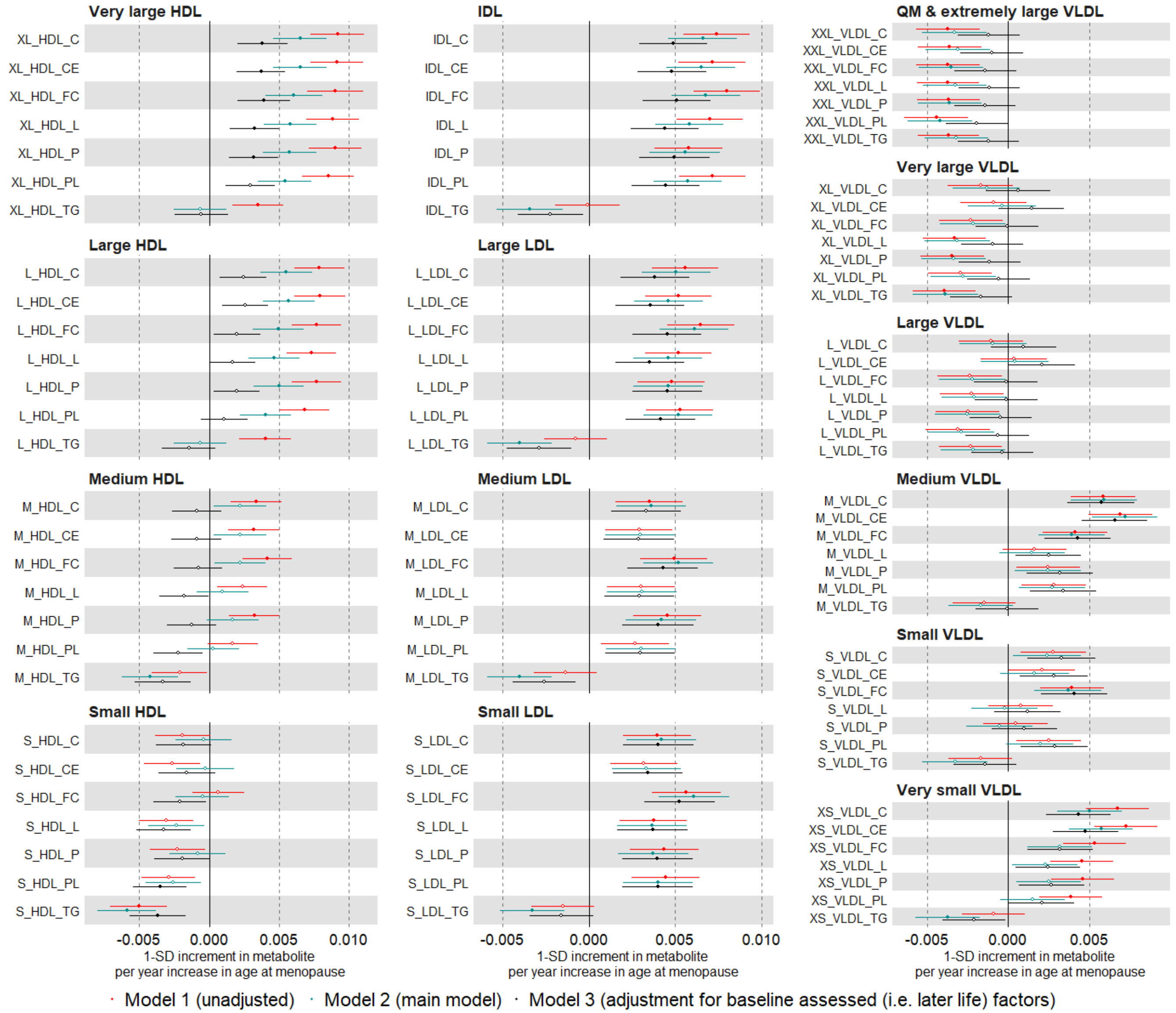

Suppl Fig 12. continued

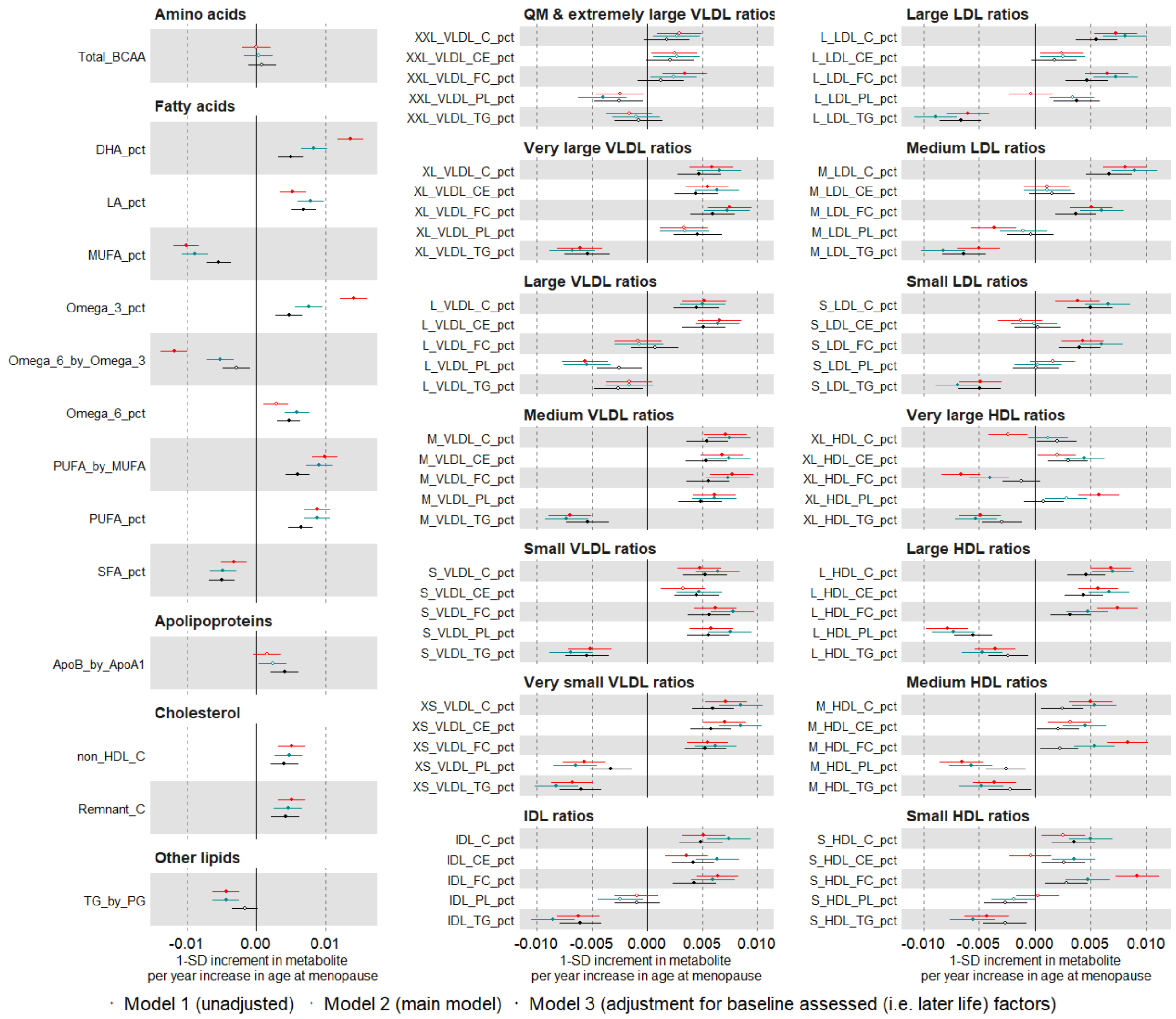

Suppl Fig 13. Multivariable regression estimates for the relation between age at natural menopause (categorised: <49 (reference), 49-50, 51-53, >53 years) and metabolic measures among females. Model adjusted for age at baseline, education, and body composition at age 10.

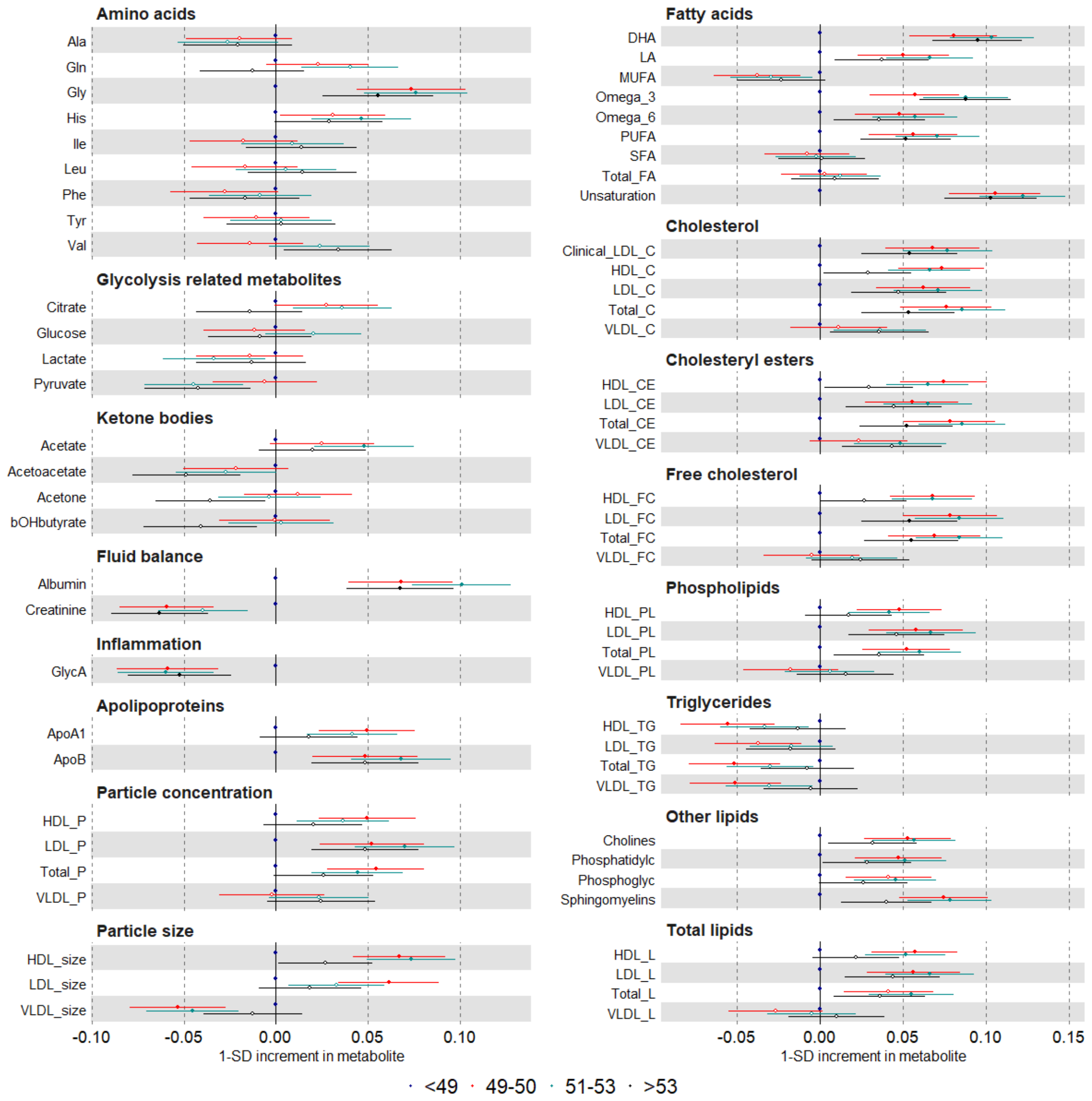

Suppl Fig 13. Continued

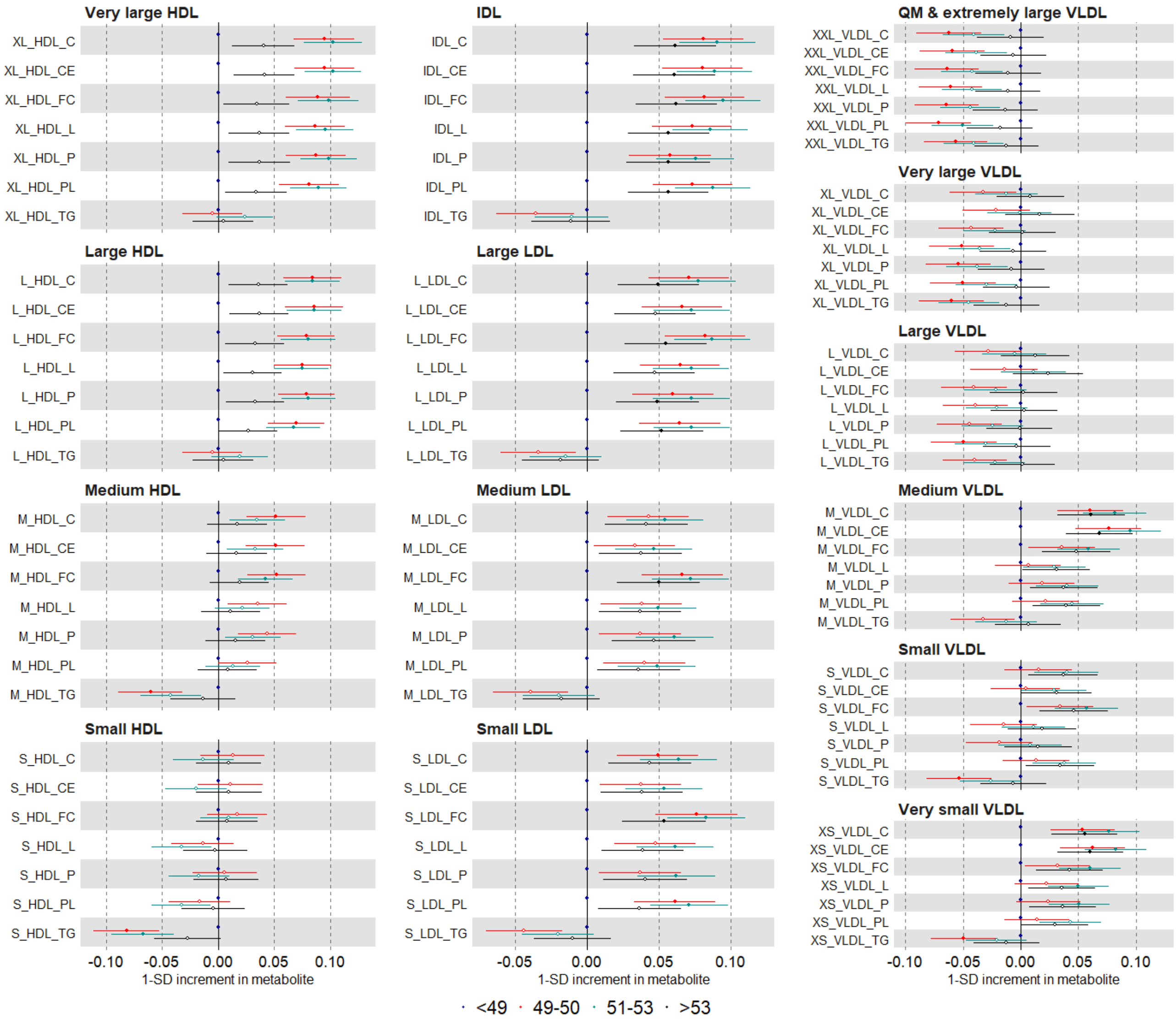

Suppl Fig 13. Continued

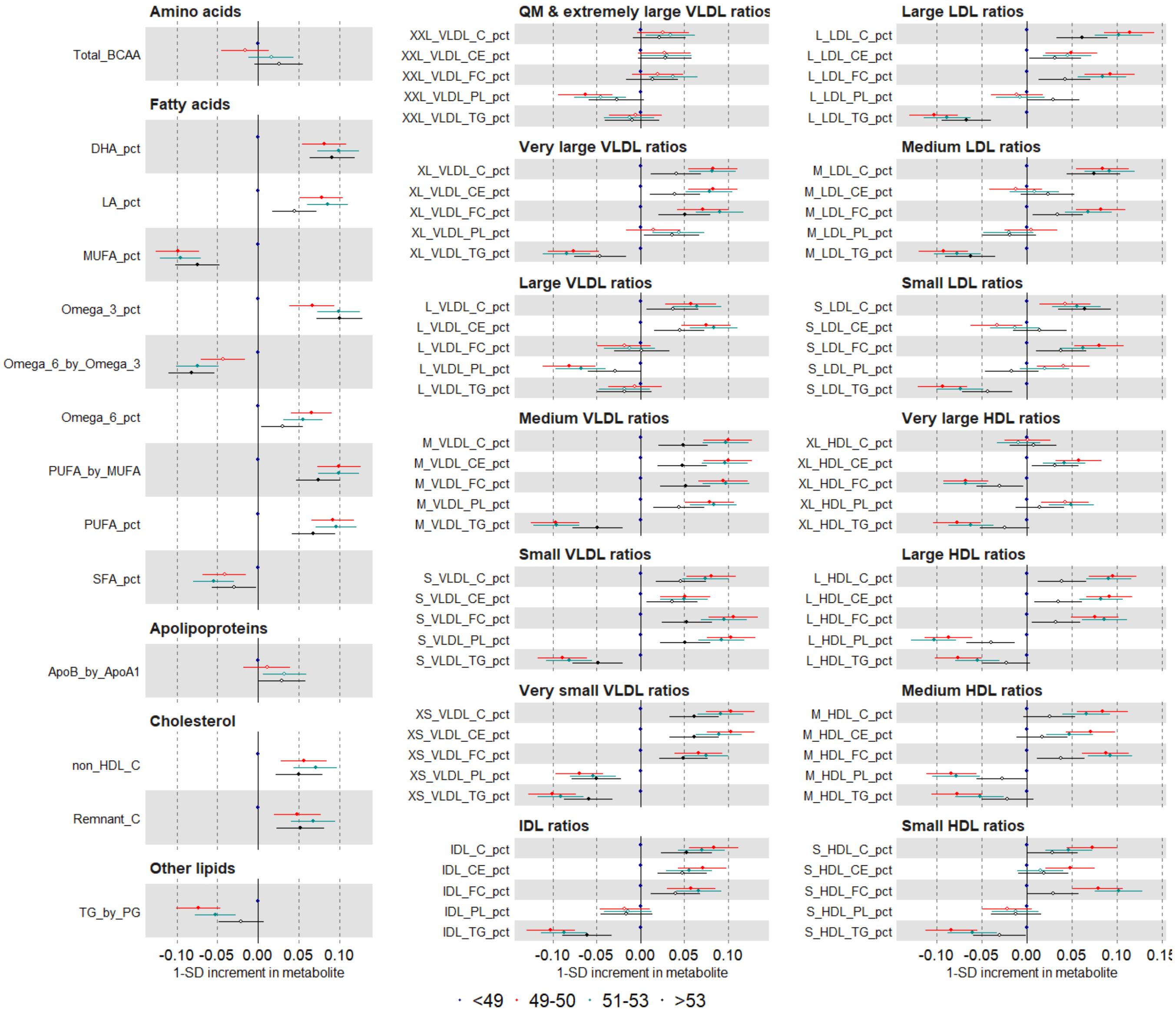

Suppl Fig 14. Multivariable regression estimates for the relation between age at menopause (restricted cubic splines with knots placed at ages 40, 49, 52, and 56) and metabolic measures among females (Mean predicted outcome levels at different menopause ages for a women who is 60 years old, had an average body size at age 10 and is educated to college or university level)

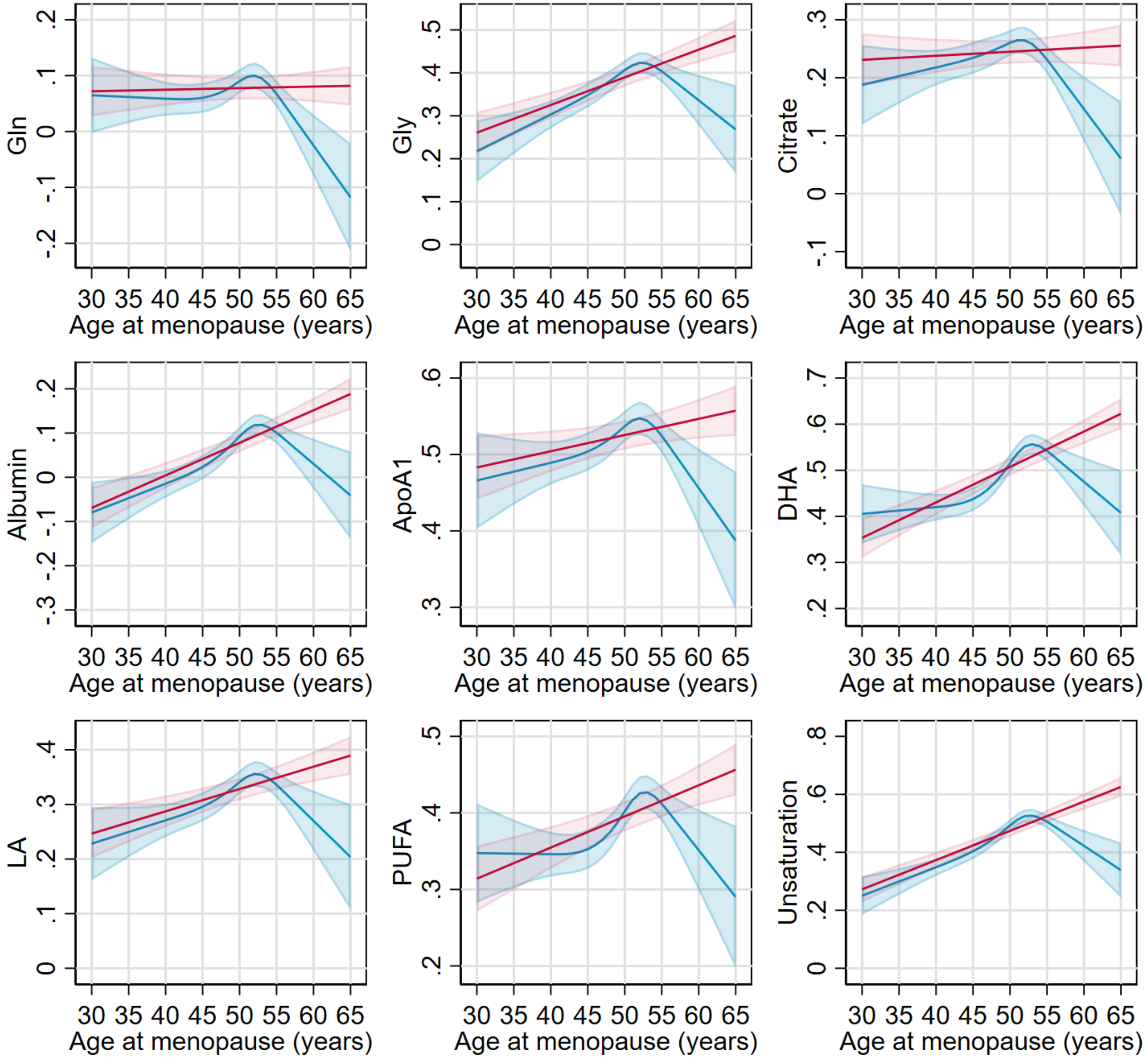

Suppl Fig 14 .continued

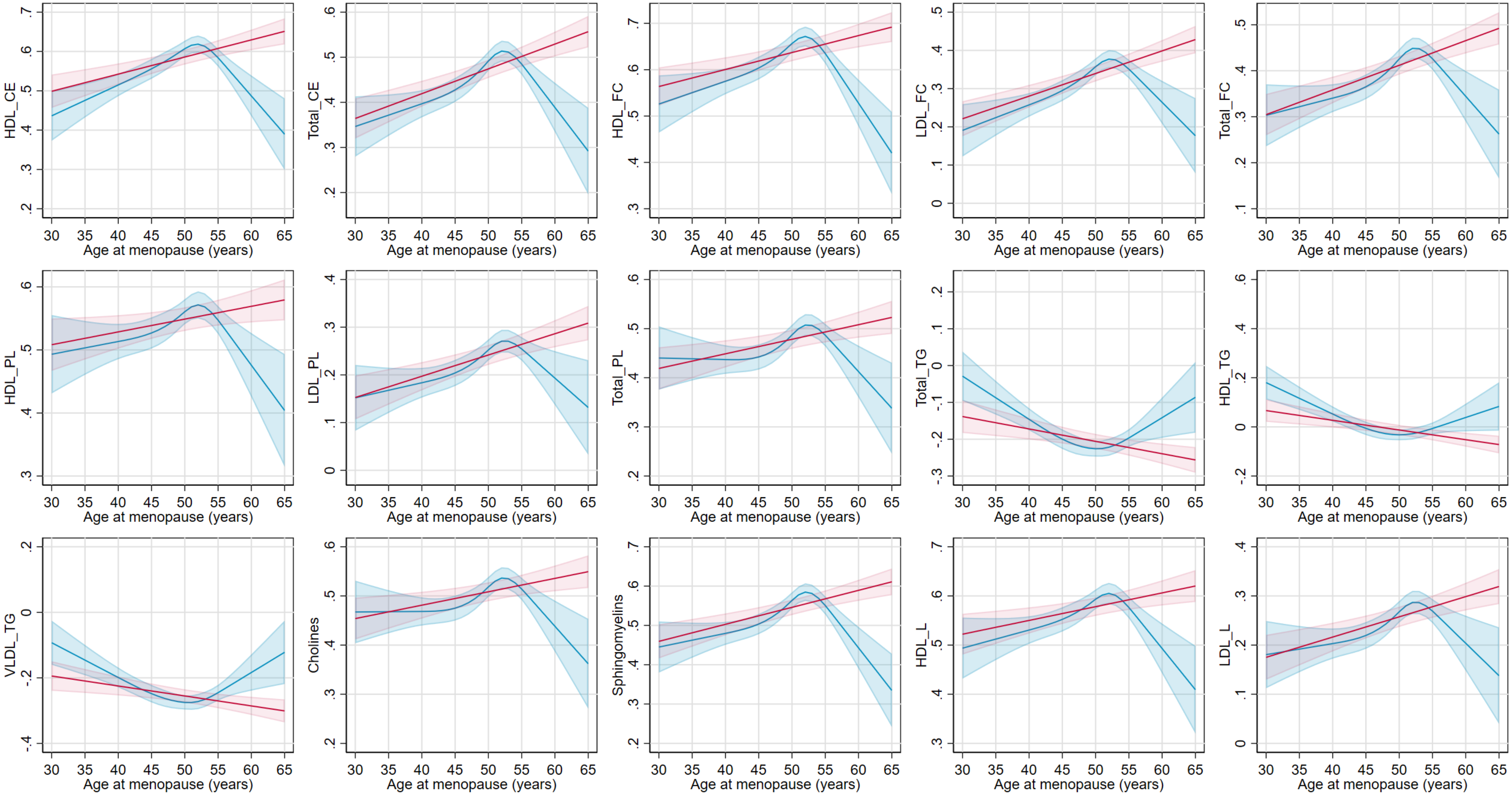

Suppl Fig 15. Age-combined and age-stratified estimates for the association between age at natural menarche and clinical chemistry biomarkers using multivariable regression (red) and one-sample Mendelian randomization (blue) restricted to women with data on age at menopause or two-sample Mendelian randomization (black) using data from all women.

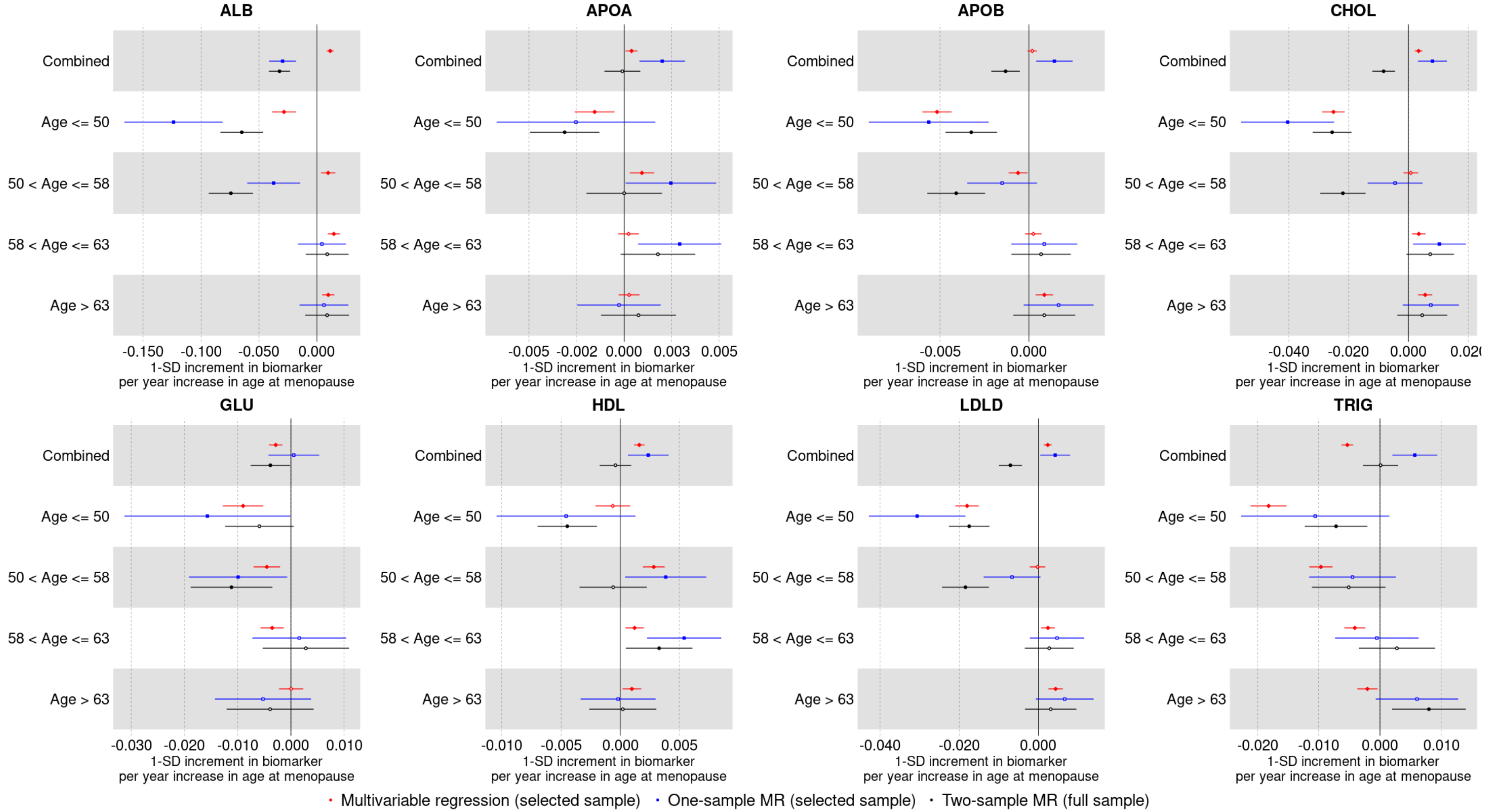


Multivariable regression was performed for women with non-missing data on age at natural menopause using ordinary least squares (OLS) regression. One-sample Mendelian randomization was performed for women with non-missing data on age at natural menopause using two-stage least square (2SLS) regression. Two-sample Mendelian randomization was performed for all women regardless of missing data on age at natural menopause using the inverse variance weighted (IVW) method.

Suppl Fig 16A. Age-combined and age-stratified estimates for the association between age at natural menarche and clinical chemistry biomarkers excluding users of statins at baseline estimated using multivariable regression (red) and Mendelian randomization (blue) restricted to women with data on age at menopause or Mendelian randomization (black) using data from all women.

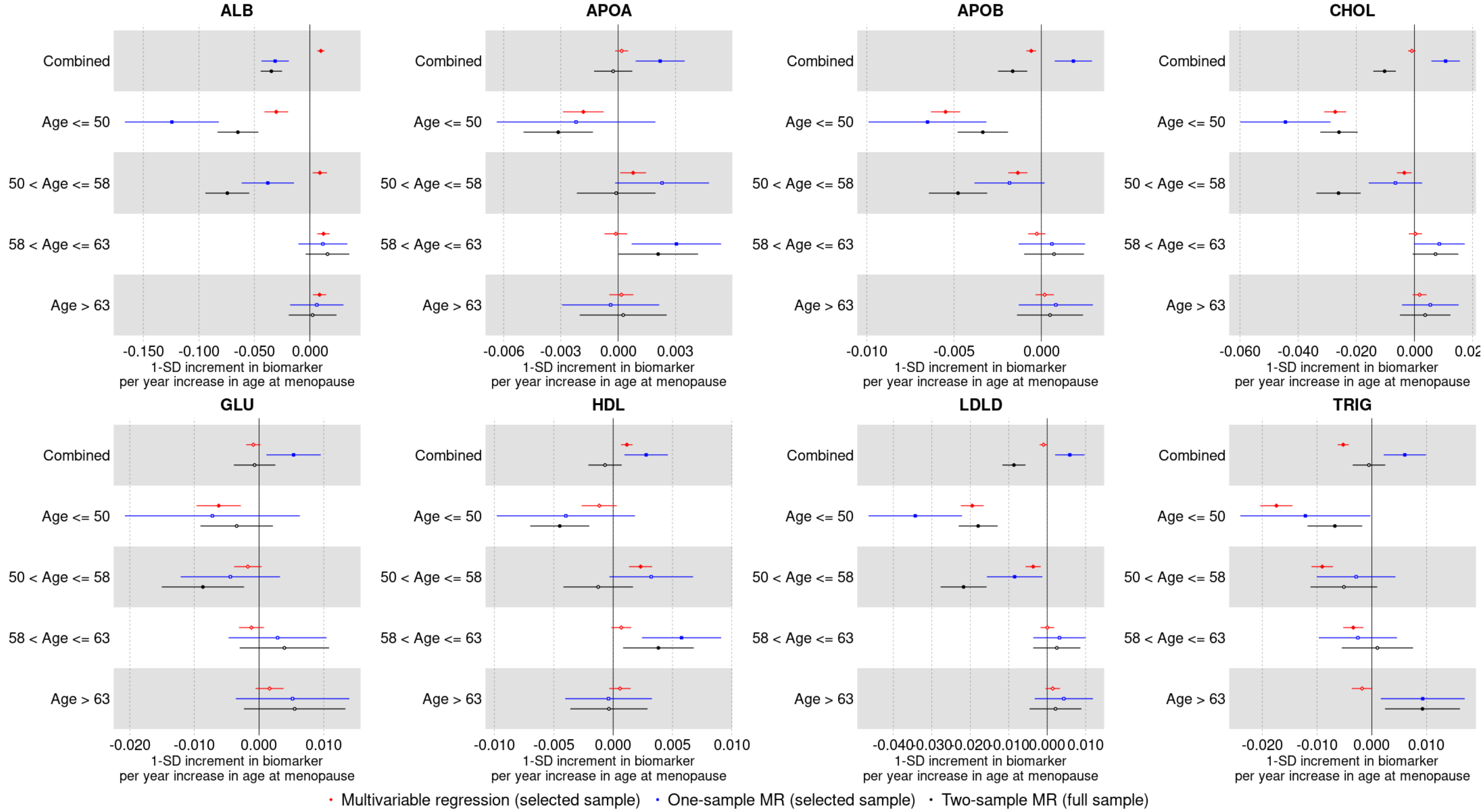


Multivariable regression was performed for women with non-missing data on age at natural menopause using ordinary least squares (OLS) regression. One-sample Mendelian randomization was performed for women with non-missing data on age at natural menopause using two-stage least square (2SLS) regression. Two-sample Mendelian randomization was performed for all women regardless of missing data on age at natural menopause using the inverse variance weighted (IVW) method.

Suppl Fig 16B. Age-combined and age-stratified estimates for the association between age at natural menarche and clinical chemistry biomarkers excluding users of hormone replacement therapy (HRT) at baseline estimated using multivariable regression (red) and Mendelian randomization (blue) restricted to women with data on age at menopause or Mendelian randomization (black) using data from all women.

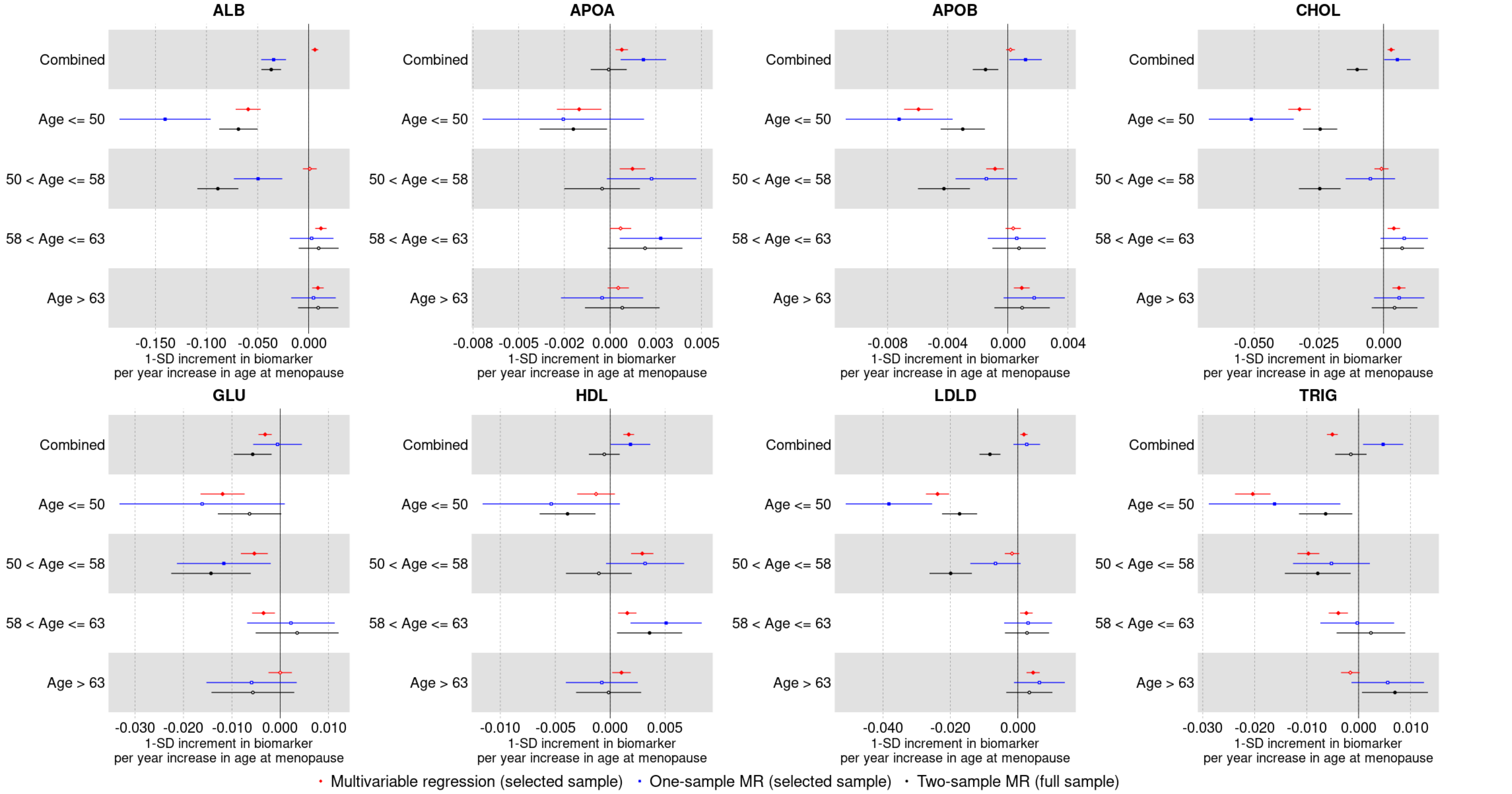


Multivariable regression was performed for women with non-missing data on age at natural menopause using ordinary least squares (OLS) regression. One-sample Mendelian randomization was performed for women with non-missing data on age at natural menopause using two-stage least square (2SLS) regression. Two-sample Mendelian randomization was performed for all women regardless of missing data on age at natural menopause using the inverse variance weighted (IVW) method.

Suppl Fig 17. Mendelian randomization estimates for the relation between reproductive markers and conventional biomarkers among unrelated individuals and within siblings.

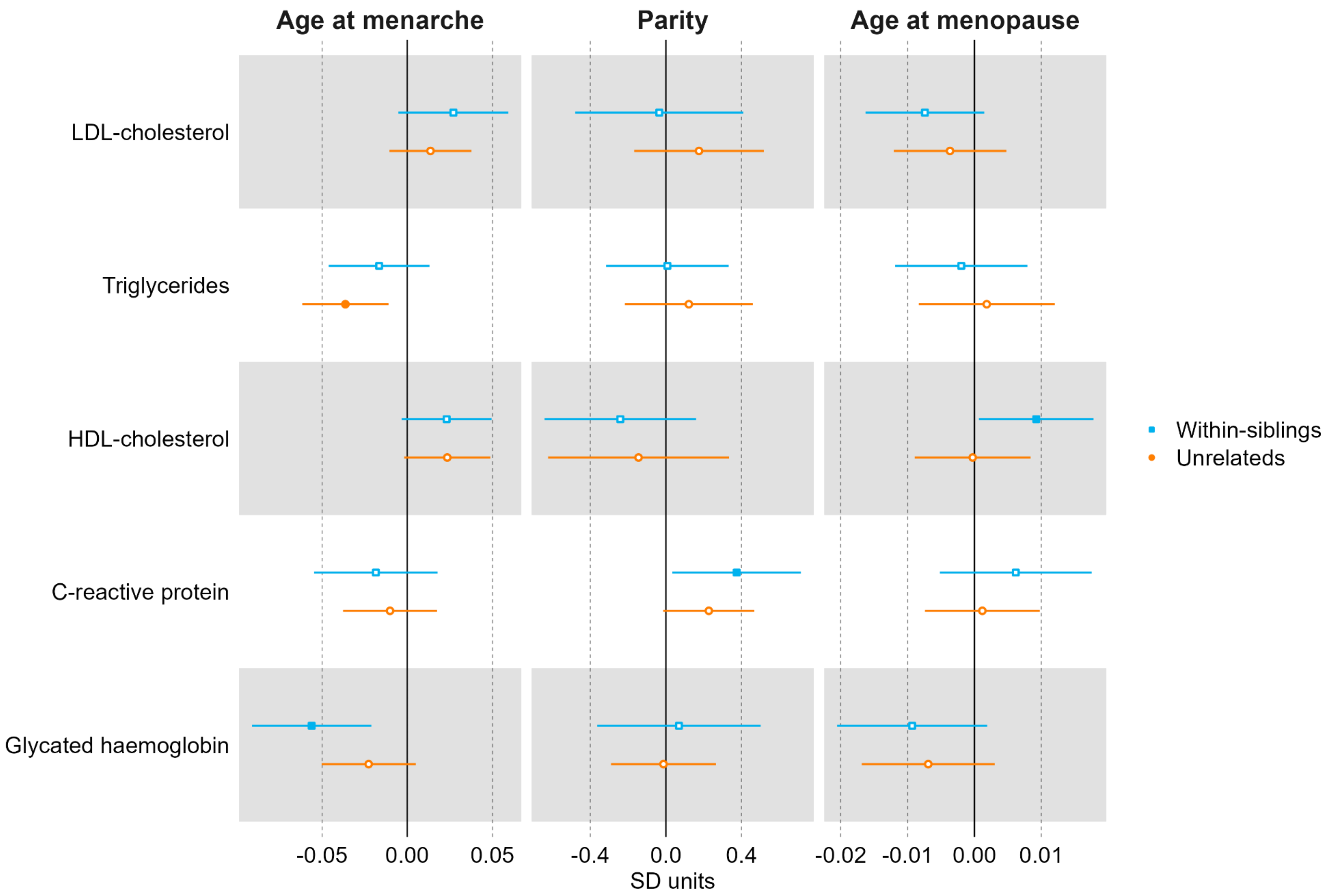

Suppl Fig 18. Mendelian randomization estimates for the relation between older age at menarche and metabolic measures among females (comparing different Mendelian randomization methods)

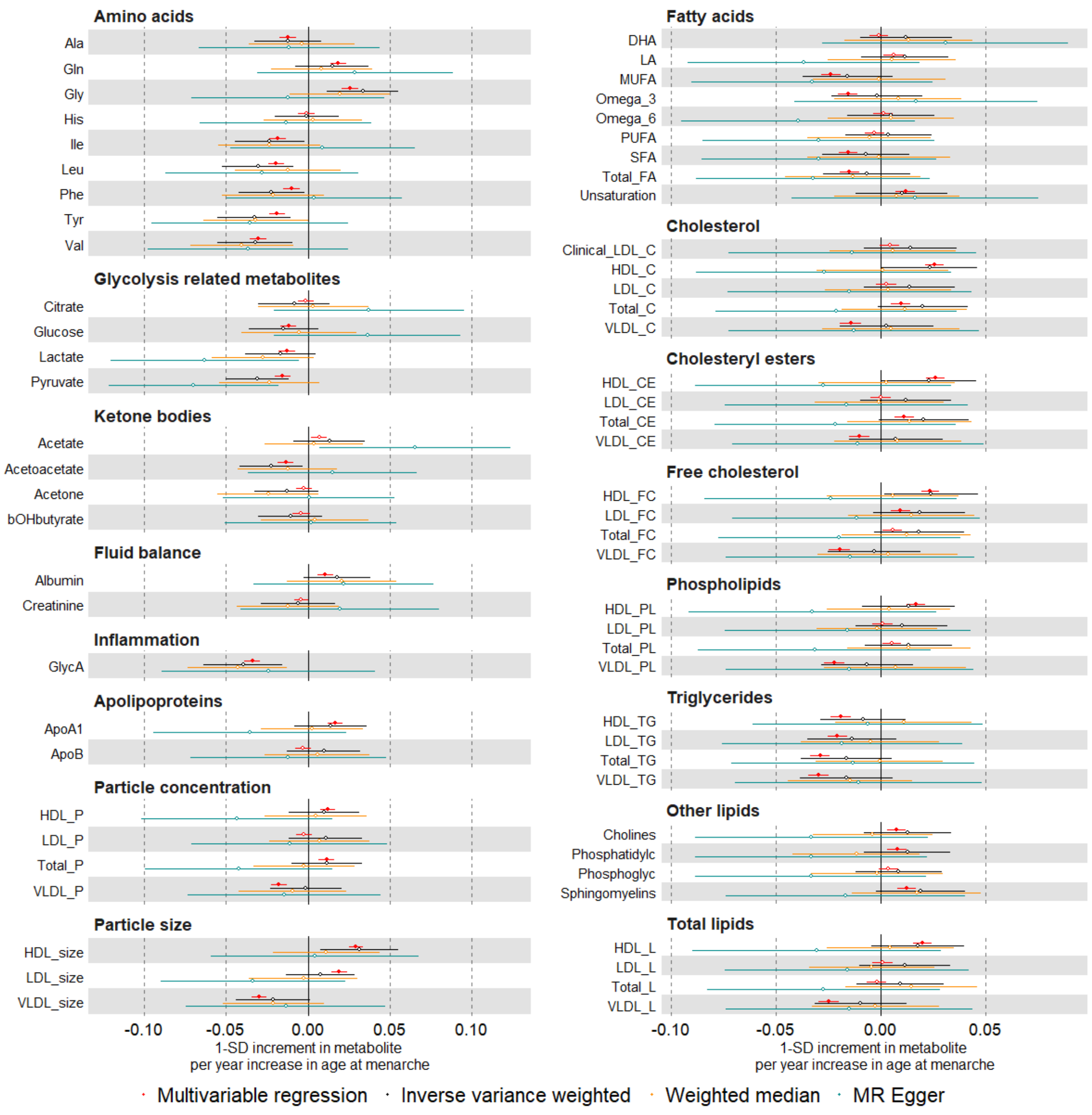

Suppl Fig 18. Continued

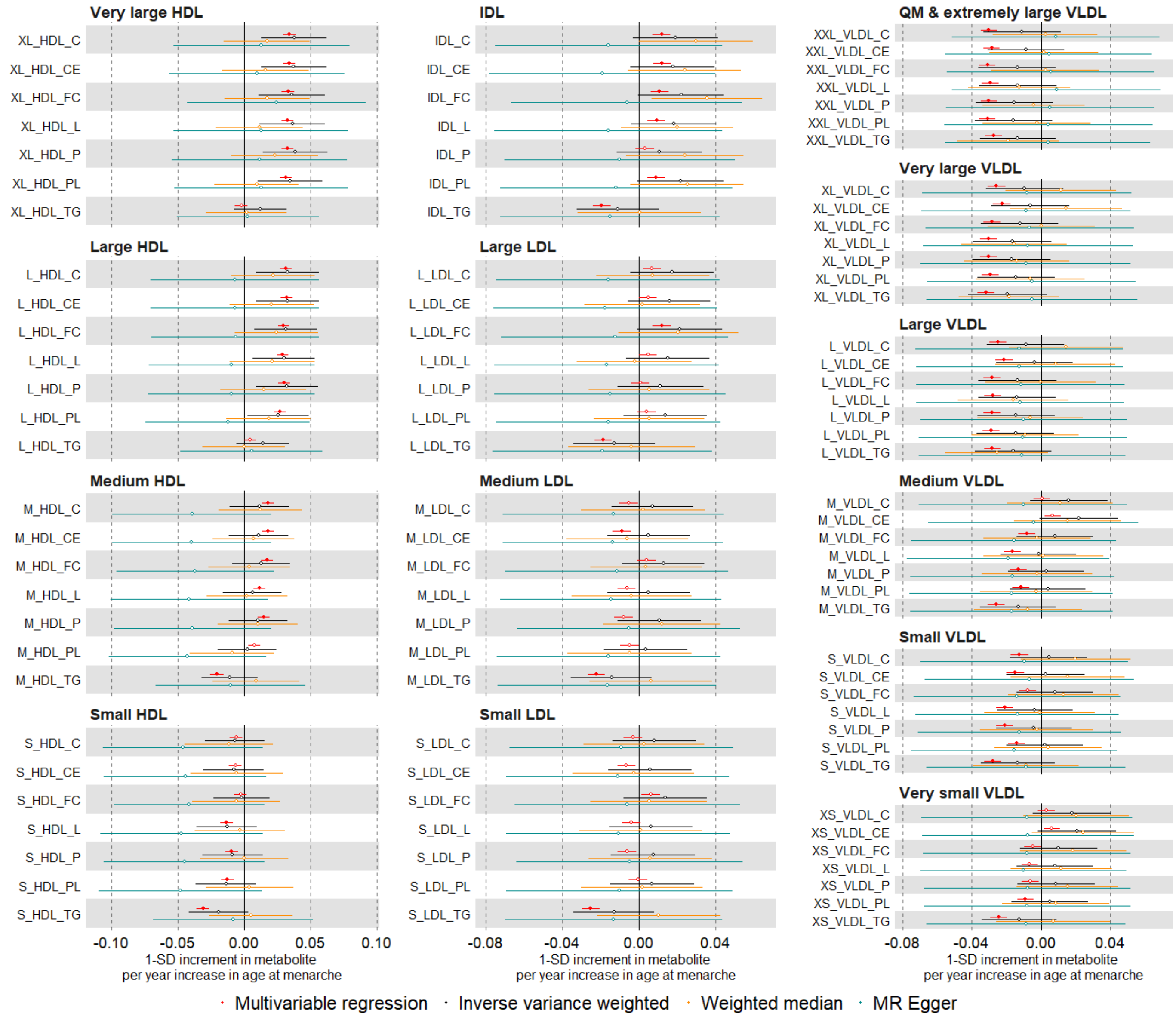

Suppl Fig 18. Continued

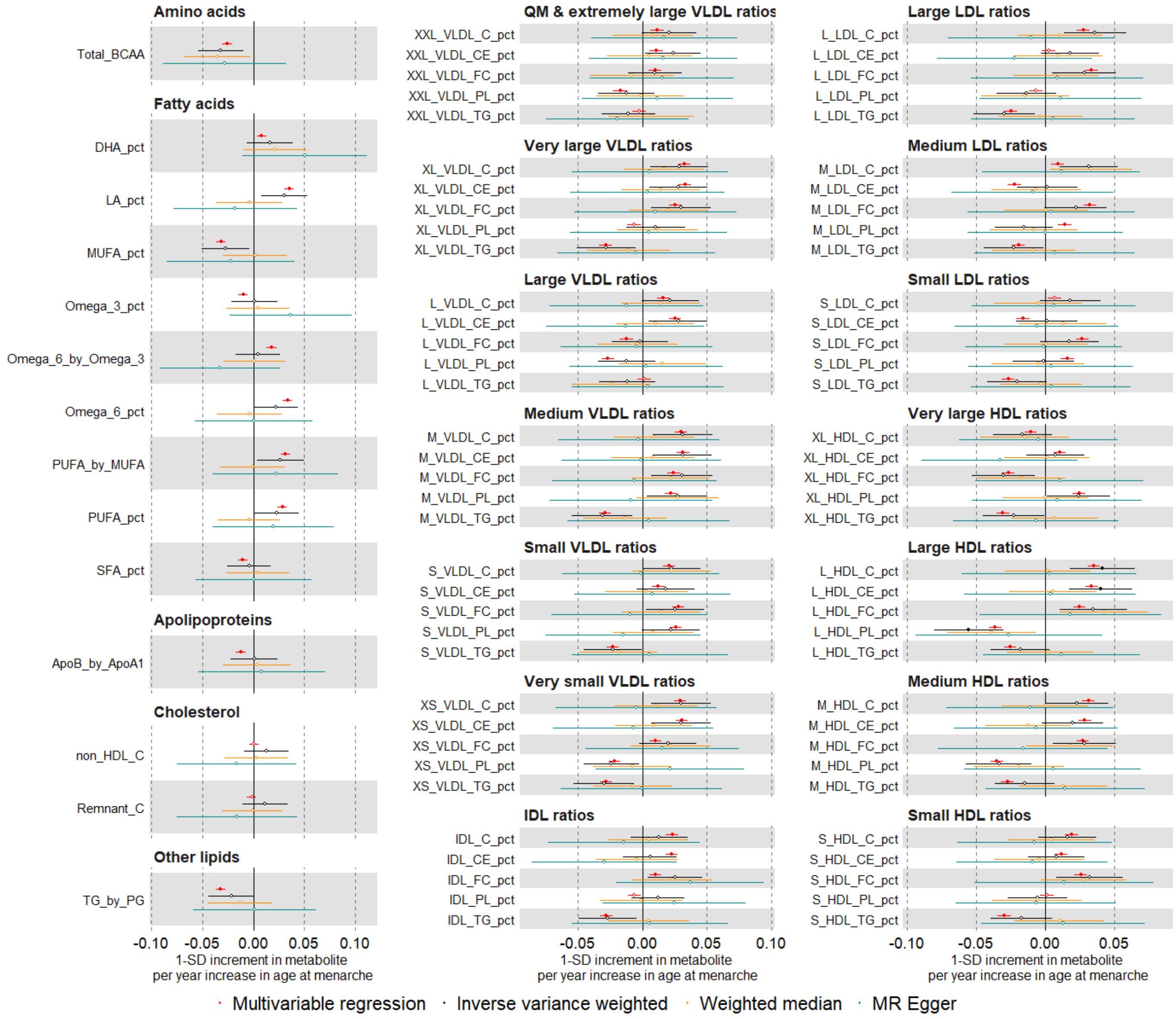

Suppl Fig 19. Mendelian randomization estimates for the relation between higher parity and metabolic measures among females (comparing different Mendelian randomization methods)

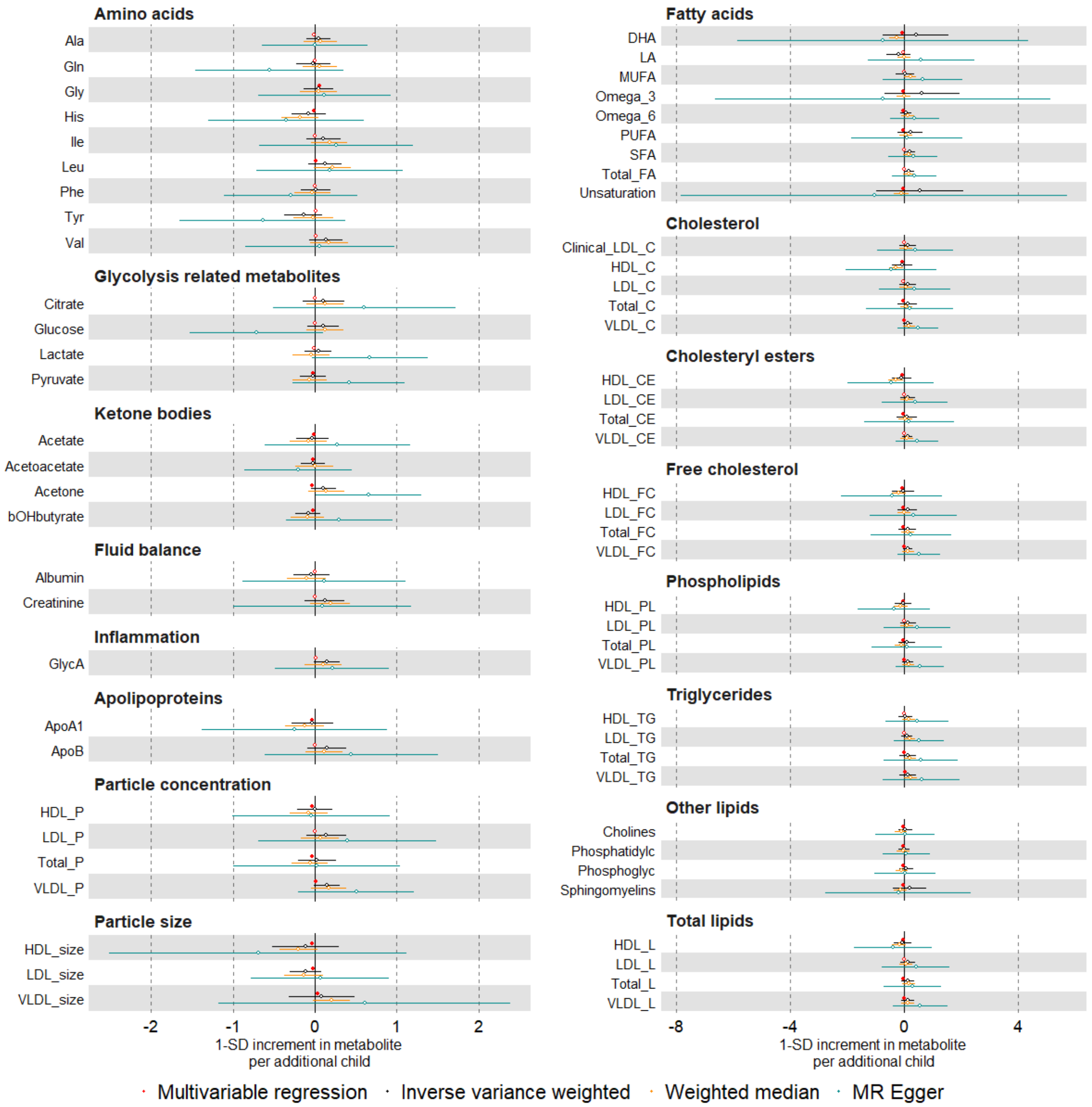

Supp Fig 19 Continued

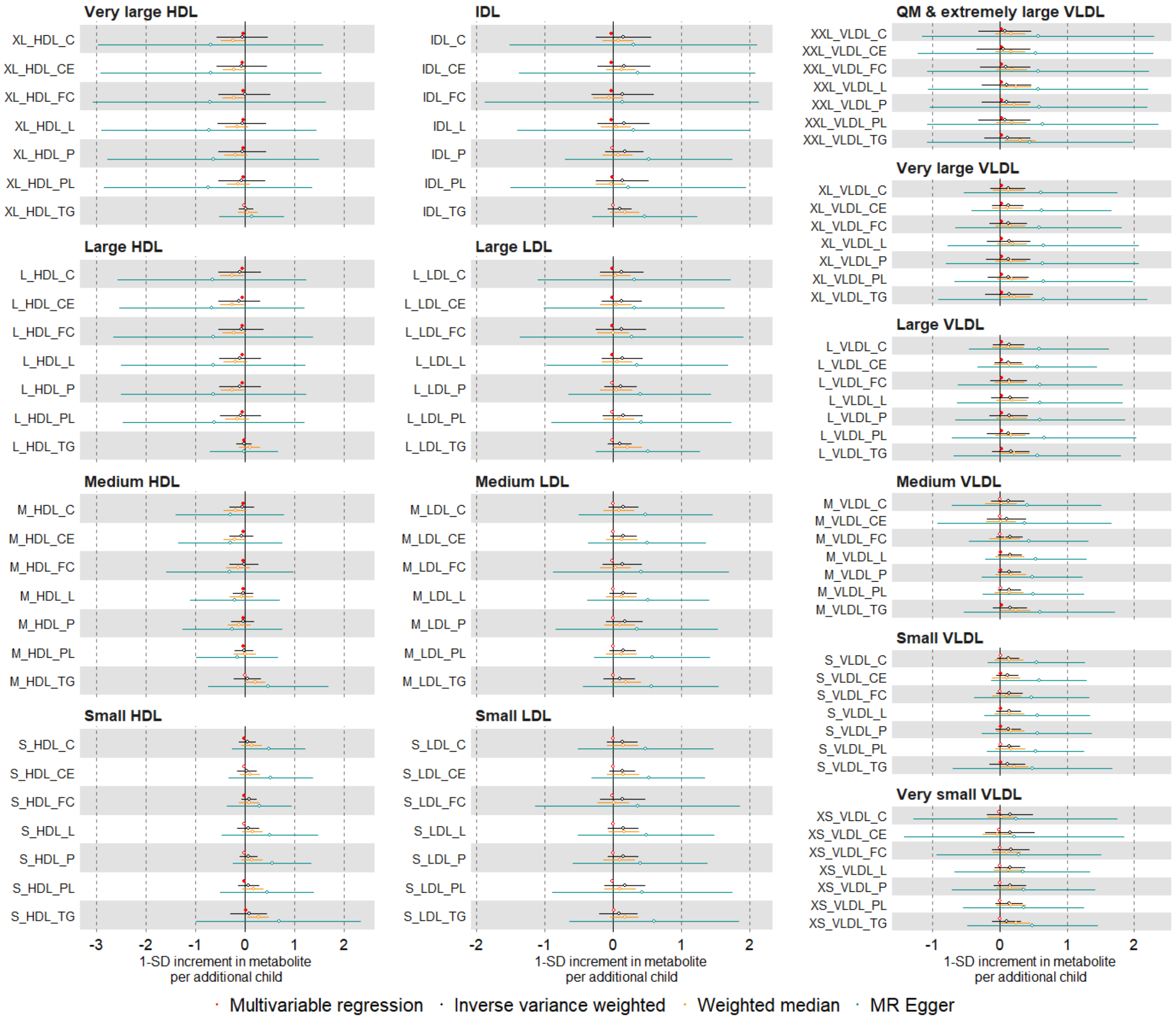

Supp Fig 19 Continued

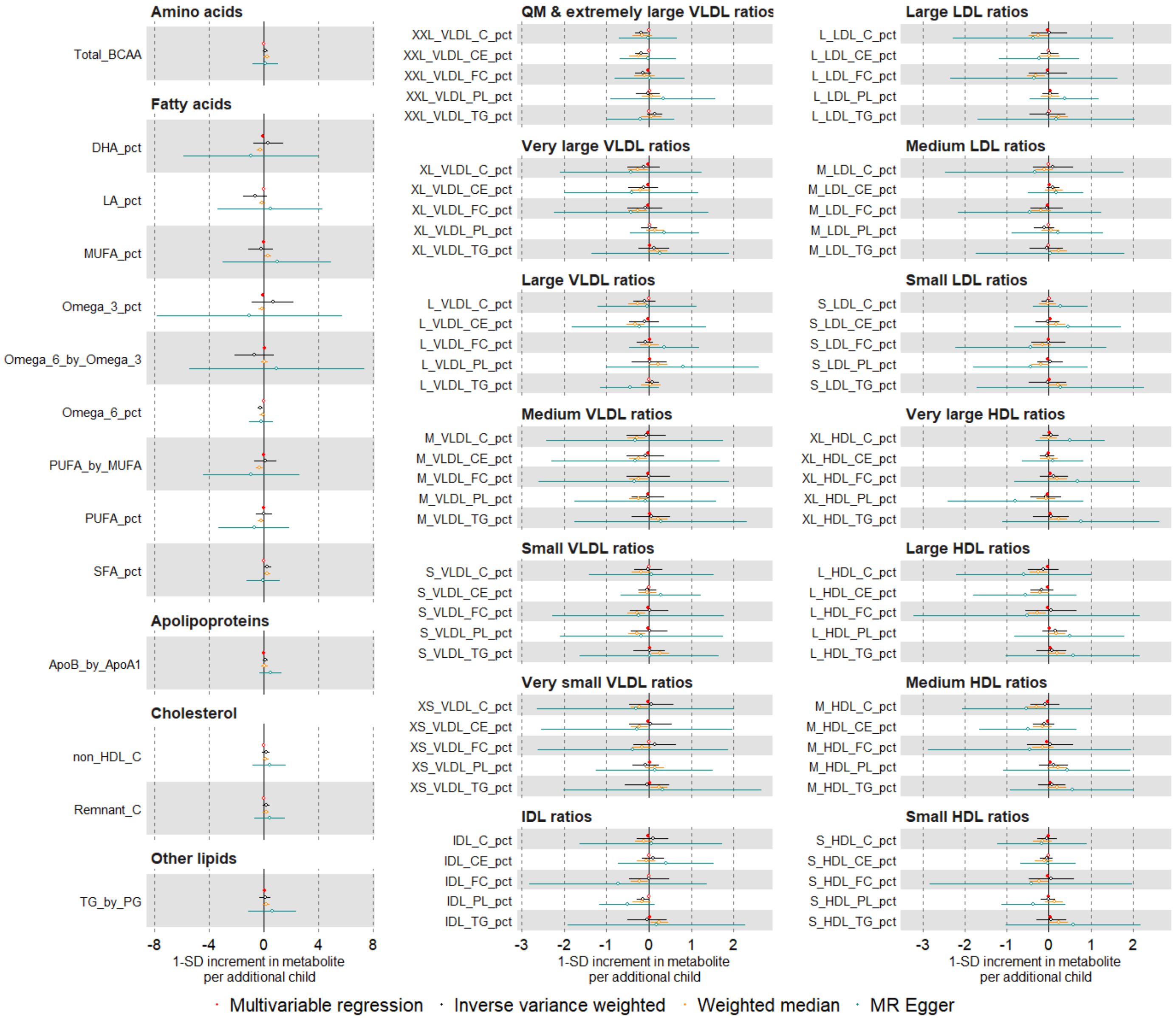

Suppl Fig 20. Mendelian randomization estimates for the relation between older age at natural menopause and metabolic measures among females (compare different Mendelian randomization methods)

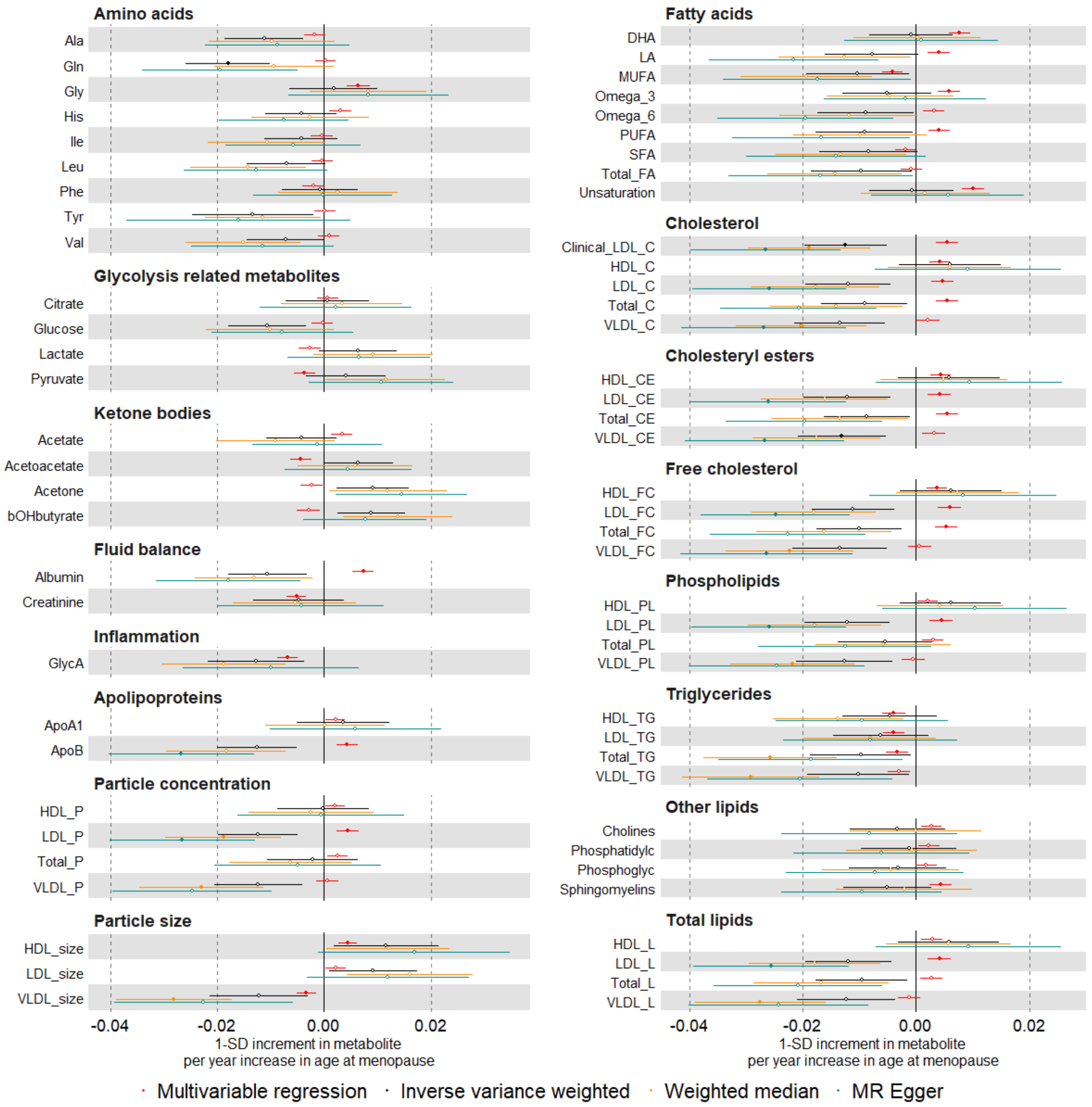

Suppl Fig 20. Continued

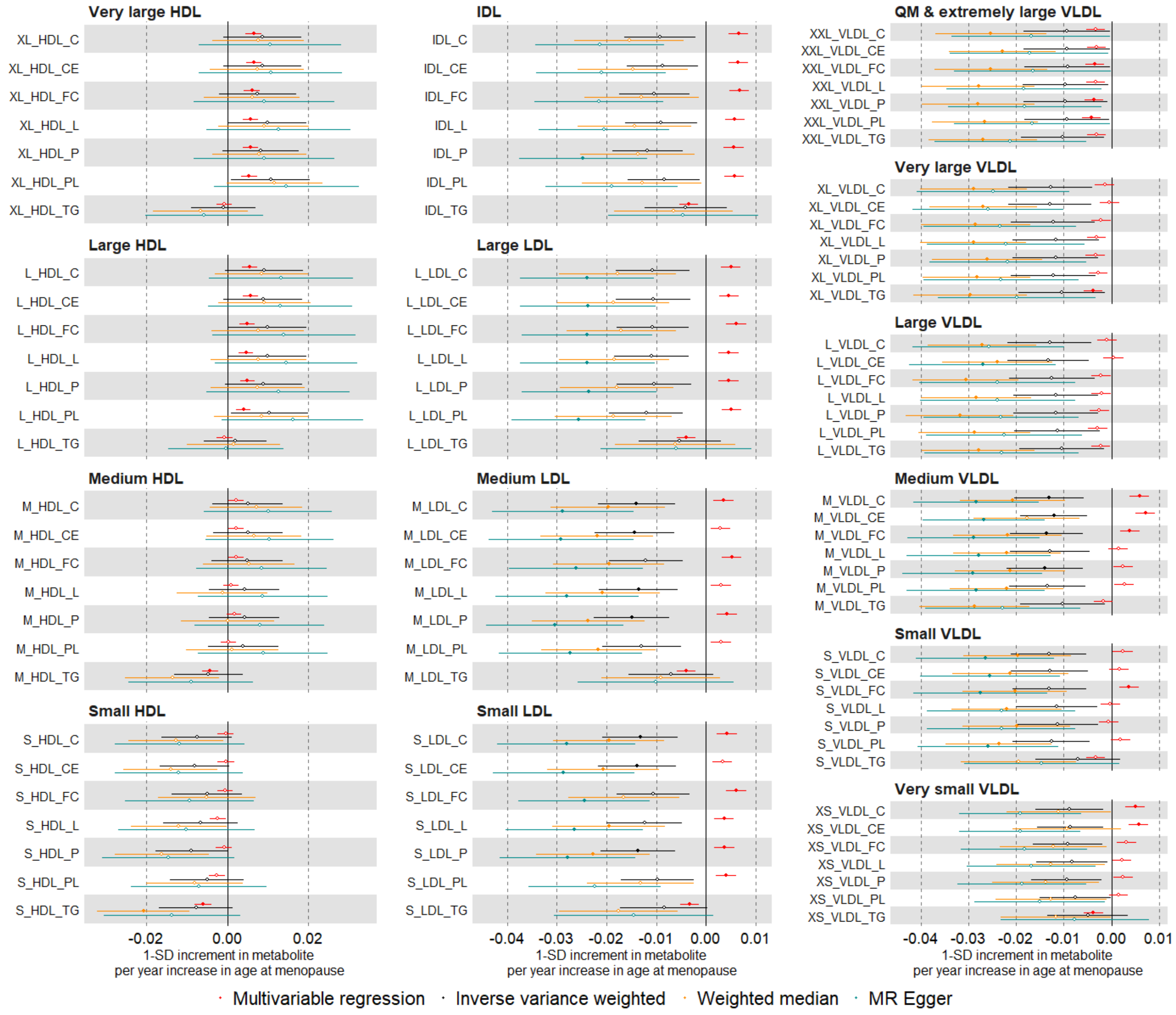

Suppl Fig 20. continued

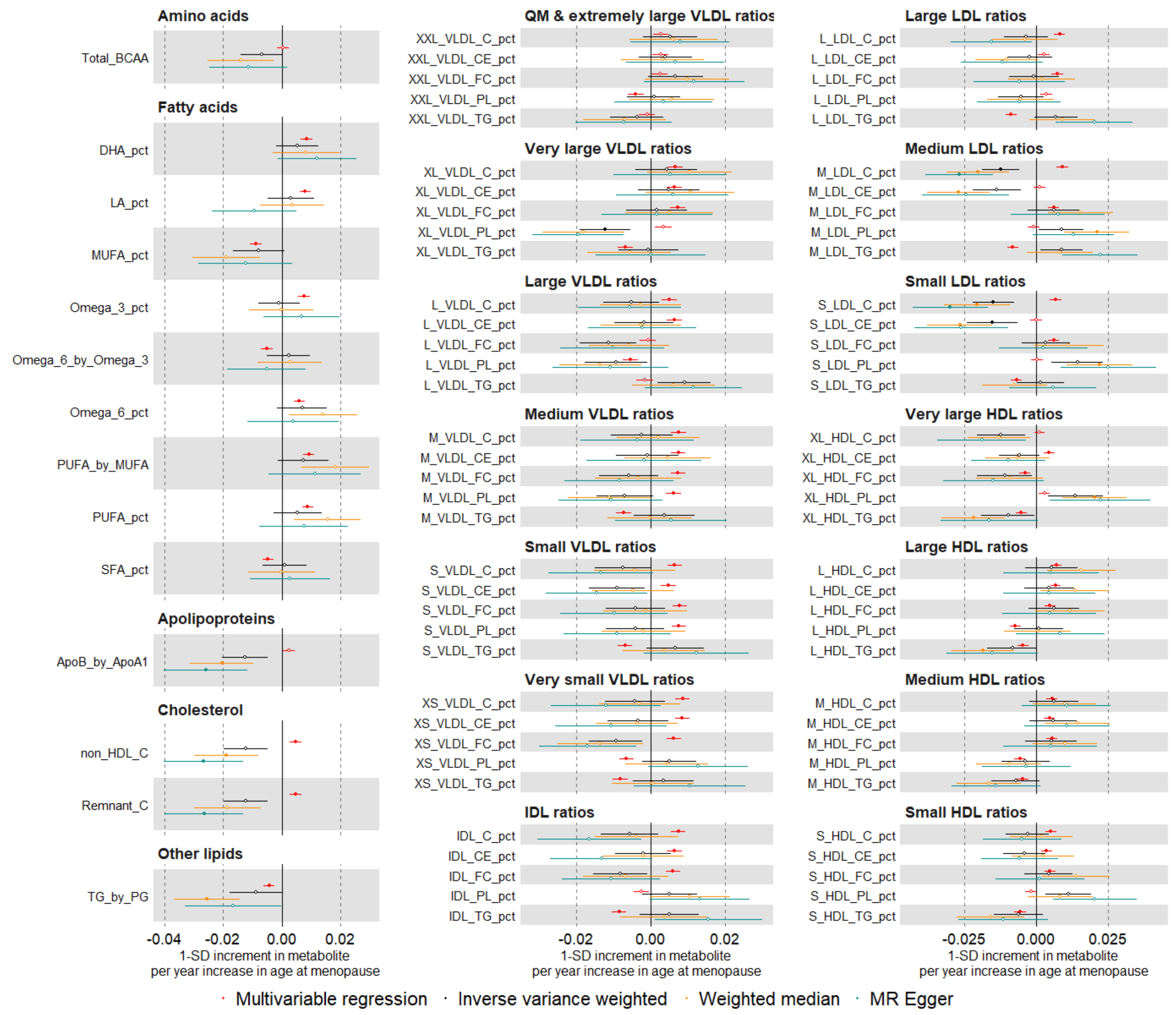

Suppl Fig 21. Mendelian randomization estimates for the relation between older age at menarche and metabolic measures among females (comparing different SNP sets)

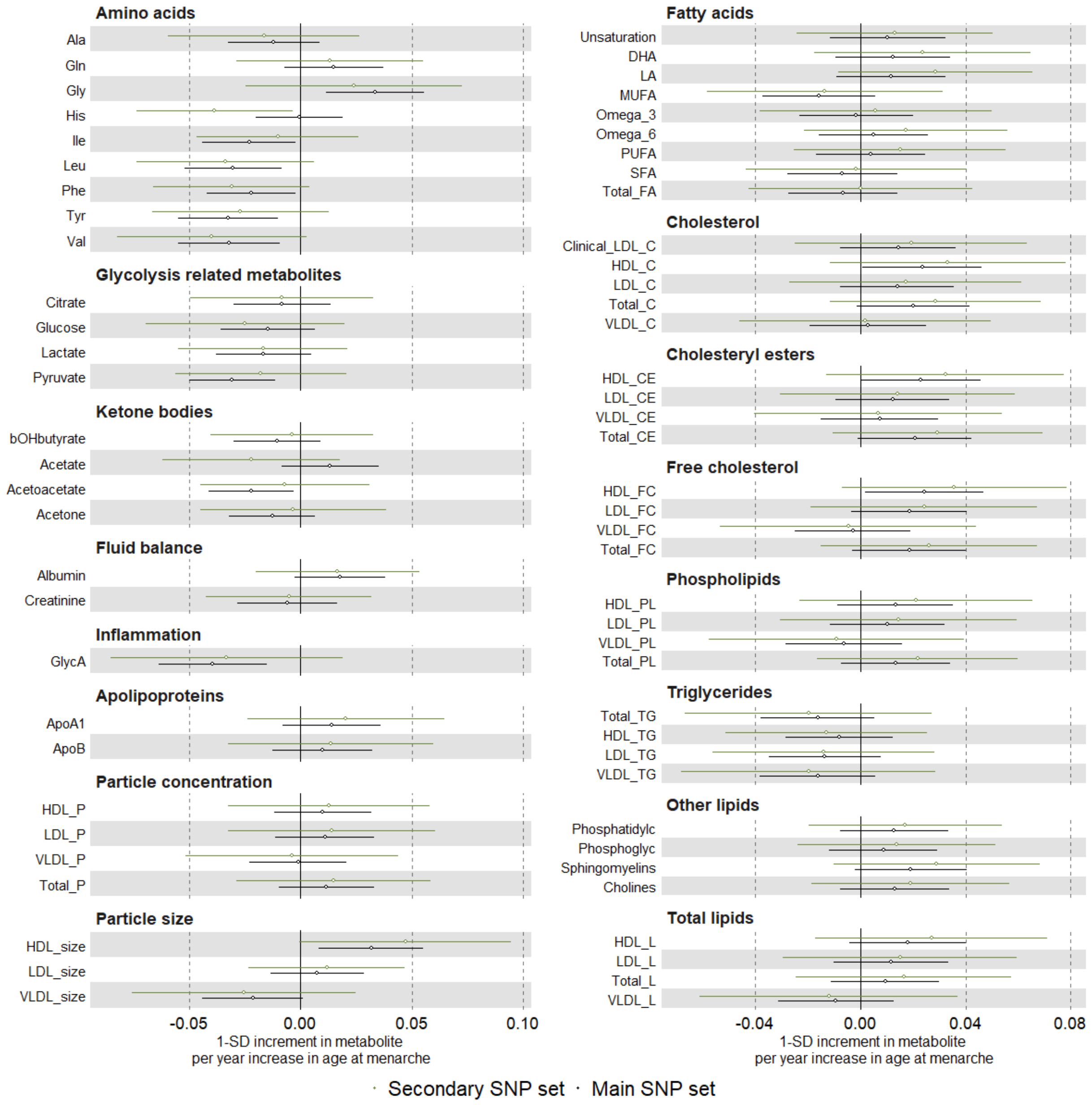

<u>Suppl Fig 21. continued</u>

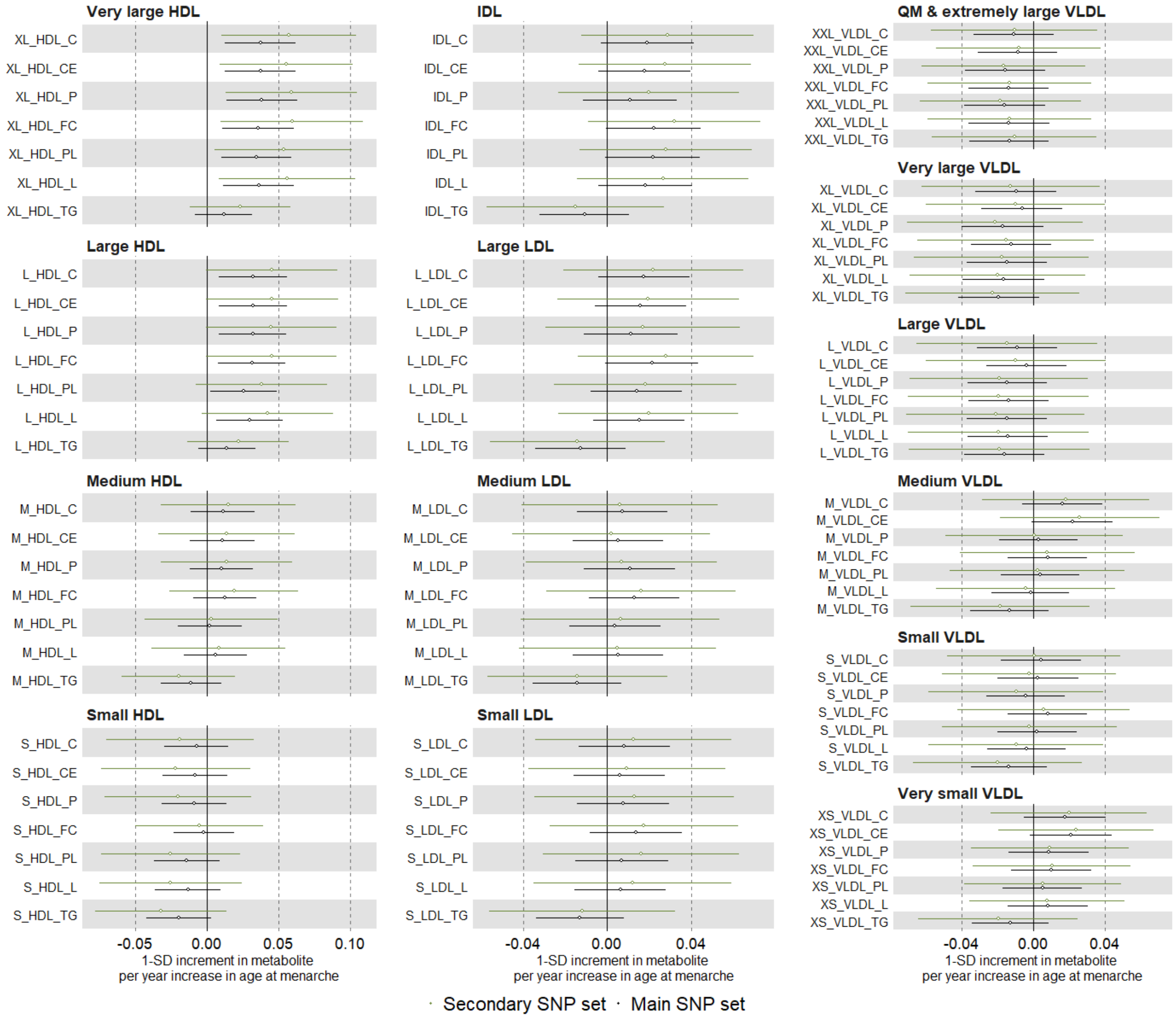

Suppl Fig 21. continued

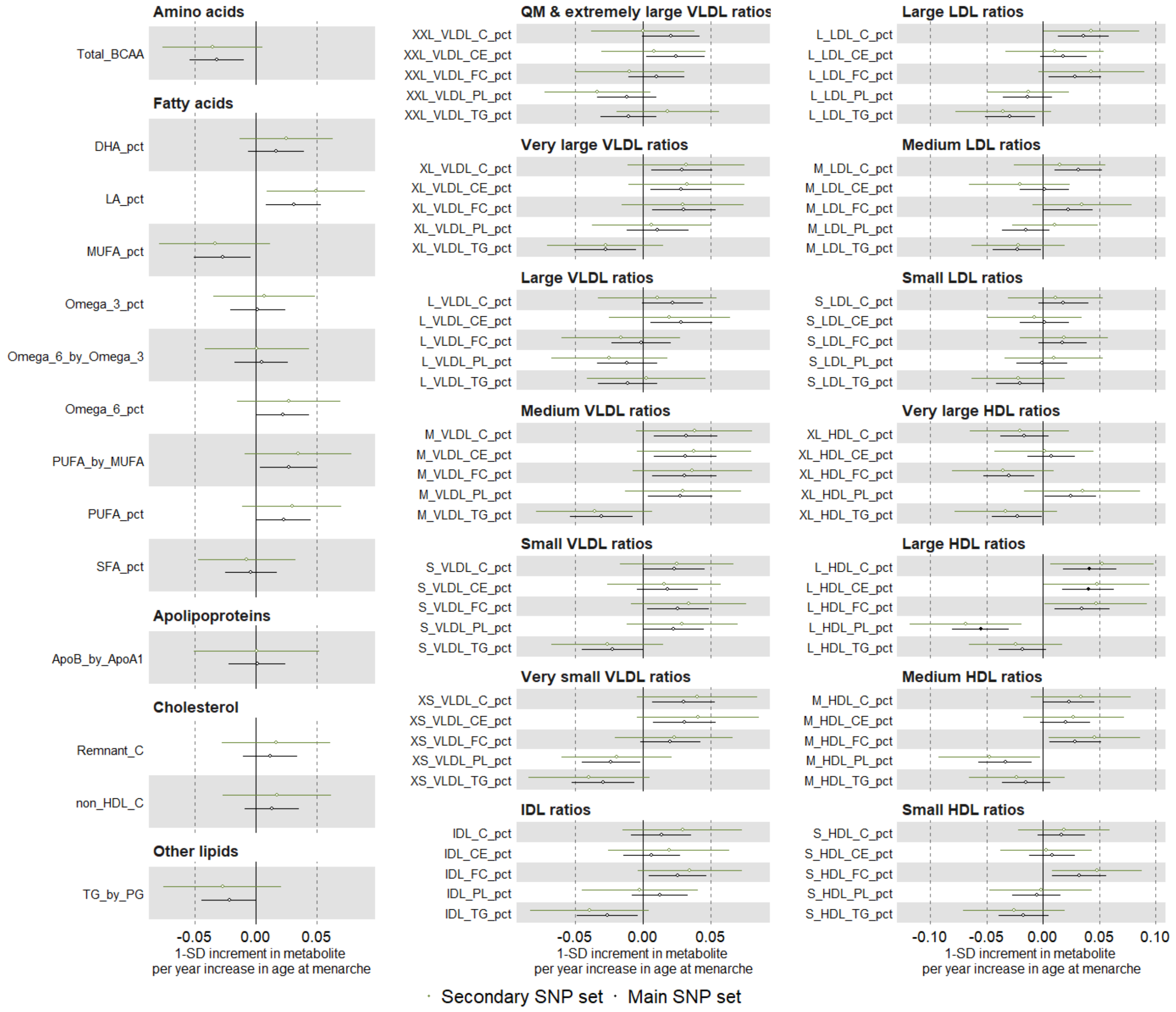

Suppl Fig 22. Mendelian randomization estimates for the relation between older age at natural menopause and metabolic measures among females (comparing different SNP sets)

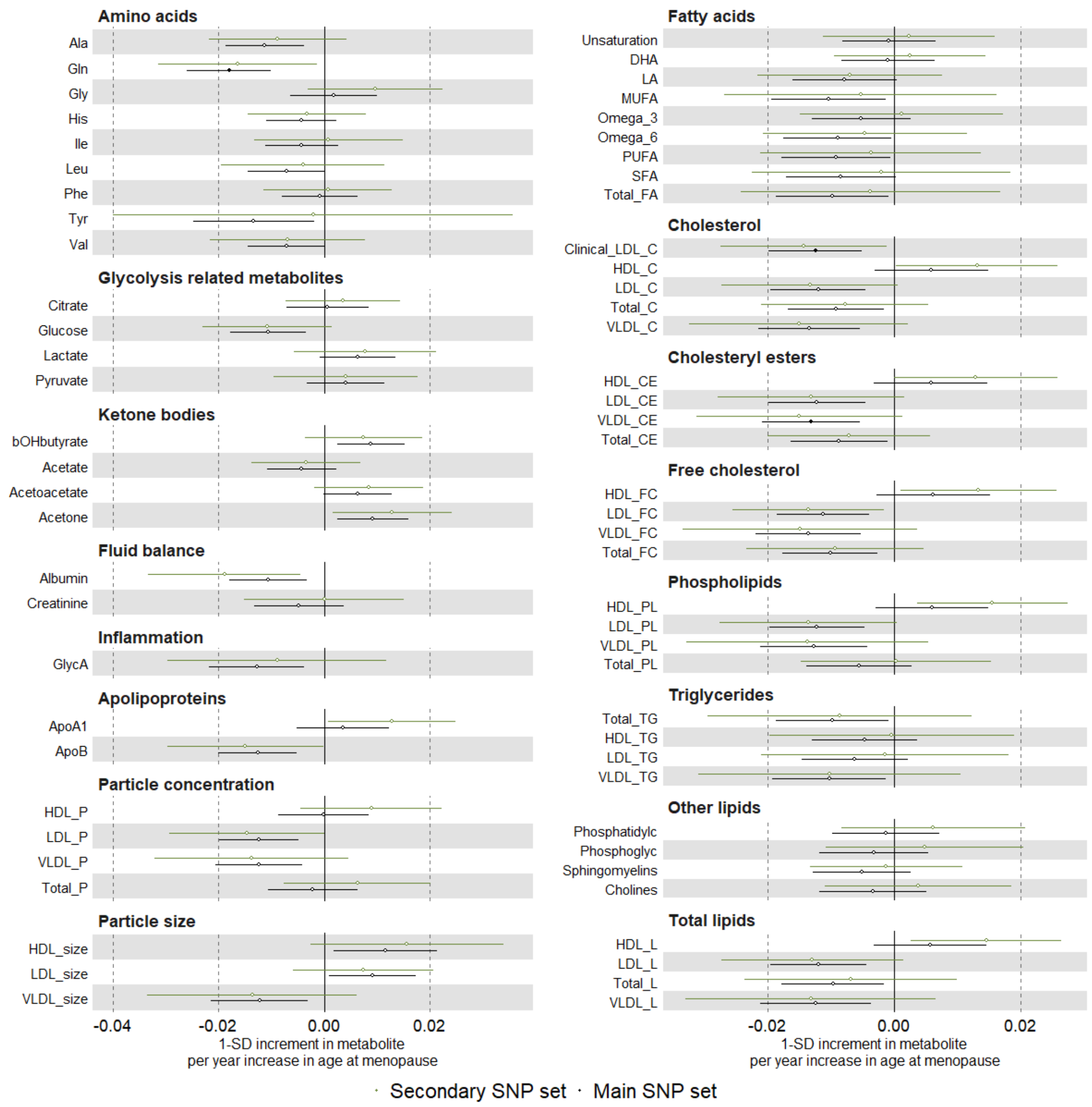

Suppl Fig 22. continued

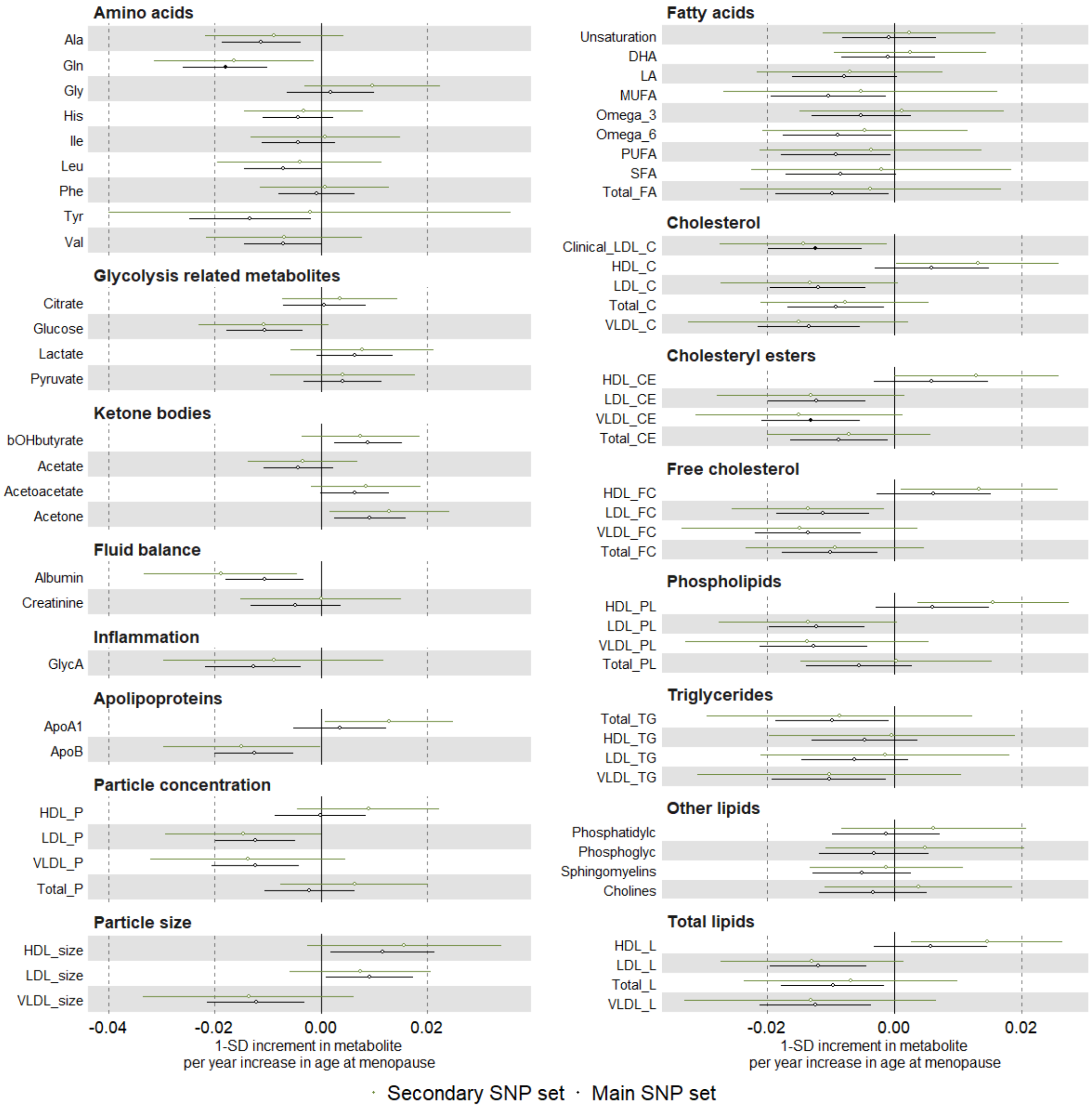

Suppl Fig 22. continued

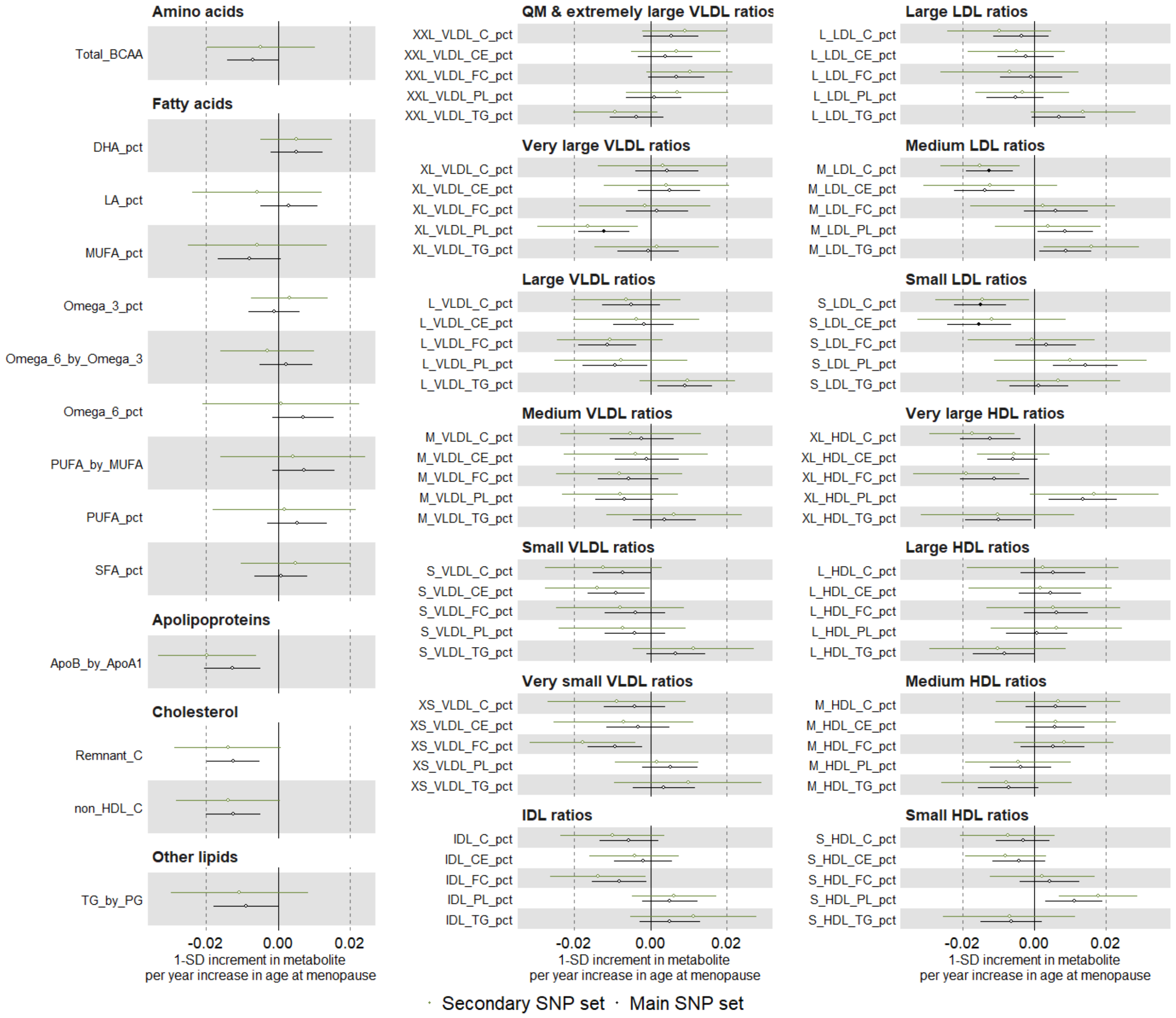
